# High definition longitudinal spatial atlas of psoriasis reveals SPRR2F⁺ keratinocytes and distinct tissue-resolution trajectories under IL-17 and IL-23 inhibition

**DOI:** 10.64898/2026.10.21.26365789

**Authors:** Tatiana Sezin, Nadia O. Prayitno, Yask Gupta, Sadegh Mousavi, Xinyi Tang, Daniel Bivona, Henner Zirpel, Sascha Ständer, David A. De Luca, Cristian Papara, Lena Pommerien, Diamant Thaҫi

**Affiliations:** University of Luebeck, Institute for Inflammation Medicine, Luebeck, Germany

**Author notes:** Authors contributed equally to the work (Tatiana Sezin, Nadia O. Prayitno, Yask Gupta).

## Abstract

How inflammation is established and resolved within tissues remains a fundamental question in human biology. Psoriasis is a chronic inflammatory skin disease driven by the IL-23/IL-17 pathway, with IL-17 pathway targeting biologics producing more rapid clinical improvement than IL-23 inhibitors (IL-23i). This difference raises the question of how IL-17 and IL-23 inhibition differentially remodel the cellular and transcriptional landscape of psoriatic skin during resolution. Here, we generated a longitudinal spatial atlas of plaque psoriasis using 2-µm high-definition spatial transcriptomics (HD-SPT), profiling 42 matched biopsies from 11 biologic treatment-naïve patients before and during IL-17- or IL-23-targeted therapy. By integrating HD-SPT with a single-cell RNA-sequencing (sc-RNAseq) reference atlas, we resolved the cellular composition and spatial organization of psoriatic skin and mapped its remodeling during treatment. We identified a distinct cluster of SPRR2F⁺ keratinocytes (KCs) that was spatially restricted to the upper stratum granulosum at the transition to the stratum corneum and was strongly associated with inflamed psoriatic epidermis. SPRR2F⁺ KCs were enriched for antimicrobial and inflammatory barrier genes, including *S100A8, S100A9, LCN2, IL36G*, and *PI3*, and their abundance correlated with disease activity during treatment. In psoriasis patient-derived human epidermal KCs (P-HEK), *SPRR2F* knockdown attenuated inflammatory and antimicrobial transcriptional responses, supporting a functional role for SPRR2F in sustaining the inflammatory KC response in psoriasis. Longitudinal profiling further revealed distinct trajectories of tissue remodeling following IL-17 and IL-23 inhibition. Following IL-17 inhibition, SPRR2F⁺ KCs and their characteristic transcriptional signature resolved rapidly, whereas IL-23i produced more gradual resolution of KC-centered transcriptional changes accompanied by broader immune and stromal remodeling. Together, these findings established the first HD-SPT atlas of a human skin disease and revealed distinct cellular and transcriptional trajectories of psoriatic skin during therapeutic resolution, identifying SPRR2F as a regulator of persistent KC inflammation.

## Introduction

Inflammation in chronic inflammatory skin diseases is a multicellular process that is established, maintained, and ultimately resolved through coordinated interactions among resident cells and cells that have migrated into the tissues across space and time^1^. Understanding how chronic inflammatory tissue states are established and dismantled requires insight into how cells are distributed and organized within tissues, how they communicate within and across tissue compartments, and also how these interactions are remodeled during a targeted therapeutic intervention^2,3^. Addressing these questions requires approaches that capture molecular heterogeneity while preserving tissue architecture across time.

Psoriasis provides an exceptional human model of chronic inflammatory skin disease with systemic implication where pathophysiology is well defined and can be followed prospectively and longitudinally using highly effective, pathway-targeted therapies^4^. A central driver of psoriatic inflammation is the IL-23/IL-17 axis, which forms a self-amplifying inflammatory circuit involving diverse immune and epidermal cells^5^. IL-23, produced predominantly by peripheral dendritic cells, promotes the activation, maintenance, and expansion of IL-17-producing T helper lymphocytes (TH-17)^6^. In turn, IL-17-family cytokines, particularly IL-17A, IL-17F, and IL-17C, act on keratinocytes (KCs) to induce inflammatory mediators and antimicrobial peptides (AMPs), alter epidermal barrier integrity, decrease the differentiation and promote the hyperproliferative epidermal phenotype characteristic of psoriasis^7^. Beyond their innate immune functions, certain AMPs may also contribute to adaptive immune activation by acting as autoantigens^8^. For example, LL37, an AMP overexpressed in psoriatic skin, has been identified as a target of autoreactive CD4⁺ and CD8⁺ T cells in patients with psoriasis^9^. KCs are therefore not passive targets of feed forward inflammation but active participants in feed-backward signaling that sustains continuous chronic inflammation in the skin. Therapeutic intervention with targeted inhibition of the IL-23/IL-17 axis consequently provides an opportunity to examine and understand how disruption at distinct points within the same inflammatory circuit shapes tissue resolution. Selective biologic treatments targeting IL-17A/F or IL-17RA and those targeting IL-23 can both achieve substantial improvement of clinical signs and symptoms, yet IL-17-targeted therapies generally in the first months of treatment lead to more rapid clinical improvement than IL-23 inhibition^10^. This difference raises the possibility that targeting distinct nodes of the same inflammatory circuit generates different trajectories of cellular and tissue resolution, rather than simply differing in the magnitude of inflammatory suppression.

Understanding these dynamics requires methods that preserve tissue architecture while capturing cellular and molecular heterogeneity at a single time point. Dissociative single-cell RNA-signaling (scRNA-seq) approaches can introduce quantitative transcriptional states at cellular resolution, but tissue dissociation removes the architectural spatial context in which these states are organized and from which their inter-cellular relationships derive biological meaning. Moreover, conventional sc-RNAseq pipelines may introduce an epidermal stratification bias, because highly differentiated KCs in the granular layers and above are relatively transcriptionally quiescent and therefore yield low unique molecular identifiers (UMI) counts and fewer detected gene features. As a result, these cells may be disproportionately excluded during standard quality-control filtering^11^. This limitation is especially relevant in psoriasis, where epidermal homeostasis is profoundly disrupted. Hyperproliferation and accelerated epidermal turnover drive rapid, abnormal early differentiation^12^. Consequently, KCs progress faster through the epidermal layers, reaching terminal states that can be systematically underrepresented by scRNA-seq approaches^13^. Conversely, conventional spot-based spatial transcriptomics preserves tissue architecture but lacks true single-cell resolution, such that individual capture regions can contain signals from multiple neighboring cells; in a highly stratified tissue such as the epidermis. This can mask transcriptional differences between adjacent layers and obscure layer-specific cellular states^14,15^. High-definition spatial transcriptomics (HD-SPT) addresses these complementary limitations by enabling quantitative transcriptomic measurements to be mapped across intact tissue at 2-µm resolution^16,17^. By combining preservation of tissue architecture with substantially greater spatial granularity, HD-SPT enables cellular populations and transcriptional changes to be interpreted within their anatomical context and allows tissue-wide remodeling to be precisely tracked during targeted therapeutic intervention.

To define how psoriatic skin is remodeled during therapeutic intervention, we generated a longitudinal spatial atlas using Visium HD-SPT (10x Genomics) across 42 matched skin biopsies from 11 previously biologic-naïve patients collected at baseline, week 2 and week 16 during treatment with anti-IL-17 or IL-23 biologics. Integration with a curated scRNA-seq reference atlas enabled high-confidence cell-type annotation, doublet identification, and spatial mapping of cell-specific transcriptional changes during therapeutic intervention. This analysis identified a previously undescribed population of SPRR2F⁺ KCs in lesional psoriatic skin that was characterized by expression of AMPs and inflammatory barrier-associated genes. Moreover, these cells were linked to disease activity during treatment and relapse. Direct comparison of the two therapeutic classes revealed a key divergence in tissue-resolution kinetics: SPRR2F⁺ KCs were rapidly depleted following IL-17inhibition but declined more gradually under IL-23 inhibition, mirroring differences in their therapeutic clinical response. Functional perturbation by siRNA-mediated *SPRR2F* knockdown in psoriasis patient-derived primary human KCs further showed that loss of *SPRR2F* attenuated cytokine-induced inflammatory responses.

Our findings identify for the first time a previously unrecognized role of SPRR2F⁺ KCs associated with psoriatic inflammation and provide functional evidence that SPRR2F contributes to immune mediated-inflammatory responses in KCs. Furthermore, they establish a cellular framework for understanding how perturbation at distinct points of the IL-23/IL-17 pathway differentially remodels chronic inflammatory tissue states and illustrate how longitudinal HD-SPT can connect molecular intervention with tissue-level remodeling in connection with clinical response.

## Results

### HD-SPT resolves the cellular and molecular architecture of plaque psoriasis

To characterize the cellular basis of differential therapeutic kinetics across the IL-23/IL-17 signaling pathway, we performed longitudinal HD-SPT of psoriatic skin during targeted inhibition IL-17 and IL-23 treatment.

We collected 42 full-thickness FFPE skin biopsies from 11 patients with moderate-to-severe plaque psoriasis (mean baseline PASI = 17.8 ± 3.3) before and during the treatment with ixekizumab (anti-IL-17A) or brodalumab an anti-IL-17RA-targeting biologic or with anti-IL-23p19-targeting biologics (guselkumab or risankizumab) (**Figure 1A, Supplementary Table 1**). Skin biopsies were obtained before treatment initiation from involved and uninvolved psoriatic skin (baseline-week 0), and thereafter only from lesional skin at week 2, and week 16, Consistent with established differences in clinical response kinetics, patients treated with IL-17A/IL-17RA-targeting biologics demonstrated more rapid improvement (at week 2) than those receiving IL-23i, with mean PASI reductions of 42% and 18% at week 2, respectively. By week 16, both treatment groups achieved substantial and comparable disease improvement, with mean PASI reductions of 88% and 78%, respectively (**Figure 1B, Supplementary Table 1**).

**Figure 1.**
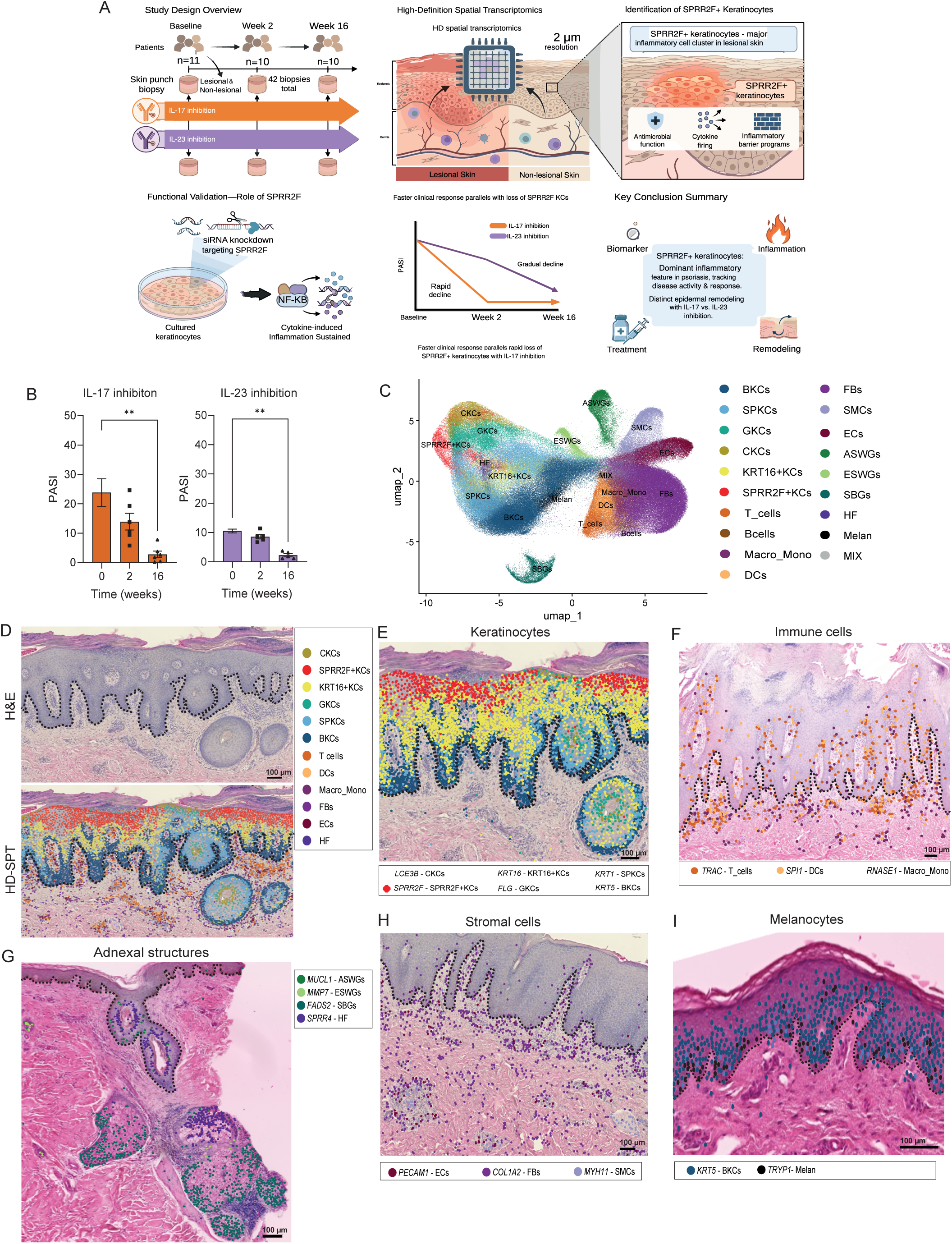
HD-SPT resolves the cellular and spatial architecture of plaque psoriasis. (A) Study design and sampling of longitudinal Visium HD spatial transcriptomic (HD-SPT) analysis. Lesional skin was sampled at baseline (week 0), week 2, and week 16 after treatment initiation, together with matched non-lesional skin at baseline only. Patients received IL-17A or IL-17RA-targeting biologics (brodalumab or ixekizumab; *n* = 6) or IL-23p19-targeting biologics (guselkumab or risankizumab; *n* = 5). Lower panel: mean PASI trajectory over the treatment course for each drug class (mean ± SEM). (B) Clinical treatment response measured by PASI in patients receiving IL-17A/IL-17RA inhibitors (left, *n* = 6) or IL-23 inhibitors (right, *n* = 5) at baseline (week 0), week 2, and week 16. Bars represent mean ± SEM; individual patients are shown as points. \*\**P* < 0.01 versus baseline; non-parametric Friedman test. (C) UMAP embedding of segmented cellular profiles from longitudinal Visium HD data. Clusters include: basal (BKCs), spinous (SPKCs), granular (GKCs), and cornified (CKCs) KCs; KRT16⁺ and SPRR2F⁺ KCs; T cells, B cells, dendritic cells (DCs), macrophages/monocytes (Macro_Mono); fibroblasts (FBs), smooth muscle cells (SMCs), endothelial cells (ECs); apocrine/eccrine sweat glands (ASWGs), eccrine sweat glands (ESWGs), sebaceous glands (SBGs), hair follicle (HF); and melanocytes (Melan). (D) Representative histology and spatial cell type mapping of lesional psoriatic skin. Top: H&E-stained tissue section. Bottom: corresponding spatial distribution of annotated cellular profiles overlaid on the same section, colored by cluster as indicated in the legend. Scale bar, 100 µm. (E–I) Spatial mapping of canonical marker genes on representative tissue sections, confirming that transcriptionally defined clusters occupy their expected anatomical niches. Each point denotes a segmented cellular profile expressing the indicated marker, colored by cluster as shown in the accompanying legends. Scale bars, 100 µm. The dashed line in D–I marks the dermal–epidermal junction.

To resolve the cellular composition of psoriatic skin at near-single-cell resolution, we generated Visium HD-SPT profiles from all 42 tissue sections. This generated 31,691,021 2 µm spatial bins, capturing up to 18,085 genes per section. H&E stained tissue images were segmented in-silico and bins within the segmented boundaries were clumped together, enabling reconstruction of near-single-cell transcriptomic profiles^18^. This approach initially generated 612,267 segmented cellular profiles. Following quality control and spatial filtering, 424,859 high-quality cellular profiles were retained. To identify and remove potential doublets, we integrated the HD-SPT dataset with a curated single-cell RNA-sequencing (scRNA-seq) reference atlas comprising from publicly available psoriatic skin datasets^19–23^ (**Supplementary Figure 1A, Supplementary Table 2-4**). Following reference-based annotation and doublet removal, we retained 379,753 high-quality spatially resolved cellular profiles spanning 14,194 genes across all samples.

Unsupervised clustering followed by marker gene analysis identified 18 transcriptionally distinct annotated clusters spanning epidermal, immune, stromal, adnexal, and melanocyte compartments, together with one additional mixed unannotated cluster (MIX), resulting in 19 total clusters (**Figure 1C-D; Supplementary Table 2**). To define the epidermal compartment, we examined KC clusters and clustered them into six transcriptionally distinct clusters corresponding to different stages of epidermal differentiation (**Figure 1E, Supplementary Figure 1B, Supplementary Table 2**). Basal KC clusters (BKCs) expressed canonical progenitor markers including *KRT5, KRT14,* and *COL17A1*. In contrast, spinous, granular, and cornified KC clusters (SPKCs, GKCs, and CKCs) demonstrated progressive activation of differentiation genes marked by *KRT1, KRT10, FLG, FLG2,* and late cornified envelope genes including *LCE2B, LCE6A,* and *LCE1B*. In addition to canonical differentiation-associated clusters, we identified two additional disease-associated KC clusters. The first corresponded to the previously described KRT16⁺ KC cluster, characterized by expression of *KRT16, KRT6A, KRT6B,* and *KRT17*. This cluster has previously been associated with KC hyperproliferation, aberrant differentiation, and inflammatory epidermal remodeling (7577575, 28576737). A second distinct KC cluster was transcriptionally separated from the KRT16⁺ KCs and other epidermal clusters. This cluster was characterized by selective expression of *SPRR2F* and *TGM3*, together with increased expression of *RHCG, PI3,* and *CXCL8*, defining an additional psoriasis-associated KC cluster.

Four immune clusters were identified (**Figure 1F, Supplementary Figure 1C, Supplementary Table 2**). T cell clusters expressed canonical T-cell receptor components including *TRAC, TRBC1,* and *TRBC2*, together with *LTB*. Macrophage/monocyte clusters expressed *RNASE1*, *C1QC, F13A1, STAB1,* and *LGMN*. Dendritic cell clusters were defined by expression of *LYZ, LCP1,* and *SPI1*. B cell clusters were identified by immunoglobulin transcripts including *IGKC, IGHG1,* and *IGHA1*, together with complement component *C3* (**Supplementary Figure 1C**).

Four adnexal clusters were identified (**Figure 1G, Supplementary Figure 1D, Supplementary Table 2**). Apocrine/eccrine sweat gland clusters (ASWGs) expressed *DCD, SCGB2A2,* and *PIP*. Eccrine sweat gland clusters (ESWGs) were characterized by expression of *MMP7* and *WFDC3*. Sebaceous gland clusters (SBGs) demonstrated enrichment of lipid metabolism-associated genes including *AWAT2, THRSP,* and *FADS2*. Hair follicle clusters (HF) expressed *SPRR4,* and *CST6*.

Three stromal clusters were resolved (**Figure 1H, Supplementary Figure 1E, Supplementary Table 2**). Fibroblast (FB) clusters expressed extracellular matrix-associated genes including *COL1A1, COL1A2, PI16, MFAP5,* and *ELN*. Smooth muscle cell (SMCs) clusters were defined by expression of contractile markers including *DES, MYH11, ACTG2,* and *ACTC1*. Endothelial cell (ECs) clusters expressed vascular markers including *VWF, PECAM1, PLVAP,* and *CCL14*. Melanocyte clusters were identified by canonical pigmentation-associated markers including *DCT, MLANA, TYRP1,* and *TYR* (**Figure 1I, Supplementary Figure 1F, Supplementary Table 2**).

In conclusion, these 19 spatially resolved transcriptional clusters establish a cellular framework for investigating how perturbation of distinct molecular components of the IL-23/IL-17 signaling pathway remodels psoriatic tissue during therapeutic response.

### SPRR2F⁺ KC expansion accompanies epidermal and dermal inflammatory remodeling in lesional psoriatic skin

To characterize the psoriatic skin composition before and during the treatment, we compared spatial cluster abundance and transcriptional programs between involved (lesional) and paired non-lesional skin.

The most evident remodeling occurred within the epidermal compartment. GKCs and SPKCs were significantly reduced in lesional skin, whereas KRT16⁺ and SPRR2F⁺ KC clusters were significantly expanded, with SPRR2F⁺ KCs strongly enriched in lesional skin (logFC=2.31, padj=2.6×10⁻^5^) (**Figure 2A, Supplementary Figure 2A, Supplementary Table 3**). Immunofluorescence (IF) validation confirmed that this increase represented a spatially distinct epidermal layer rather than a clustering artifact. SPRR2F⁺ KCs localized predominantly to the upper suprabasal epidermis, whereas KRT16⁺ KCs occupied overlapping but broader lower suprabasal regions (**Figure 2B**).

**Figure 2.**
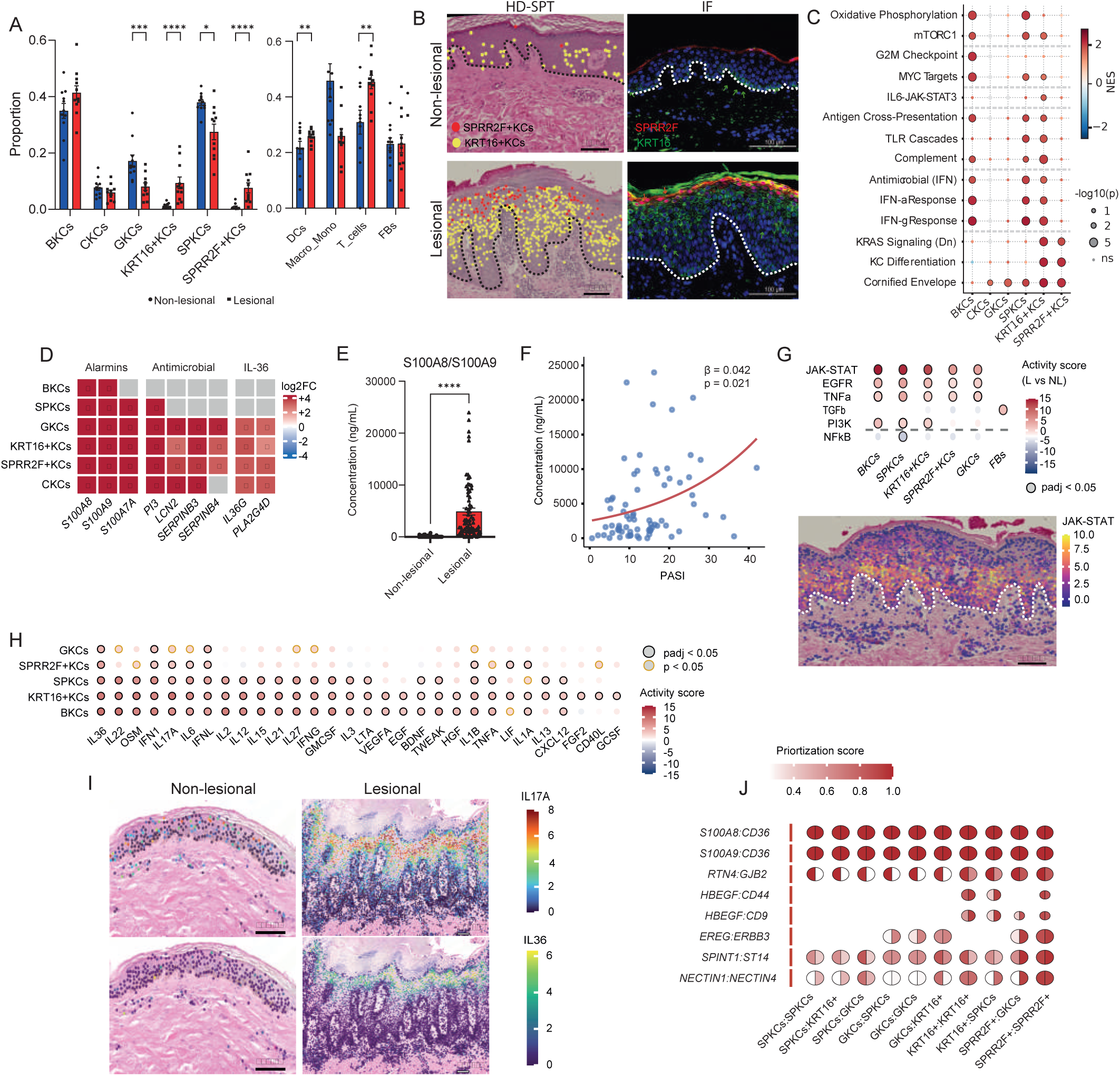
SPRR2F⁺ keratinocytes are enriched in the inflammatory psoriatic epidermis. (A) Cluster proportions in lesional versus paired non-lesional skin at baseline (*n* = 11 patients), quantified using propeller. Left: KC clusters. Right: immune and stromal clusters (DCs, Macro_Mono, T cells, FBs). Bars show mean proportion ± SEM; each point is one patient. Blue, non-lesional; red, lesional. \**P* < 0.05, \*\**P* < 0.01, \*\*\**P* < 0.001, \*\*\*\**P* < 0.0001; propeller test. (B) Representative non-lesional (top) and lesional (bottom) skin. Left: SPRR2F⁺KCs (red dots) and KRT16⁺KCs (yellow dots) overlaid on H&E. Right: immunofluorescence for SPRR2F (red) and KRT16 (green) with DAPI nuclear counterstain (blue). A dashed white line marks the dermal– epidermal junction. Scale bars, HD-SPT: 100 µm; IF: 100 µm. (C) Gene set enrichment in lesional versus non-lesional skin across KC clusters. Dot color indicates normalized enrichment score (NES); dot size indicates −log₁₀(*P*); ns, not significant. (D) Differential expression (log₂ fold change, lesional versus non-lesional) of alarmin, antimicrobial and IL-36-axis genes across KC clusters. Color scale runs from −4 (blue) to +4 (red); grey denotes genes not tested or not significant. Asterisks mark significant changes (lfsr < 0.05). (E) S100A8/S100A9 protein concentration in lesional versus non-lesional skin-rinsing samples from an independent cohort (*n* = 72 patients, 144 samples). Bars show mean ± SEM; each point is one sample. \*\*\*\**P* < 0.0001, non-parametric Wilcoxon test. (F) S100A8/S100A9 concentration versus PASI across all samples; red line shows the fitted regression (β = 0.042, *P* = 0.021). (G) Top: PROGENy pathway activity in lesional versus non-lesional skin by cluster. Dot color indicates activity score; open black circles mark adjusted *P* < 0.05. Bottom: spatial distribution of JAK-STAT1 activity on a representative lesional section, colored from dark blue (low) to red (high). (H) CytoSig cytokine-response activity in lesional versus non-lesional skin across KC clusters. Dot color indicates activity score; grey circles mark adjusted *P* < 0.05 and open circles unadjusted *P* < 0.05. (I) Spatial distribution of inferred IL-17A (top) and IL-36 (bottom) response activity on representative non-lesional and lesional sections, colored from dark blue (low) to yellow (high). Scale bars, 100 µm. (J) MultiNicheNet ligand–receptor interactions prioritized in lesional versus non-lesional skin, by sender:receiver cluster pair (sender left half of the circle; receiver right half of the circle). Red denotes interactions upregulated in lesional skin and blue those that are attenuated; dot size indicates prioritization score.

To further define the molecular identity of SPRR2F⁺ KCs expanded in lesional epidermis, we examined their transcriptional profiles. SPRR2F⁺ KCs displayed one of the strongest transcriptional shifts among KC clusters, with 942 of 10,021 genes significantly upregulated (logFC > 0; lfsr < 0.05) compared with non-lesional skin (**Supplementary Fig. 2B, Supplementary Table 4**). Their transcriptional program was dominated by epidermal differentiation and barrier remodeling pathways, including cornified envelope formation (padj = 1.38 × 10⁻⁷), keratinization (padj = 1.38 × 10⁻⁷), and integumentary developmental programs (padj = 4.9 × 10⁻⁴), together with KRAS signaling and neutrophil degranulation modules (**Figure 2C, Supplementary Table 5**).

In parallel, SPRR2F⁺ and KRT16⁺ KCs shared activation of psoriasis-associated inflammatory programs, including S100-family alarmins (*S100A8, S100A9, S100A7A*), epithelial defense molecules (*PI3, LCN2, SERPINB3, SERPINB4*), and IL-36 pathway mediators (*IL36G, PLA2G4D*) (**Figure 2D, Supplementary Table 4**). However, compared to SPRR2F⁺ KCs, KRT16⁺ KCs exhibited a broader inflammatory transcriptional response including interferon, innate immune, cytokine-effector, proliferative, and metabolic pathways, suggesting distinct inflammatory signatures across these disease-associated KC clusters (**Figure 2C, Supplementary Table 5**).

Among these shared mediators, the alarmins S100A8 and S100A9 are secreted by activated KCs and accumulate at the skin surface, making them accessible to non-invasive sampling and thus well suited to test whether this transcriptional signature is reflected at the protein level. To this end, we analyzed skin rinsing samples from an independent cohort of 72 patients with moderate-to-severe plaque psoriasis (**Supplementary Table 6**). This orthogonal approach confirmed increased lesional S100A8/S100A9 protein levels **(Figure 2E**) and demonstrated significant correlation with PASI score (padj=0.021) (**Figure 2F**), supporting the translational relevance of the inflammatory program associated with SPRR2F⁺ KCs.

To determine whether these transcriptional differences reflected distinct signaling environments. PROGENy^24^ pathway activity inference and scoring methodology are described in **Methods**. JAK-STAT signaling emerged as the dominant activated pathway across lesional KC clusters (**Figure 2G, Supplementary Table 7**). Activity was highest in BKCs (score=15.4) and SPKCs (score=14.0), which are positioned near the cytokine-rich dermal-epidermal interface, and progressively decreased toward SPRR2F⁺ KCs (score=6.4) and GKCs (score=5.7). TNFα, PI3K, and EGFR signaling were similarly activated across overlapping KC clusters, indicating broad inflammatory pathway convergence within the lesional epidermis.

CytoSig-based^25^ cytokine-response analysis revealed distinct inflammatory programs among lesional KC clusters (**Figure 2H-I, Supplementary Table 7**). SPRR2F⁺ KCs were enriched for canonical psoriasis-associated cytokine pathways, including IL-17A, IL-36, IL-1α, IL-1β, IL-6, type I interferon (IFN1), type III interferon (IFN-λ), and LIF signaling. In contrast, KRT16⁺ KCs displayed a broader cytokine-response landscape incorporating IL-22, IFN-γ, IL-12, and IL-27 activity. BKCs and SPKCs similarly demonstrated broad Th1/interferon-associated activation. Notably, BKCs, SPKCs, and KRT16⁺ KCs showed suppression of TGFβ-superfamily signaling, whereas SPRR2F⁺ KCs and GKCs retained a more restricted cytokine-response profile. Together, these data define two distinct inflammatory epidermal programs: a broad interferon-associated response in lower epidermal clusters and a more focused IL-17/IL-36-associated program in upper epidermal SPRR2F⁺ and GKCs.

Beyond transcriptional identity and signaling activity, we examined whether SPRR2F⁺ KCs were associated with altered local cellular communication. MultiNicheNet^26^ ligand-receptor prioritization analysis showed increased intercellular signaling in lesional skin, including *S100A8/S100A9– CD36* interactions across multiple KC clusters (**Figure 2J, Supplementary Table 8, Supplementary Fig. 2C**). SPRR2F⁺ KCs were the main source of prioritized outgoing interactions, including some of the highest-scoring autocrine interactions (*S100A8/S100A9– CD36*, *EREG–ERBB2* and *SPINT1–ST14*), highlighting SPRR2F⁺ KCs as a prominent source of inflammatory signaling within the epidermis, whereas no SPRR2F⁺ interactions were prioritized in non-lesional skin.

Inflammatory remodeling extended beyond the epidermis into the dermal compartment. T cells (logFC=1.13, padj=2.1×10⁻³) and DCs (logFC=0.76, padj=2.1×10⁻³) were expanded and transcriptionally activated in lesional skin (**Figure 2A, Supplementary Table 3-4**). Since spatial proximity can introduce transcriptional overlap between neighboring dermal clusters, all dermal transcriptional programs were cross-validated against an independent in-house psoriatic skin single-cell atlas (**Supplementary Figure 1A**).

Lesional T cells acquired inflammatory, interferon-responsive, and skin-homing transcriptional programs, while DCs showed increased antigen presentation and T-cell recruitment signatures (**Supplementary Figure 2D, Supplementary Table 4**). Although macrophage/monocyte abundance was unchanged, these cells demonstrated transcriptional activation of complement and myeloid effector programs (**Supplementary Figure 2D, Supplementary Table 4**). FBs were reduced in abundance but underwent marked extracellular matrix remodeling, including collagen expression and activation of TGFβ signaling (score=3.8, padj=2.3×10⁻^4^) **Supplementary Figure 2E, Supplementary Table 3 and 7**).

These findings establish an inflammatory cascade in psoriasis as a coordinated tissue-wide inflammatory ecosystem involving epidermal, immune, and stromal remodeling. FBs undergo TGFβ-associated extracellular matrix remodeling, while immune compartments acquire activated inflammatory and antigen-presentation programs. Within the epidermal compartment, disease-associated KRT16⁺ and SPRR2F⁺ KC clusters exhibit distinct inflammatory profiles, with SPRR2F⁺ KCs representing a prominent upper-epidermal component characterized by alarmin production and antimicrobial barrier remodeling.

### KC-centered transcriptional resolution precedes restoration of tissue composition during IL-17 inhibition

To investigate how IL-17 inhibition remodels lesional psoriatic tissue over time, we applied the spatial framework to serial biopsies collected at weeks 2 and 16 from patients treated with ixekizumab or brodalumab, revealing a two-phase, KC-centered resolution pattern characterized by early transcriptional changes followed by broader restoration of tissue composition (**Figure 1A**).

At week 2, no cellular cluster showed significant compositional contraction, although SPRR2F⁺ KCs displayed the strongest trend toward reduction (logFC =−0.82, padj =0.16) (**Figure 3A-B, Supplementary Figure 3A, Supplementary Table 9**) compared to baseline. Instead, the earliest response was characterized by transcriptional remodeling across disease-associated KC clusters. SPRR2F⁺, KRT16⁺, and GKCs showed suppression of the lesional alarmin– antimicrobial–IL-36-associated transcripts, including *S100A8*, *PI3*, *LCN2*, and *IL36G* (**Figure 3C, Supplementary Figure 3B, Supplementary Table 10**). Consistent with these transcriptional changes, S100A8/S100A9 protein levels measured in independent skin rinsing samples significantly decreased by week 2, preceding major compositional and clinical resolution (**Figure 3D, Supplementary Table 6**).

**Figure 3.**
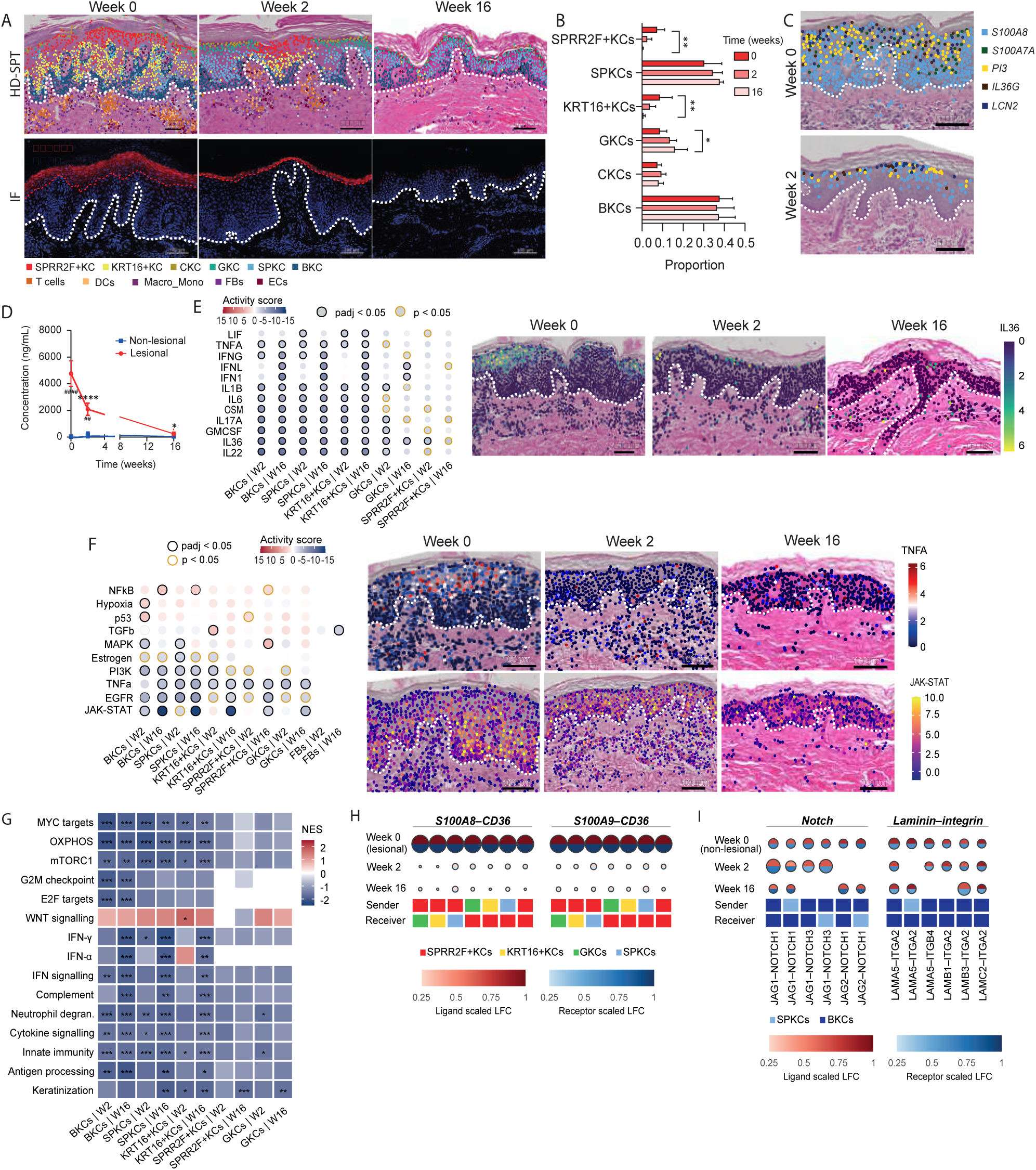
IL-17 inhibition drives rapid keratinocyte-centered resolution of psoriatic inflammation. (A) Representative lesional skin at baseline (week 0), week 2 and week 16 under IL-17/IL-17RA inhibition. Top: annotated cellular profiles overlaid on H&E, colored by cluster as in the key. Bottom: immunofluorescence for SPRR2F (red) with DAPI nuclear counterstain (blue). Scale bars, 100 µm. Right: KC cluster proportions across the three time points (*n* = 11 patients), quantified using propeller. Bars show mean proportion ± SEM; *padj < 0.05, **padj < 0.01 (propeller test). (B) Spatial distribution of lesional alarmin, antimicrobial and IL-36-axis transcripts at week 0 (top) and week 2 (bottom) on representative sections. Each point is a segmented profile expressing the indicated gene: *S100A8* (light blue), *S100A7A* (green), *PI3* (yellow), *IL36G* (dark red), *LCN2* (dark blue). (C) S100A8/S100A9 protein concentration in lesional (red circles) and non-lesional (blue squares) skin-rinsing samples across the treatment course in the independent cohort (*n* = 72 patients). Data are mean ± SEM. ####*P* < 0.0001 lesional versus non-lesional at baseline; ##adjusted *P* < 0.01 versus non-lesional at the specific time point; \**P* < 0.05, \*\*\*\**P* < 0.0001 versus baseline (mixed-effects model (REML) with Šidák’s multiple comparisons test). (D) Spatial distribution of inferred IL-36 response activity on representative lesional sections at weeks 0, 2 and 16, colored from dark blue (low) to yellow (high). Scale bars, 100 µm. (E) CytoSig cytokine-response activity at week 2 and week 16 versus baseline, by KC cluster. Dot color indicates activity score, from red (increased) to blue (decreased); grey circles mark adjusted *P* < 0.05 and yellow circles unadjusted *P* < 0.05. (F) Left: PROGENy pathway activity at week 2 and week 16 versus baseline, by cluster. Dot color indicates activity score, from red (increased) to blue (decreased); black circles mark adjusted *P* < 0.05 and yellow circles unadjusted *P* < 0.05. Right: spatial distribution of inferred TNFα (top) and JAK-STAT (bottom) activity on representative lesional sections at weeks 0, 2 and 16. Scale bars, 100 µm. (G) Gene set enrichment at week 2 and week 16 versus baseline, by KC cluster. Color indicates normalised enrichment score (NES), from red (enriched) to blue (depleted). The dashed line separates proliferative and metabolic gene sets (above) from inflammatory and barrier gene sets (below). *padj < 0.05, **padj < 0.01, ***P < 0.001. (H) MultiNicheNet ligand–receptor interactions after IL-17 inhibition at baseline (lesional skin), week 2 and week 16 compared to baseline. Compared to baseline lesional-enriched interactions *S100A8-CD36* and *S100A9-CD36* were reduced after treatment. In contrast, Notch-JAG and laminin-integrin interactions, which were more abundant in non-lesional skin were induced after treatment (I). Colored squares indicate the sender and receiver clusters. Each dot is split into the scaled ligand LFC (top, red) and receptor LFC (bottom, blue); dot size indicates prioritisation score.

Cytokine activity inference revealed rapid reversal of inflammatory cytokines across KC clusters (**Supplementary Table 11**). In SPRR2F⁺ KCs and GKCs, IL36 signaling was among the earliest to normalize, whereas BKCs, SPKCs, and KRT16⁺ KCs showed broader suppression of lesional cytokine activities. Across these clusters, the majority of baseline-enriched cytokine-associated pathways were significantly reversed by week 2 (BKCs: 33/36, 92%; SPKCs: 27/30, 90%; KRT16⁺ KCs: 28/36, 78%), including IL17A, IL22, IL36, OSM, IL6, and TNFα signaling (**Figure 3E, Supplementary Table 11**). In parallel, upstream pathway analysis demonstrated reversal of inflammatory signaling activity, including TNFα, EGFR, MAPK, PI3K, and JAK-STAT across lesional KC clusters (**Figure 3F**). In KRT16⁺ KCs, suppression of inflammatory signaling was accompanied by reduction of proliferative and metabolic pathways, including MYC signaling, oxidative phosphorylation, and mTORC1 activity, suggesting early restoration of epidermal homeostasis (**Figure 3G, Supplementary Table 12**).

By week 16, the early molecular changes were accompanied by broader restoration of tissue composition. SPRR2F⁺ and KRT16⁺ KC clusters significantly contracted (logFC=−0.19 and −0.20, respectively; padj = 2.9 × 10⁻³ for both), accompanied by reductions in T cells (logFC = −0.04, padj = 5.7 × 10⁻⁴) and DCs (logFC =−0.03, padj =4.8 × 10⁻³). In parallel, GKCs increased in abundance (logFC=0.04, padj=0.02), together indicating a shift towards the cellular composition of non-lesional skin (**Figure 3A-B, Supplementary Table 9**).

Treatment response at the end of induction phase (week 16) was marked by progressive normalization and restoration of epidermal landscape. Across KC clusters, interferon, innate immune, complement, cytokine-response, and antigen-processing-related pathways remained suppressed (**Supplementary Table 13**). SPRR2F⁺ KCs showed one of the most extensive transcriptional remodeling, with significant (padj<0.05) differentially expressed genes increasing from 179 at week 2 to 763 at week 16 and reversing 56.8% (535/942) of the upregulated genes induced during lesional disease (**Supplementary Table 10 and 14, Supplementary Figure 3C**). Despite fewer induced genes overall, SPRR2F⁺ and KRT16⁺ KCs exhibited larger per-gene effect sizes, suggesting that resolution was characterized by a more focused and intensified transcriptional program (**Supplementary Figure 3D**).

Upstream cytokine analysis confirmed suppression of IL-17A signaling in KRT16⁺ KCs (score = −4.97, padj= 2.27 × 10⁻⁶), accompanied by resolution of interferon-associated activity, including IFN-γ, IFN-α, and IFN-λ signaling (**Figure 3E, Supplementary Table 15**). JAK-STAT activity was also strongly reduced in KRT16⁺ KCs by week 16 (score =−13.35, padj= 9.96 × 10⁻³⁹), consistent with reversal of the JAK-STAT activation observed in lesional skin at baseline (**Figure 3F, Supplementary Table 15**).

Spatial communication analysis showed parallel remodeling of epidermal-associated communication during IL-17 inhibition (**Supplementary Table 16**). At a prioritization score ≥ 0.75, non-lesional-enriched interactions were prioritized more after treatment (43% at week 2, 62% at week 16), whereas lesional-enriched interactions were reduced (38% and 52%) and rarely higher (3% and <1%), indicating a shift towards the non-lesional communication pattern. S100A8/S100A9–CD36 interactions, enriched in lesional skin at baseline, were reduced at both weeks, and by week 16 most lesional-enriched interactions involving SPRR2F⁺ KCs were also reduced (**Figure 3H**). In contrast, interactions including Notch (JAG–NOTCH) and laminin– integrin signaling, which are enriched in non-lesional skin, were prioritized after treatment, consistent with a shift towards basal keratinocyte differentiation and basement membrane engagement (**Figure 3I, Supplementary Table 16**).

Resolution extended beyond the epidermis to the immune and stromal compartments (**Supplementary Figure 3F**). T cells and DCs progressively lost inflammatory features, including reduced expression of genes associated with migration, antigen presentation, and chemokine signaling (**Supplementary Table 10**). In DCs, *CCL19* showed the strongest decline among immune-related genes, consistent with a reduction in the mature migratory DC phenotype (**Supplementary Table 10**). FBs regained markers of quiescence that were reduced during disease, including *PI16*, *CYBRD1*, *MGP*, and *MFAP4*, followed by broader resolution of stromal activation at the pathway level (**Supplementary Table 13**). By week 16, TGFβ signaling was significantly reduced (score = −2.65, padj = 0.009; **Supplementary Table 15**), accompanied by suppression of extracellular matrix remodeling genes, including *COL6A3* and *C1S* (**Supplementary Table 14**).

Collectively, IL-17 inhibition induced a predominantly KC-centered resolution response that extended across immune and stromal compartments. Early transcriptional changes at week 2 preceded broader restoration of tissue composition by week 16, indicating that treatment response is characterized predominantly by progressive normalization of pre-existing inflammatory gene signature rather than the emergence of new transcriptional patterns.

### IL-23i induce delayed and incomplete resolution of SPRR2F⁺ KCs with distinct stromal and immune remodeling compared to IL-17 inhibition

To determine whether IL-23 inhibition resolves psoriatic skin through the same cellular sequence as IL-17 inhibition or follows a distinct trajectory, we applied the same spatial framework to serial biopsies from patients treated with guselkumab (n = 3) or risankizumab (n = 3) and compared responses across cellular clusters at matched time points (**Figure 1A**).

At week 2, no cellular cluster showed significant compositional change under IL-23 inhibition, mirroring the early response to IL-17inhibition (**Figure 4A-B, Supplementary Fig. 4A, Supplementary Table 9**). By week 16, however, the cellular response was characterized by significant contraction of T cells (logFC = −0.03, padj= 0.0002) and DCs (logFC = −0.02, padj= 0.007), together with expansion of FBs (logFC = 0.02, padj= 0.007). SPRR2F⁺ and KRT16⁺ KCs also declined numerically between baseline and week 16 (logFC = -0.14 and logFC = -0.11, respectively), but neither reached statistical significance, indicating that compositional resolution of the inflammatory KC clusters was delayed relative to IL-17 inhibition (**Figure 4A-B, Supplementary Fig. 4A**).

**Figure 4.**
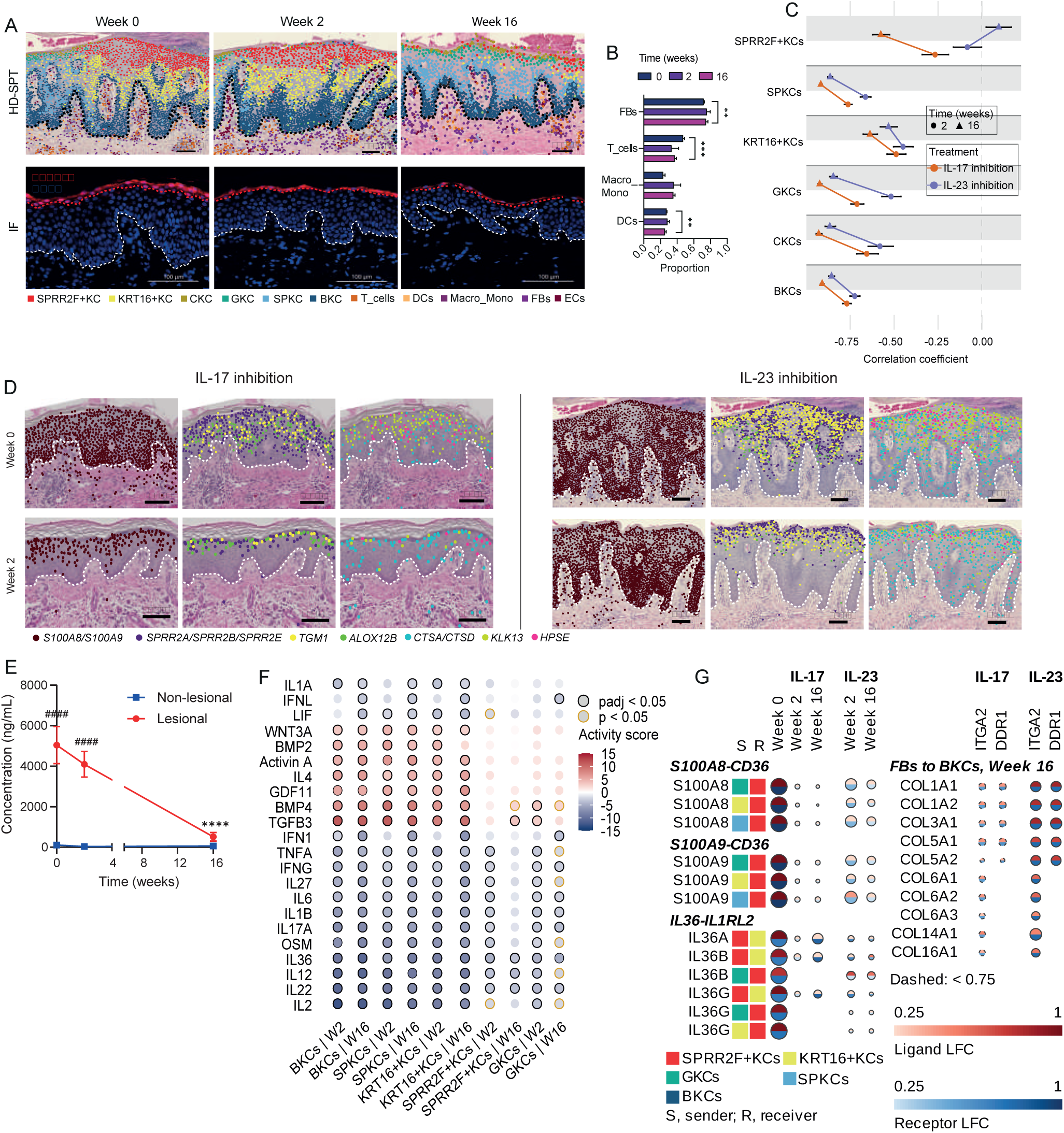
IL-23 inhibition induces distinct tissue remodeling compared with IL-17 inhibition. (A) Representative lesional skin at weeks 0, 2 and 16 under IL-23 inhibition. Top: annotated cellular profiles overlaid on H&E, colored by cluster as in the key. Bottom: immunofluorescence for SPRR2F (red) with DAPI nuclear counterstain (blue). Scale bars, 100 µm. Dashed black line in HD-SPT images and dashed white line in IF images marks the dermal–epidermal junction. (B) Relative abundance of immune and stromal clusters (DCs, Macro_Mono, T cells, FBs) at baseline (week 0), week 2 and week 16 under IL-23 inhibition (*n* = 5 patients), quantified using propeller. Bars show mean ± SEM; **Padj < 0.01, ***Padj < 0.001. (C) Correlation between baseline lesional expression and treatment response, by KC cluster. Circles denote week 2 and triangles week 16; orange, IL-17/IL-17RA inhibition; purple, IL-23 inhibition. More negative coefficients indicate stronger correction of the baseline lesional transcriptional profile. Horizontal lines connect the two time points within a treatment arm. (D) Spatial expression of lesional genes at week 0 and week 2 in representative sections under IL-17 inhibition (ixekizumab, left) and IL-23 inhibition (guselkumab, right). Each dot marks a segmented profile expressing the indicated gene(s): *S100A8/S100A9* (brown), *SPRR2A/SPRR2B/SPRR2E* (dark blue), *TGM1* (yellow), *ALOX12B* (green), *CTSA/CTSD* (turquoise), *KLK13* (light green), *HPSE* (magenta). Genes sharing a color are shown combined, and a dot marks a profile expressing at least one of them. Where gene genes are co-expressed, dots overlap and only the uppermost is visible. Dashed white line, dermal–epidermal junction. Scale bars, 100 µm. (E) S100A8/S100A9 protein concentration in lesional (red circles) and non-lesional (blue squares) skin-rinsing samples across the treatment course under IL-23 inhibition (*n* = 31 patients). Data are mean ± SEM. ####*P* < 0.0001 lesional versus non-lesional; \*\*\*\**P* < 0.0001 versus baseline; mixed-effects model (REML) with Šidák’s multiple comparisons test. (F) CytoSig cytokine-response activity at week 2 and week 16 versus baseline under IL-23 inhibition, by KC cluster. Dot colour indicates activity score, from red (increased) to blue (decreased); black circles mark adjusted *P* < 0.05 and yellow circles unadjusted *P* < 0.05. (G) MultiNicheNet ligand–receptor interactions at baseline (Base; lesional vs non-lesional) and at weeks 2 and 16 versus baseline under IL-17 inhibition and IL-23 inhibition. Left: S100A8–CD36, S100A9–CD36 and IL-36–IL1RL2 interactions between KC clusters; colored squares indicate the sender (S) and receiver (R) clusters. Right: collagen–ITGA2 and collagen– DDR1 interactions from FBs to BKCs prioritized at week 16. Each dot is split into the scaled ligand LFC (top, red) and receptor LFC (bottom, blue); dot size indicates [prioritization score]; dashed outlines mark scores < 0.75.

At the transcriptional level, the week 2 response differed less in the number of genes reversed than in the magnitude of their correction (**Supplementary Fig. 4B, Supplementary Table 17)**. Specifically, in SPRR2F⁺ KCs, a comparable fraction of up-regulated lesional genes was already reversed by both treatments (8.6%, 81/942 under IL-23 inhibition versus 10.9%, 103/942 under IL-17 inhibition), although the magnitude and direction of the response began to diverge between treatment arms already at this early time point (**Figure 4C**, **Supplementary Table 17, Supplementary Fig. 4B**). Under IL-17 inhibition, genes most strongly dysregulated in lesional skin showed the largest corrective changes at week 2 (r = −0.27, n = 592), whereas this relationship was not yet apparent under IL-23 inhibition (r=−0.08), indicating that early correction of the lesional transcriptional signature was more pronounced with direct IL-17 inhibition than IL-23 inhibition (**Figure 4C**). Consistent with this distinction, alarmins including *S100A8*, *S100A9*, and *S100A12*, keratinization genes including *TGM1*, *ALOX12B*, *SPRR2A*, *SPRR2B*, and *SPRR2E*, and proteases including *KLK13*, *HPSE*, *CTSD*, and *CSTA* were suppressed by week 2 under IL-17 inhibition but remained largely unchanged under IL-23 inhibition (**Figure 4D, Supplementary Table 17**). Consistent with these transcriptional changes, S100A8/S100A9 protein levels in skin-rinsing samples from an independent cohort remained unchanged at week 2 under IL-23 inhibition compared with IL-17 inhibition, but declined by week 16, further supporting delayed resolution observed after IL-23i (**Figure 4E, Supplementary Table 6**).

Despite these differences in kinetics, the two treatments converged on many of the same inflammatory pathways. At week 2, IL-36 signaling was significantly suppressed under both IL-17 and IL-23 inhibition across all KCs, while IL-23 inhibition additionally reduced IL-17A, IL-1β, and IL-6 signaling (**Figure 3E**, **Figure 4F, Supplementary Fig. 4C, Supplementary Table 11 and 19**).

In SPRR2F⁺ KCs, at week 2, IL-23 inhibition was associated with significant changes across 22 cytokine pathways compared with only 2 under IL-17 inhibition (padj< 0.05; **Supplementary Table 11 and 19)**; however, the underlying cytokine activity scores were strongly correlated between treatments (r=0.79, **Supplementary Fig. 4D**), indicating that the greater number of significant pathways under IL-23 likely reflected differences in statistical power rather than fundamentally distinct cytokine responses.

At week 2, KRT16⁺ KCs showed a similar degree of transcriptional convergence, with 17.9% (143/799) and 22.0% (176/799) of baseline-upregulated genes reversed under IL-23 and IL-17 inhibition (**Supplementary Fig. 4E**), respectively, together with comparable suppression of the alarmin–IL-36 axis and proliferative, metabolic, and inflammatory genes (**Supplementary Table 17**). GKCs showed a larger difference in gene reversal (11.6%, 71/610, versus 19.0%, 116/610) (**Supplementary Fig. 4F**), while the remaining KC clusters showed moderate-to-strong inverse correlations between baseline lesional expression and week-2 response under IL-23 inhibition (r = −0.45 to −0.72; **Supplementary Fig. 4G**).

By week 16, SPRR2F⁺ KCs have emerged as the principal point of divergence between the two therapeutic trajectories. IL-17 targeting biologics reversed 56.8% (535/942) of genes induced in lesional SPRR2F⁺ KCs, compared with 6.2% (58/942) under IL-23i (**Supplementary Table 22, Supplementary Fig. 4H**). The transcriptional response was characterized predominantly by changes in cornified-envelope and keratinization gene families, including *KLK*, *KRT*, *LCE*, and *SPRR*, together with *CDSN* and *TGM3*. These genes were suppressed following IL-17 inhibition, whereas their expression remained largely unchanged or increased under IL-23 inhibition (**Supplementary Table 22, Supplementary Fig. 4H)**. Accordingly, cornified-envelope and keratinization gene sets remained enriched in SPRR2F⁺ KCs after IL-23 inhibition (NES = 1.87, padj= 0.003), indicating that IL-23i at week 16 did not fully resolve the expression of these genes compared to lesional skin at baseline (**Supplementary Table 23**). Furthermore, the baseline lesional expression remained inversely associated with week-16 expression under IL-17 inhibition (r =−0.57), but not under IL-23i (r=0.10, padj= 0.22), further distinguishing the transcriptional trajectory of SPRR2F⁺ KCs (**Figure 4C**). Disease-relevant signaling activity showed a similar pattern, with JAK-STAT activity significantly reduced under IL-17 inhibition (padj= 0.027) but not under IL-23 inhibition (padj = 0.092), despite moderate overall correlation between treatment arms (r = 0.395; **Supplementary Table 15 and 24, Supplementary Fig. 4I**).

In contrast, the remaining KC clusters largely converged by week 16. In BKCs, SPKCs, and GKCs, lesional pathways including JAK-STAT, EGFR, PI3K, TNFα, and IL-36 were suppressed to similar magnitudes under both treatments, accompanied by comparable reversal of baseline transcriptional profiles (**Supplementary Table 24, Supplementary Fig. 4J**). The inverse relationship between baseline lesional expression and week-16 response in all KCs clusters but SPRR2F+ KCs strengthened under both treatments (r =−0.53 to −0.93), indicating progressive normalization of the shared lesional transcriptional signature (**Figure 4C**, **Supplementary Fig. 4K**).

Inferred communication between KC clusters showed a similar, though less pronounced, shift under IL-23i than under IL-17 inhibition (**Supplementary Figure 4L, Supplementary Table 25**). Consistent with this, fewer interactions enriched in non-lesional skin were prioritized after IL-23i treatment, reaching 44% at week 16 compared with 62% following IL-17 inhibition. Interactions involving SPRR2F⁺ KCs were reduced to a similar overall extent in both treatment arms, but the affected signaling programs differed. By week 16, the CD36-centred alarmin axis was more extensively reduced following IL-17 inhibition, with all eight *S100A8/S100A9–CD36* interactions involving SPRR2F⁺ KCs as receivers reduced, compared with four under IL-23i. In contrast, IL-36 (*IL36–IL1RL2*) interactions were reduced almost exclusively under IL-23i (**Figure 4G**). Thus, IL-23 inhibition remodeled KC communication more gradually than IL-17 inhibition, with alarmin-associated signaling around SPRR2F⁺ KCs reduced later and less completely.

Outside the KC compartment, IL-23i produced a distinct stromal response. At week 2, FBs showed pronounced recovery, including increased expression of *PI16* (logFC = +0.98 versus +0.57 under IL-17 inhibition), together with *MFAP4*, *PRELP*, and *ADAM33*, and induction of genes associated with extracellular matrix remodeling and fibrillar organization, including *CTSK*, *FSTL1*, *CCDC80*, and *ELN* (**Supplementary Table 10 and 17, Supplementary Fig. 4M**). Recovery of *PI16* and *ELN* persisted at week 16 (logFC = +0.71 and +0.66, respectively), accompanied by suppression of TGFβ signaling (score = −1.81, padj= 0.07) (**Supplementary Table 21 and 24**). Consistent with this stromal remodeling, FB to KC signaling remained prioritized under IL-23i through week 16, mainly through collagen–integrin and collagen–DDR1 interactions from FBs to BKCs (including *COL1A1/COL1A2/COL3A1–ITGA2* and *COL14A1–ITGA2*), whereas under IL-17 inhibition FB to KC interactions were prioritized at week 2 but fewer remained by week 16 (**Supplementary Table 25**). These findings indicate that IL-23 inhibition was associated with more sustained FB to KC signaling than IL-17 inhibition at week 16 of treatment.

The immune compartment showed a similar distinction in kinetics. Although T cells and DCs declined compositionally by week 16 under IL-23 inhibition, their transcriptional responses were comparatively modest compared to baseline. Both treatments initially suppressed *COTL1* in T cells at week 2, whereas IL-17 inhibition produced broader transcriptional deactivation by week 16, including reduced expression of *CD3E*, *LTB*, *TRBC2*, *CORO1A*, and *LCP1* (logFC range −0.46 to −0.71) (**Supplementary Table 10 and 14**). These genes showed similar but non-significant trends under IL-23i, indicating that the remaining T cells retained greater transcriptional activity despite their reduced abundance (**Supplementary Fig. 4N**). In DCs, *CCL19* was similarly suppressed at week 2 under both treatments (logFC=−0.64), but declined further by week 16 under IL-17 inhibition (logFC =−1.22) without significant additional suppression under IL-23 inhibition (logFC = −0.54) (**Supplementary Table 10 and 21, Supplementary Fig. 4O**). Thus, IL-23 inhibition produced substantial cellular and stromal remodeling while leaving a comparatively active residual immune transcriptional response.

These findings identify SPRR2F⁺ KCs as the principal cellular point of divergence in tissue resolution between IL-17-targeting biologics and IL-23i. Most KC clusters ultimately converged towards a common transcriptional resolution profile, whereas SPRR2F⁺ KCs remained incompletely resolved under IL-23i, retaining the cornified-envelope, keratinization, and inflammatory gene-sets that were rapidly suppressed by IL-17 inhibition. This differential resolution was accompanied by distinct temporal remodeling of KC communication and a more pronounced FB recovery transcriptional state under IL-23i, defining distinct trajectories of tissue remodeling that parallel the divergent kinetics of clinical improvement.

### SPRR2F⁺ KCs occupy the upper stratum granulosum, co-express AMPs and change with disease activity

To further investigate the distinct resolution kinetics of SPRR2F⁺ KCs following IL-17 versus IL-23 inhibition, we examined their protein expression, spatial localization in psoriatic skin, co-expression of multiple antimicrobial peptides (AMPs), and dynamics during treatment. We performed immunofluorescence (IF) on lesional and paired non-lesional skin from patients with psoriasis at baseline (**Supplementary Table 1**).

Co-staining with markers of epidermal differentiation localized SPRR2F⁺ KCs to the upper epidermis in psoriasis-affected skin (baseline). Loricrin, restricted to the upper stratum granulosum at the interface with the stratum corneum, showed near-complete overlap with SPRR2F⁺ cells, whereas TGM3 and involucrin, which span the stratum spinosum and stratum granulosum, overlapped with SPRR2F only in their most superficial layers. The co-localization of SPRR2F with TGM3 in these superficial layers is consistent with a more advanced stage of differentiation. SPRR2F was not detected in the stratum basale or lower stratum spinosum, localizing SPRR2F⁺ KCs to the upper stratum granulosum at the transition to the stratum corneum (**Figure 5A**). This localization persisted while the epidermis remained histologically inflamed, including at week 2 under IL-23i (**Figure 5B**). Interestingly, SPRR2F⁺ and KRT16⁺ KCs exhibited distinct spatial and temporal dynamics during treatment. In contrast to SPRR2F⁺ KCs, KRT16⁺ KCs remained detectable in treated skin through week 16 under IL-23i but progressively shifted from suprabasal towards basal layers, a change already evident at week 2. Consistent with these distinct localization patterns, SPRR2F and KRT16 were also transcriptionally separated into two distinct clusters in the HD-SPT dataset.

**Figure 5.**
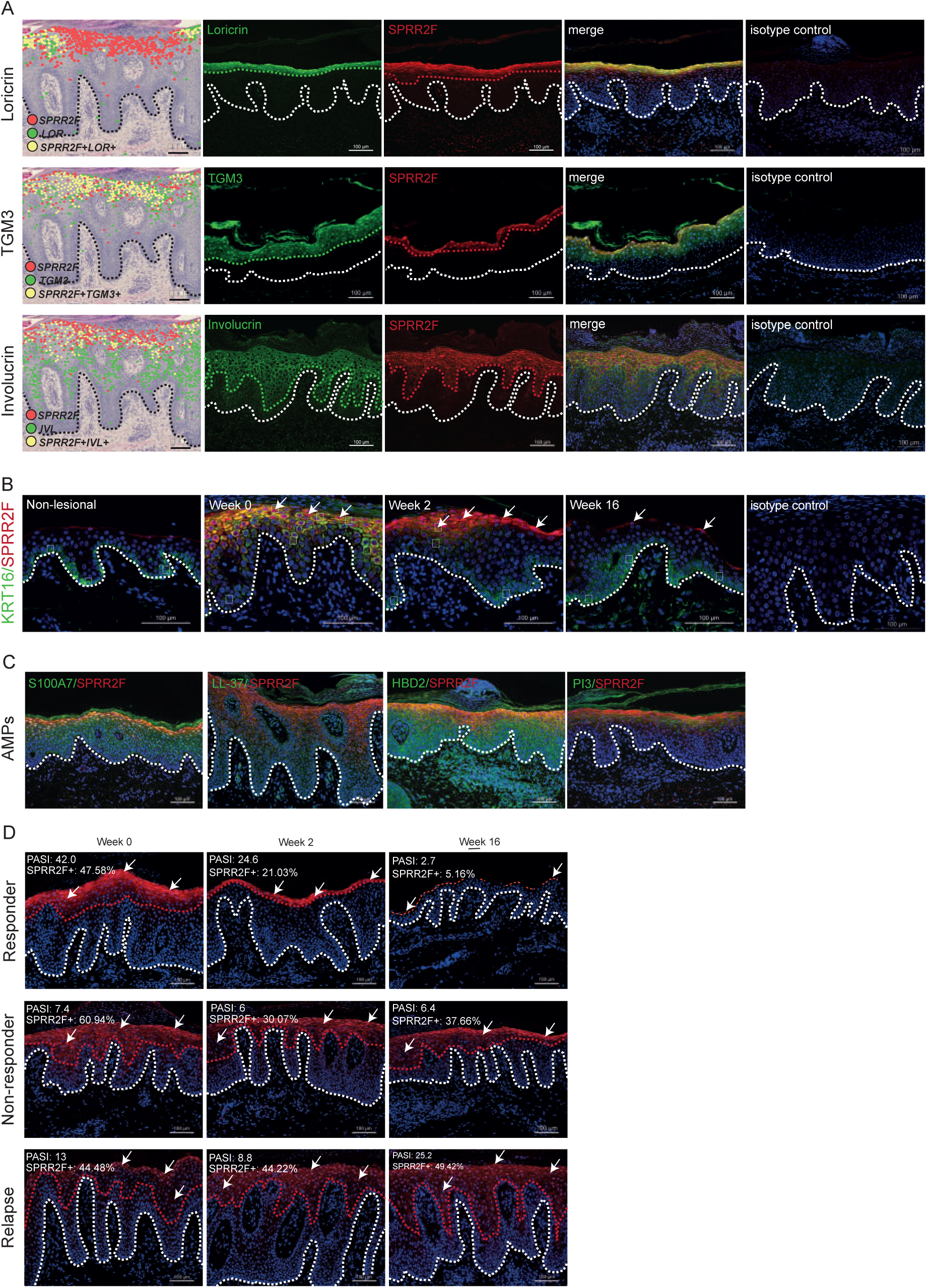
SPRR2F⁺ keratinocytes localize to the upper epidermis and associate with inflammatory activity and treatment response. (A) Spatial and protein-level localization of SPRR2F relative to markers of terminal epidermal differentiation in lesional skin. Rows: loricrin (LOR, top), transglutaminase 3 (TGM3, middle) and involucrin (IVL, bottom). Left: segmented cellular profiles overlaid on H&E, with red denoting profiles expressing *SPRR2F*, green the differentiation marker, and yellow profiles expressing both. Right: immunofluorescence for the differentiation marker (green), SPRR2F (red) and the merged channels with DAPI nuclear counterstain (blue). Dashed lines mark the dermal–epidermal junction (white or black) and the upper boundary of the marker-positive layer (colored). Scale bars, 100 µm. (B) Immunofluorescence for KRT16 (green) and SPRR2F (red) with DAPI nuclear counterstain (blue) in non-lesional skin and in lesional skin at baseline (week 0), week 2 and week 16. Dashed white line marks the dermal–epidermal junction. Scale bars, 100 µm. (C) Co-immunofluorescence of SPRR2F (red) with psoriasis-associated AMPs (green) in lesional skin: psoriasin (S100A7), LL-37 (CAMP), human β-defensin 2 (HBD2) and elafin (PI3), with DAPI nuclear counterstain (blue). Dashed white line marks the dermal–epidermal junction. Scale bars, 100 µm. (D) Immunofluorescence for SPRR2F (red) with DAPI nuclear counterstain (blue) in lesional skin from representative responder, non-responder, and relapse patients at baseline (week 0), week 2 and week 16 (left to right). Dashed white line marks the dermal–epidermal junction, dashed red line marks the upper boundary of the SPRR2F+ layer. Scale bars, 100 µm.

To determine whether the antimicrobial peptides identified in SPRR2F⁺ KCs by HD-SPT were also expressed at the protein level in lesional skin at baseline, we performed immunofluorescence analysis. SPRR2F⁺ KCs co-expressed multiple psoriasis-associated antimicrobial peptides, including psoriasin (S100A7), LL-37 (CAMP), human β-defensin 2 (HBD2), and elafin (PI3) (**Figure 5C**), providing independent protein-level validation of coordinated antimicrobial and inflammatory barrier activation in these KCs.

We related SPRR2F⁺ KC abundance to clinical course in serial, paired lesional biopsies (weeks 0, 2 and 16) from three patients with contrasting outcomes (**Figure 5D; Supplementary Table 1**). Clinical response was graded by PASI change from baseline. One patient treated with ixekizumab reached a PASI90 response (PASI 42.0, 24.6 and 2.7; −94%). A second ixekizumab-treated patient did not respond (PASI 7.4, 6.0 and 6.4; −14%, below PASI50). Because both received the same drug, differences in tissue between them are unlikely to reflect treatment class. The third patient relapsed under anti-TNF-α therapy (certolizumab pegol), offering a rare opportunity to examine SPRR2F⁺ KCs at clinical flare independently of the IL-17/IL-23 axis. In this patient, PASI fell from 13.0 to 8.8 at week 2 and then rose to 25.2 at week 16, 94% above baseline.

SPRR2F⁺ area tracked these trajectories. In the PASI90 responder it fell from 47.6% to 21.0% and then 5.2% (−89%), with histological resolution of epidermal hyperplasia. In the non-responder it fell to 30.1% at week 2 but rebounded to 37.7% at week 16 (−38%), about seven-fold above the responder. In the relapsing patient it never resolved (44.5%, 44.2% and 49.4%; +11%). Notably, it remained at baseline levels at week 2, despite the transient clinical improvement, and was still high at the week-16 flare. Thus, in all three patients, the direction of change in SPRR2F⁺ area matched the direction of change in PASI. Although limited to one patient per trajectory, this proof-of-concept series indicates that SPRR2F⁺ KC persistence accompanies persistent or recurrent disease, and motivates validation in larger, prospectively sampled cohorts.

In summary, these findings localize SPRR2F⁺ KCs to the upper stratum granulosum, where they co-express AMPs and markers of terminal differentiation. Their spatial dynamics distinct from those of KRT16⁺ KCs, their differential responses to IL-17 and IL-23 pathway inhibition, and their tracking of clinical trajectory during treatment suggest that SPRR2F⁺ KCs change dynamically with disease activity rather than representing a fixed feature of psoriatic skin.

### SPRR2F amplifies cytokine-driven inflammatory responses in KCs

To determine whether SPRR2F contributes functionally to the inflammatory response induced by psoriasis-associated cytokines, we first assessed whether cytokine stimulation of P-HEK recapitulated the transcriptional signature identified in lesional skin. P-HEK cells were stimulated with IL-17A, IL-23, and TNF-α (50 ng/mL each) and profiled by bulk RNA sequencing

Cytokine stimulation induced a lesional-like inflammatory transcriptome observed in lesional KCs (**Supplementary Table 26**), with cytokine-responsive genes consistently mirroring the lesional-versus-non-lesional expression patterns across all KC clusters (Spearman ρ = 0.11–0.33, all P < 10⁻⁴, **Supplementary Table 27, Figure 6A**), including concordant directional changes far exceeding those expected by chance (binomial padj =5.8 × 10⁻²⁶). This concordance was particularly evident in upper KC layers, including CKCs and SPRR2F⁺ KCs, where the lesional transcriptional profile was mirrored in P-HEK cells following cytokine stimulation. The stimulated P-HEK cells reproduced key features of this lesional signature, including induction of S100 alarmins (*S100A7A, S100A8, S100A9, S100A12*), multiple SPRR2 paralogs (*SPRR2A, SPRR2B, SPRR2D, SPRR2E, SPRR2G*), and antimicrobial and IL-36-axis molecules such as *LCN2, IL36G,* and *PI3* (ρ = 0.16, padj = 5.5 × 10⁻⁶; binomial P = 9.1 × 10⁻⁸; **Figure 6B, Supplementary Table 28**). Consistent with this transcriptional overlap, cytokine stimulation recapitulated the core inflammatory architecture of lesional SPRR2F+ KCs, with coordinated activation of TNFα- and IL-17-associated signaling, cornified-envelope remodeling, and innate antimicrobial responses (FDR = 3.7×10⁻⁸, 5.6×10⁻⁸, and 5.6×10⁻⁵, respectively, **Supplementary Table 29**). These findings indicate that cytokine stimulation of P-HEK cells captured the core alarmin–SPRR–antimicrobial response of lesional CKCs and SPRR2F+ KCs.

**Figure 6.**
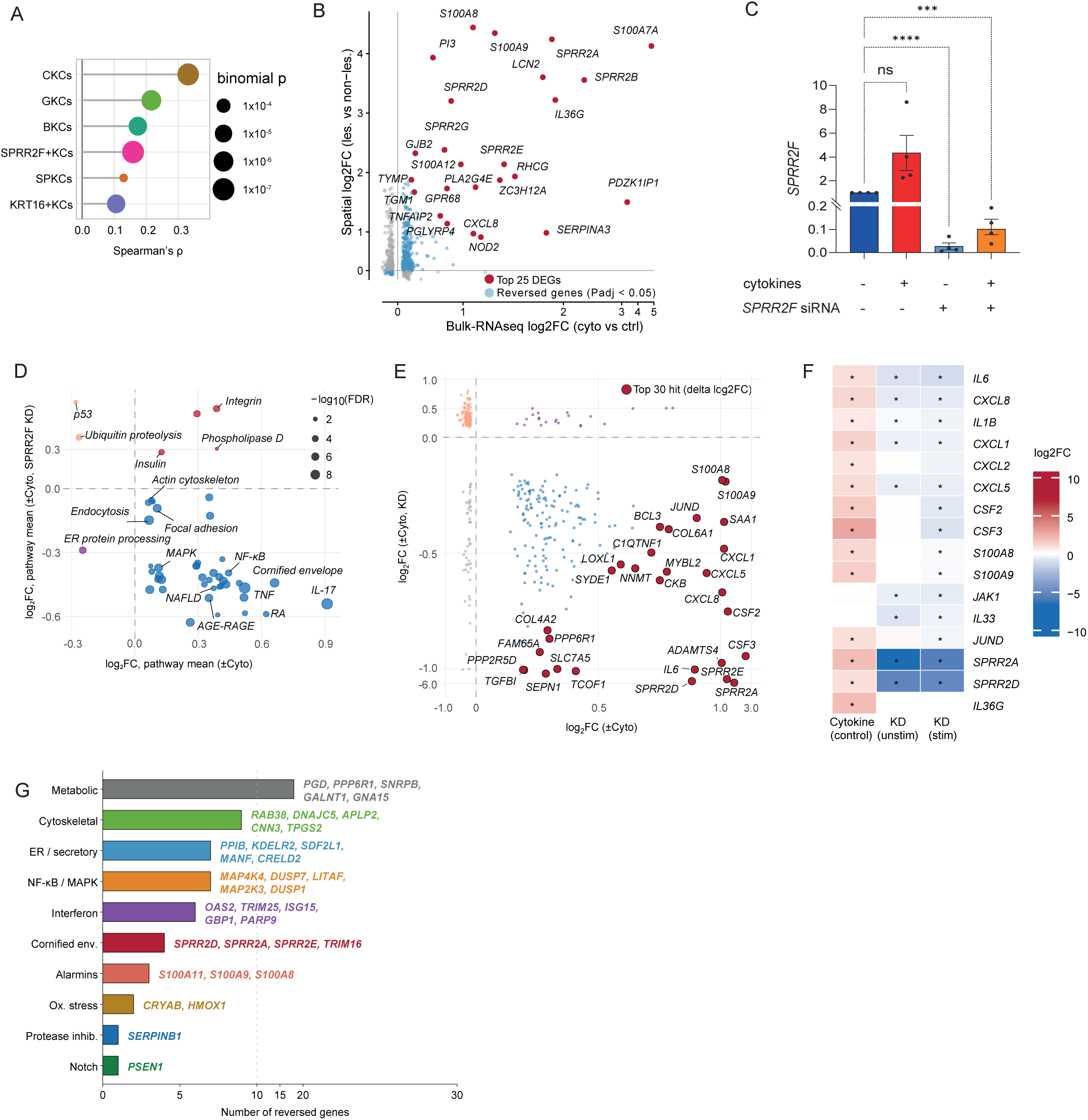
*SPRR2F* knockdown attenuates inflammatory responses to psoriasis-associated cytokines in patient-derived primary KCs. (A) Concordance between the cytokine-stimulated bulk P-HEK transcriptional response and the spatial lesional-versus-non-lesional signature, by spatial KC cluster. The *x* axis shows the Spearman correlation coefficient; dot size indicates the binomial *P* value for directional concordance; dot color denotes cluster as in Fig. 1. (B) Bulk log₂ fold change for cytokine stimulation versus control (Silencer Select siRNA negative-treated) P-HEK cells plotted against the spatial log₂ fold change for lesional versus non-lesional SPRR2F⁺ KCs. Red points denote the 25 most strongly changed genes and are labelled; light blue points denote concordant genes at adjusted *P* < 0.05; grey points denote all remaining genes tested. (C) *SPRR2F* expression measured by qPCR in P-HEK cells treated with control or SPRR2F-targeting siRNA, with and without cytokine stimulation (IL-17A, IL-23 and TNF-α). Bars show mean ± SEM; each point is one independent experiment (*n* = 4). Note the split *y* axis. \*\*\**P* < 0.001, \*\*\*\**P* < 0.0001 (repeated measures one-way ANOVA with the Geisser-Greenhouse correction, followed by Holm-Šidák’s multiple comparisons test). (D) Pathway-level mean log₂ fold change in response to cytokine stimulation in control P-HEK cells treated with Silencer Select negative-control siRNA (*x*-axis) plotted against the corresponding response following *SPRR2F* knockdown (*y*-axis). Dot size represents −log₁₀(FDR). Among cytokine-stimulated cells, blue denotes pathways whose cytokine-induced response is attenuated by *SPRR2F* knockdown, whereas red denotes pathways that are not attenuated. Among unstimulated cells, purple denotes pathways induced by *SPRR2F* knockdown, whereas orange denotes pathways that are not induced. (E) Log₂ fold change in response to cytokine stimulation in control P-HEK cells treated with Silencer Select negative-control siRNA (*x*-axis) plotted against the corresponding log₂ fold change in cytokine-stimulated P-HEK cells following *SPRR2F* knockdown (*y*-axis). Dark red points denote the 30 genes exhibiting the largest differences between the two responses. These genes, which are also labelled, represent genes induced by cytokine stimulation whose expression is suppressed following *SPRR2F* knockdown. Among cytokine-stimulated cells, blue points denote genes whose cytokine-induced expression is suppressed by *SPRR2F* knockdown, whereas purple points denote genes induced by *SPRR2F* knockdown. Among unstimulated cells, grey points denote genes whose expression is suppressed by *SPRR2F* knockdown, whereas orange points denote genes induced by *SPRR2F* knockdown. (F) Log₂ fold changes in selected inflammatory, alarmin, and cornified-envelope genes across three experimental contrasts in P-HEK cells: cytokine-stimulated control cells treated with Silencer Select negative-control siRNA, unstimulated P-HEK cells subjected to *SPRR2F* knockdown, and cytokine-stimulated P-HEK cells subjected to *SPRR2F* knockdown. Colors indicate the magnitude and direction of differential gene expression, ranging from −10 (blue) to +10 (red). Asterisks denote genes with significant differential expression after false discovery rate (FDR) correction (adjusted *P* < 0.05). (G) Functional categories of the genes are reversed towards the non-lesional state by *SPRR2F* knockdown under cytokine stimulation. Bar length indicates the number of genes per category; representative genes are listed beside each bar. The dotted vertical line marks 10 genes.

Given that cytokine-stimulated P-HEK cells recapitulate the core alarmin–SPRR–AMP module of SPRR2F+ KCs, we next determined whether SPRR2F regulates this response. To address this, we performed siRNA-mediated *SPRR2F* knockdown in P-HEK cells before stimulation with IL-17A, IL-23, and TNF-α, followed by bulk RNA-seq. Both unstimulated and cytokine-stimulated P-HEK cells in cell culture were treated with either Silencer Select negative control or with Silencer Select siRNA targeting *SPRR2F*, enabling us to distinguish the effects of *SPRR2F* depletion at baseline from those specifically emerging during cytokine-induced activation.

Efficient *SPRR2F* knockdown was confirmed by qPCR, demonstrating a >95% reduction in *SPRR2F* expression in cytokine-stimulated knockdown cells relative to cytokine-stimulated controls (**Figure 6C**). The specificity of the siRNA sequence was further confirmed by BLAST analysis against the human RefSeq RNA database, which identified 100% identity exclusively with two annotated *SPRR2F* transcript isoforms and no full-length matches to *SPRR2A, SPRR2D,* or *SPRR2E* (**see Methods**).

At baseline, *SPRR2F* knockdown altered 1,614 genes, including 701 upregulated and 913 downregulated genes (padj < 0.05) (**Supplementary Table 30**). Knockdown induced Notch pathway components including *JAG1, JAG2,* and *NOTCH1* and modestly reduced inflammatory mediators including *IL6, CXCL8,* and *IL1B* (**Supplementary Table 30**). Importantly, 76 of 157 genes shared between the knockdown response and the SPRR2F⁺ KC lesional signature were shifted towards the non-lesional profile (padj < 0.05, lfsr < 0.05), spanning cornified-envelope and differentiation, NF-κB and interferon-response, and Notch-processing related genes (**Figure 6D, Supplementary Table 31**). Thus, *SPRR2F* knockdown altered components of the lesional transcriptional profile even in the absence of exogenous cytokine stimulation.

The effect of *SPRR2F* knockdown was more pronounced following cytokine stimulation. Compared with stimulated control P-HEK cells, *SPRR2F* knockdown suppressed 621 genes and induced 411 genes (padj < 0.05) (**Supplementary Table 32**). The most strongly suppressed transcripts included the SPRR2 paralogs *SPRR2A, SPRR2D,* and *SPRR2E* (log2FC = −5.73, −5.21, and −4.51, respectively), together with *TGFBI* (−1.35) and *IL6* (−1.19). Other suppressed inflammatory regulators, including *CXCL5, SAA1, BCL3,* and *JUND*, pointed to attenuation of a broader NF-κB/AP-1 inflammatory network rather than selective loss of individual alarmins. This broader effect was reflected at the pathway level, with suppression of MAPK, IL-17, TNF, and innate immune signaling, together with changes in cornified-envelope and senescence pathways (**Supplementary Table 33**).

Notably, 172 genes induced by cytokine stimulation in control KCs were significantly attenuated following *SPRR2F* knockdown (**Supplementary Table 34**). These included inflammatory mediators *CXCL8, IL1B, CXCL1, CSF2,* and *CSF3*, as well as the alarmins *S100A8* and *S100A9*. *JAK1* and *IL33* were also reduced following knockdown (**Figure 6E-F**). Comparing these knockdown-responsive genes with the SPRR2F⁺ KC lesional signature identified 58 genes that were shifted towards the non-lesional profile (padj < 0.05, lfsr < 0.05) (**Supplementary Table 35**). These included *S100A8* and *S100A9*, the same SPRR2 paralogs, *TRIM16*, NF-κB-associated genes (*MAP4K4, LITAF, TNFRSF1A*), interferon-response genes (*GBP1, TRIM25, OAS2, ISG15*), the Notch-processing enzyme *PSEN1*, *S100A11*, and the barrier-defense gene *SERPINB1*, together with genes involved in oxidative stress, secretion, cytoskeletal organization, and metabolism (**Supplementary Table 36, Figure 6G**).

In conclusion, these findings indicate that SPRR2F is not merely a marker of inflammatory KCs. Cytokine stimulation recapitulated the SPRR2F-associated lesional signature, whereas SPRR2F knockdown attenuated many of the same inflammatory, interferon-response, differentiation, and barrier-associated genes, including the S100A8/S100A9 alarmin axis (**Figure 6G**). These results identify SPRR2F as a positive regulator of IL-17 cytokine-driven inflammatory gene expression in KCs that may help to sustain inflammatory epidermal response.

## Discussion

Using longitudinal HD-SPT of skin biopsies derived from psoriatic patients during treatment with IL-17 and IL-23 inhibitors, we identify SPRR2F⁺ KCs as an upper-epidermal, IL-17/IL-36-associated KC population whose rapid decline parallels earlier clinical response to IL-17 inhibition and initially persists under IL-23 inhibition. Functionally, SPRR2F promotes cytokine-driven inflammatory gene expression in P-HEK cells. These findings support the epidermis as an active participant in psoriatic inflammation cascade and identify a KC population that distinguishes the tissue responses to two mechanistically distinct biologic classes.

The relationship between KCs and the immune system in plaque psoriasis has been debated for decades. Until the 1970s, psoriasis was regarded primarily as a disorder of KC dysfunction^27^, a view reflected in therapeutic approaches of the era that targeted the epidermis directly e.g. with retionids, including serial dermatome shaving^28^. Accumulating evidence for dysregulated T cell and DC activation subsequently led to its reframing as an immune-mediated disease driven by the IL-23/IL-17 axis^29^, in which KCs were viewed primarily as responders to IL-17A and IL-22 produced by the dermal infiltrate^30^. Evidence has since restored an active role for KCs. Gain-of-function *CARD14* mutations, which enhance NF-κB activation and psoriasis-associated gene expression in KCs, cause familial psoriasis^31^, and KC-derived LL-37 complexed with self-DNA activates plasmacytoid DCs to initiate inflammation^32^. KCs can also retain disease-associated properties independently of the inflammatory milieu, including a psoriatic gene expression profile in iPSC-derived KCs generated weeks after removal from lesional skin^33^, and transcriptional memory of prior inflammation through persistent chromatin accessibility at inflammatory loci in murine epidermal stem cells^34^. Activated KCs in turn produce cytokines, chemokines and AMPs that recruit and activate immune cells, establishing a bidirectional, self-amplifying circuit in which KCs are active participants^30,35^. However, which KCs sustain this circuit, where they reside within the lesion, and how they respond to targeted therapy in patients have remained uninvestigated.

Our high-definition, longitudinal spatial atlas addresses these questions by resolving, at near single-cell resolution and within the same patients across the treatment course, how this inflammatory network is spatially organized and which KCs sustain it. SPRR2F⁺ KCs were transcriptionally and spatially distinct from the established KRT16⁺ disease-associated KCs^36,37^, largely restricted to lesional skin and localized to the upper stratum granulosum. There, SPRR-family cornified envelope genes were co-expressed with S100A8/S100A9 alarmins, IL-36 cytokines, AMPs and NF-κB-dependent cytokines (**Figure 2**), all of which have been implicated in psoriatic inflammation^38,39^, placing SPRR2F⁺ KCs at the intersection of dysregulated cornification and immune amplification. Although several of these alarmin and AMPs were shared with KRT16⁺ KCs, the two differed in their cytokine-response profiles: KRT16⁺ and lower epidermal KCs displayed a broad interferon-associated response, whereas SPRR2F⁺ KCs carried a more focused IL-17/IL-36-associated gene signature. SPRR2F⁺ KCs also showed the highest intercellular communication prioritization within the lesional epidermis, driven by autocrine *S100A8/S100A9–CD36* signaling. SPRR2F knockdown in P-HEK attenuated inflammatory output both at baseline and under psoriasis-associated cytokine stimulation (IL-17A, IL-23 and TNF-α), indicating that SPRR2F contributes to, rather than merely marks, the KC inflammatory response, and suggesting that the cornification and immune-amplifying arms of this program are functionally linked rather than independent consequences of psoriatic inflammation. This coupling may reflect a broader principle of inflammatory epithelial biology, whereby structural differentiation and immune effector genes become co-regulated in disease-associated epithelia, as increasingly recognised across barrier tissues including the skin, gut and respiratory^36,40,41^. To our knowledge, however, such coupling has not previously been shown at this level of spatial and functional detail in plaque psoriasis. These findings support a view of the epidermis not only as the target of psoriatic inflammation but as an active component of its tissue-level organization and have a potential for future targeted treatment especially in milder diseases, where topical treatment might be sufficient.

A central and unresolved question in plaque psoriasis therapeutics has been why IL-17 and IL-23 pathway inhibitors, which intercept the same inflammatory axis at distinct pharmacological nodes, differ in their response kinetics: head-to-head trials have shown faster early PASI reductions with IL-17 inhibitors^42^, but comparable or superior PASI90 responses at one year with IL-23i^43^. This difference has been attributed to the upstream versus downstream position of their targets in the IL-23/Th17/IL-17 hierarchy^29,44^, and transcriptomic studies have shown that IL-17 inhibition normalizes KC inflammatory gene circuits earlier and more broadly than upstream cytokine inhibition^38,45^, consistent with KCs being the principal tissue target of IL-17^46,47^. Our spatially resolved treatment trajectories add cellular resolution to these observations. Under IL-17 inhibition, the alarmin–antimicrobial–IL-36 transcripts of SPRR2F⁺ KCs were early decreased by week 2, and 56.8% of their lesional transcriptional signature was reversed by week 16, compared with only 6.2% under IL-23 inhibition. Although no cell type changed significantly in abundance at week 2, SPRR2F⁺ KCs showed the strongest, albeit non-significant, trend towards reduction under IL-17 inhibition and had contracted significantly by week 16. The rapid resolution of SPRR2F⁺ KCs therefore parallels the kinetic advantage of IL-17 inhibition, consistent with their focused IL-17/IL-36-associated cascade at baseline.

In contrast, the response to IL-23 inhibition was not simply a delayed version of the IL-17 response but was qualitatively distinct. By week 16, BKCs, SPKCs, GKCs and KRT16⁺ KCs had converged towards a shared resolution signature under both treatments, whereas SPRR2F⁺ KCs remained largely unresolved under IL-23 inhibition, retaining enrichment of cornified-envelope and keratinization genes without significant reduction in JAK-STAT activity. SPRR2F⁺ KCs therefore represented the principal point of divergence between the two therapeutic trajectories. Outside the epidermis, IL-23 inhibition restored homeostatic FB genes, including *PI16,* MFAP4, PRELP and *ELN,* more strongly than IL-17 inhibition and, by week 16, was accompanied by FB expansion that was not observed under IL-17 inhibition. This is in line with single-cell evidence that FB and myeloid remodeling precede KC transcriptional changes under IL-23 inhibition (38291032) and suggests that IL-23 inhibition engages KCs through an indirect, stromal-first route and also indirectly through TH-17 cells.

The atlas also offers a temporal sequence of tissue remodeling during cytokine inhibition. By week 2, both treatment classes induced significant transcriptional responses without significant changes in overall cell-type composition. Instead, early changes were concentrated within the KC compartment, including inhibition of alarmin–antimicrobial–IL-36 transcripts in SPRR2F⁺ KCs, KRT16⁺ KCs and GKCs under IL-17 inhibition, and the suprabasal-to-basal shift of KRT16⁺ KCs under IL-23 inhibition. This suggests that the earliest phase of biologic therapy acts primarily through transcriptional reprogramming and spatial reorganization of existing KCs rather than through the elimination or recruitment of cell populations. These findings extend previous bulk transcriptomic studies, in which IL-17 inhibition produced broad transcriptome normalization by week 2, with early suppression of activated KC effector genes predicting clinical outcomes at week 12^38,45^, and in which molecular responses to IL-23p19 inhibitors were evident by week 4, before maximal clinical response^48^. Thus, the tissue appears to lose its inflammatory transcriptional identity before it loses its inflammatory cellular composition, a principle with emerging support in other immune-mediated inflammatory diseases treated with targeted biologics^49,50^.

The persistence of SPRR2F⁺ KCs in the patient who relapsed, despite transient clinical improvement (**Figure 5D**), raises the question of whether the epidermis itself contributes to inflammatory memory in plaque psoriasis. Our data raise the possibility that, once established, *SPRR2F* expression can be rapidly re-induced upon cytokine re-exposure, in line with evidence that epithelial cells retain transcriptional memory of prior inflammation^34,41^. Notably, SPRR2F⁺ KCs remained incompletely resolved at week 16 under IL-23i despite substantial clinical improvement, indicating that residual epidermal/dermal disease can persist beyond what is captured by clinical assessments such as PASI. Whether such residual SPRR2F⁺ KCs influence the durability of remission remains to be determined. Clinically, residual SPRR2F expression, detectable by IF in tissue biopsies, could indicate incomplete molecular remission, and SPRR2F⁺ KCs may represent a potential therapeutic target beyond upstream cytokine inhibition and contribute to better understanding of the remission and disease modification. Combined with non-invasive measurement of S100A8/S100A9 in skin rinsing samples, which mirrored the differential kinetics of IL-17 and IL-23 pathway inhibition from the earliest time point, such tissue and surface markers could complement PASI in treatment monitoring. These hypotheses and potential biomarker applications will require investigation in larger longitudinal cohorts with post-withdrawal follow-up.

Our study has several limitations. Although it is, to our knowledge, the largest longitudinal HD-SPT study of human skin to date (42 biopsies) and the only one directly comparing IL-17- and IL-23-targeted therapies in patients with moderate to severe psoriasis through week 16, it included only 11 individuals of European ancestry. In addition, the IL-17 arm combined IL-17A inhibition and IL-17RA blockade, limiting our ability to resolve drug-specific effects, although both produced a similar early trend towards reduction in SPRR2F⁺ KCs, supported by IF (**Supplementary Fig. 6**). A key technical limitation was proximal transcript contamination arising from imperfect bin-to-cell assignment in the dense and morphologically heterogeneous skin microenvironment, particularly in immune cells, in which FB-derived collagen transcripts and BKC-derived signals could be detected. Similar transcript misassignment has been reported in high-resolution spatial transcriptomic studies of other tissues, including the lung^51^. To minimize these effects, we used nucleus-based segmentation and restricted cell-level analyses to nucleus-associated transcripts, an approach previously applied to address this issue^51^. We further removed doublets using a curated scRNA-seq reference psoriatic skin atlas. Additionally, we cross-validated dermal transcriptional signals against it to distinguish genuine immune-cell expression from signals more consistent with neighboring FBs or KCs. Genes with discordant spatial and single-cell expression patterns were interpreted cautiously or excluded from population-level analyses. However, nucleus-based segmentation reduced transcript and UMI recovery, underscoring the need for improved skin-specific whole-cell segmentation and bin-to-cell assignment approaches (34650268). Finally, non-lesional and healing skin showed lower transcript and UMI counts, consistent with reduced transcriptional activity^2^; samples were therefore normalized individually before integration to improve comparability across disease states.

In brief, this study illustrates what becomes interpretable about inflammatory disease when tissue architecture, cellular identity and temporal dynamics are resolved together. Bulk transcriptomics identifies disease-associated genes but loses cellular and spatial context; dissociative single-cell approaches resolve cellular heterogeneity but lose the tissue architecture that gives spatial relationships, such as the proximity of SPRR2F⁺ KCs to the cornified surface, their biological meaning; and conventional spot-based spatial transcriptomics preserves tissue context but merges adjacent epidermal layers and neighboring niches. Combining Visium HD with longitudinal within-patient sampling and perturbation with two drug classes enabled the identification of spatially restricted KCs, the reconstruction of treatment trajectories under two mechanistically distinct interventions, and the functional interrogation of SPRR2F. Plaque psoriasis, with its accessible tissue, well-investigated immunopathology and remarkable therapeutic toolkit, is an ideal system for this kind of spatial functional genomics, and the framework established here provides a template for other chronic inflammatory diseases in which the tissue-level organization of the inflammatory state remains poorly understood.

## Materials and Methods

### Visium HD-SPT

#### Procurement of skin samples

The Visium HD spatial transcriptomics (HD-SPT) cohort included 11 patients with moderate-to-severe plaque psoriasis (mean PASI score of 17.8 ± 3.3 at baseline) treated with brodalumab (*n* = 3), ixekizumab (*n* = 3), risankizumab (*n* = 3), or guselkumab (*n* = 2). Detailed information on the patient cohort is outlined in **Supplementary Table 1**. Briefly, our cohort comprised 8 males (73%) and 3 females (27%) with a mean age of 44.3 ± 4.7 years and a mean BMI of 28.9 ± 0.6 kg/m² (mean ± SEM). Four mm punch skin biopsies were collected from lesional and non-lesional skin sites at baseline (week 0; lesional *n* = 11; non-lesional *n* = 11), at week 2 (*n* = 10), and at week 16 (*n* = 10) after initiation of treatment. Two of 44 biopsies were unavailable (one week 2 and one week 16 sample from two brodalumab-treated patients); all other patients completed longitudinal sampling. The study was approved by the local ethics committee of the University of Luebeck under the protocol “Clinical Demonstrator 1” (ethics approval 20-170; ethicPool follow-on number 2023-800), and sample analysis was approved under the protocol “Identification of Biomarker to predict treatment outcome and disease prevention using multiomic analysis in patients with chronic inflammatory barrier diseases” (ethics approval 22-133; ethicPool follow-on number 2025-237).

#### Skin samples processing

Freshly obtained skin biopsies were immediately placed in 4% (v/v) paraformaldehyde (PFA) solution and fixed for 48 to 72 hours at 4°C while rotating. After fixation, tissue was dehydrated using increasing ethanol concentrations (70%, 80%, and 96%) and then treated with xylol to displace the ethanol. The tissue was subsequently embedded in paraffin at 60°C and FFPE blocks were stored at RT until downstream processing. 5 µm-thick FFPE tissue sections were mounted onto SuperFrost slides (VWR International, Cat. No. 631-0108). For deparaffinisation, the slides were incubated at 60°C for 2 hours, immersed in xylene (Sigma-Aldrich, St. Louis, MO, USA, Cat. No. 214736), and rehydrated in decreasing ethanol concentrations (Sigma-Aldrich, St. Louis, MO, USA, Cat. No. E7023). Haematoxylin and eosin (H&E) staining was performed using Mayer’s haematoxylin solution (Millipore Sigma), Bluing reagent (Epredia, Kalamazoo, MI, USA; Cat. No. 6769001), and alcoholic eosin (Sigma-Aldrich, St. Louis, MO, USA, Cat. No. HT110116). High-resolution brightfield images of the stained slides were acquired using a Keyence BZ-X810 microscope (Keyence) with a 20× objective. Afterwards, the slides were placed into 10x Genomics cassettes and processed according to the manufacturer’s instructions. Briefly, slides were destained with 0.1 N HCl (Sigma-Aldrich, St. Louis, MO, USA, Cat. No. H9892) and washed with TE Buffer (Promega, Cat. No. V6231). The whole human transcriptome probe panel (10x Genomics), consisting of ∼3 specific probes for each targeted gene, was hybridized to mRNA molecules from the spatial slide after de-crosslinking. Probe pairs were ligated at their junctions, forming single-stranded ligation products. Before probe release and capture, Visium HD slides (10x Genomics) were thawed, prepared by washing with 0.1XSSC Buffer (Millipore Sigma), and air-dried in the Visium CytAssist instrument (10x Genomics). Meanwhile, the tissue sections were stained with 10% (v/v) alcoholic eosin solution. In the Visium CytAssist, probes were treated with RNase, permeabilized, and captured by the poly(dT) regions of the pre-coated capture probes on the Visium HD slides. Ligation products were then extended by adding a Spatial Barcode, UMI, and partial Read 1 primer, generating spatially barcoded ligation products. These were subsequently released from the slides, pre-amplified, and purified with SPRISelect Reagent (Beckman Coulter GmbH, Krefeld, Germany; Cat. No. B23317).

#### Library construction and sequencing

The pre-amplified products were then used for further library construction. To prevent PCR saturation and over-amplification, the optimal sample index PCR cycle number was determined by qPCR of the pre-amplification products. To generate final library molecules, the amplified materials were then indexed via sample index PCR, cleaned with SPRISelect reagent, and assessed for quality and amplicon length using an Agilent Bioanalyzer High Sensitivity chip (Agilent Technologies, Cat. No. 5067-4626). Quantification of the obtained library was performed by qPCR with the NEBNext Library Quant Kit for Illumina (New England Biolabs, Ipswich, MA, USA, Cat. No. E7630S) using a StepOne™ Real-Time PCR System (Thermo Fisher). Visium HD spatial gene expression FFPE libraries consisted of Illumina paired-end sequences flanked with P5 and P7 adaptors, with a 43 bp unique molecular identifier (UMI) and Spatial Barcode encoded in TruSeq Read 1. Small RNA Read 2 (Read 2S) was used to sequence the ligated probe products. Sequencing was performed using 150 bp paired-end reads on an Illumina NovaSeq X-10 (Illumina) by the commercial vendor BMKgene GmbH, targeting a minimum of 275 million read pairs per fully covered area as recommended for FFPE tissue. Prior to sequencing, quality control of the library was conducted using an Agilent TapeStation System (Agilent Technologies).

### Building a reference scRNA-seq atlas of psoriatic skin

To integrate our Visium HDST data with scRNA-seq data, we analyzed publicly available scRNA-seq datasets as outlined in **Supplementary Table 37** from the Gene Expression Omnibus (GEO)^52^ database including GSE151177, GSE162183, GSE228421, and GSE183047^19–23^,. In total, our scRNA-seq object contained data from 49 patients with plaque psoriasis across 38 lesional and 17 non-lesional skin sites. In addition, 17 skin samples from healthy controls were also included. Raw gene expression matrices were processed using Seurat (v.5)^53^ with quality control steps, including MiQC^54^ filtering and doublet detection with scDblFinder^55^. Cells were filtered based on UMI count (≥ 1000 and < 30000), gene count (≥500 and ≤5000), mitochondrial gene percentage (≤ 10%), and Log10 genes per UMI (≥ 0.8). To exclude low-quality samples, we filtered out samples with < 500 cells. Integration was performed using the JoinLayers function, followed by SCTransform (SCT) normalisation^56^. Principal component analysis (PCA) on 100 PCs was applied to variable genes, with batch correction using Harmony. Graph-based clustering was performed using FindClusters (first 25 PCs) and FindClusters (resolutions 0.1–0.8), while uniform manifold approximation and projection (UMAP) dimensionality reduction was performed using the first 25 Harmony-reduced dimensions^57^. A total of 177,006 cells (91,953 cells from psoriatic lesional skin, 63,281 cells from psoriatic non-lesional skin, and 21,772 cells from healthy controls) passed stringent quality control, detecting 15,455 genes across all samples. Cluster identities were assigned at resolution 0.6, yielding 24 clusters. Differential gene expression analysis was performed with PrepSCTFindMarkers and FindAllMarkers. To annotate the clusters, we used the top differentially expressed gene from each cluster along with the expression of known markers identified in previous scRNA-seq studies. Further subclustering of cluster 4, cluster 10, cluster 13, and cluster 18 with the Louvain algorithm and UMAP dimensionality reduction identified a total of 33 distinct cell clusters, including basal KCs (BKCs_I, BKCs_II, BKCs_III, and BKCs_IV), spinous KCs (SKCs_I and SKCs_II), granular KCs (GKCs), secretory papillary FBs (SPFs), secretory reticular FBs (SRFs), pro-inflammatory FBs (PIFs), mesenchymal FBs (MSFs), pericytes (Peri), lymphatic ECs (LECs) and vascular ECs (VECs), pro-inflammatory macrophages (PIMs), anti-inflammatory macrophages (AIMs), monocytes (Mono), conventional DCs type 1 (cDC1), conventional DCs type 2 (cDC2), Langerhans cells (LCs), B cells (B_cells), mast cells (Mast), CD8+ regulatory T cells (Tregs_CD8), CD8+ effector T cells (Teff_CD8), CD8+ memory T cells (Tmem_CD8), CD4+ regulatory T cells (Tregs_CD4), melanocytes (Melan), sebaceous glands (SG), and five mixed clusters (MIX1, MIX2, MIX3, MIX4, and MIX5). A full list of the identified clusters and the respective cellular markers used for cell annotation is presented in **Supplementary Table 38**. The mixed clusters were removed from the object for downstream processing. The B cell cluster (B_cells) did not pass downstream quality control and was also removed from the final object (**Supplementary Fig. 1A**).

### Spatial data processing and segmentation

The sequencing data were pre-processed using Space Ranger (v3.0) software^16^. The Space Ranger count pipeline aligned FASTQ files to the GRCh38 human reference genome, assigned spatial Unique Molecular Identifiers (UMIs) and barcodes, and generated gene expression matrices mapped to the microscopic tissue images. The 2-µm square-bin (Visium HD) resolution was processed through the ENACT^58^ pipeline (sparse count matrix, gene and barcode tables, and the associated histology image). Within the pipeline (configuration “ENACT_Naive”), each sample’s whole-slide histology image was loaded and normalized, and nuclear/cell segmentation was performed with StarDist^59^ on image patches of 16,000 pixels. Segmented cells were represented as polygons, and the 2-µm Space Ranger bins were likewise represented as polygons and assigned to segmented cells by spatial join by bin-to-cell assignment method. Counts from all bins assigned to a given cell were aggregated to build a per-sample cell-by-gene count matrix. This cell-segmented count matrix constituted the per-sample output subsequently loaded into R, with cell-type calls for all downstream analyses in this manuscript generated in the R programming language.

### Spatial object construction and quality control

Each sample’s segmented count matrix and image were loaded into a Seurat^53^ object, and treatment, patient identifier, study week, and lesional/non-lesional site were assigned from a per-sample datasheet. Quality control was then applied per sample, with mitochondrial content and hemoglobin content computed per bin, and log10 genes-per-UMI. Low-quality bins were removed using Seurat’s implementation of MiQC^54^. This was followed by hard thresholds retaining bins with mitochondrial percentage < 10%, hemoglobin percentage = 0%, and log10 genes-per-UMI between 0.85 and 0.995. Each quality-filtered sample was then deconvolved against the single-cell reference with RCTD^60^ to assign primary/secondary cell-type calls per bin, which were retained as bin-level metadata; bins were excluded if they were “doublet_certain”.

Per-sample objects were merged and re-normalized with SCTransform^56^ after re-splitting by sample to preserve per-sample sequencing-depth models. Variable features were defined from cluster markers (FindAllMarkers, adjusted *P* < 0.01), and the merged object was scaled, PCA-reduced, and integrated across samples with Harmony^57^ (Seurat IntegrateLayers, SCT-normalized layer). Shared-nearest-neighbor graphs were built on 30 Harmony dimensions, clustering was run across a resolution and cells were embedded with UMAP (30 Harmony dimensions), with a resolution of 0.6 selected. Following clustering, cell-type labels were refined through an extensive iterative process in which individual clusters were repeatedly re-clustered at cluster-specific resolutions on the shared-nearest-neighbor graph and relabeled by marker expression, with clearly mixed or low-quality sub-clusters removed at each step. This resolved, among others, T-cell, dendritic cell/macrophage-monocyte, endothelial/pericyte/adipocyte, smooth-muscle, and multiple dermal FB subpopulations, as well as KC clusters spanning basal (BKCs), cornified (CKCs), granular (GKCs), spinous (SPKCs), KRT16+, and SPRR2F+KCs. Final cluster identity was confirmed by differential marker expression (FindAllMarkers, log2 fold-change threshold 0.25, positive markers only). Sample identifiers were harmonized across acquisition batches, and the resulting annotated object was used for all downstream analyses below.

### Pseudobulk differential expression analysis

For each pairwise comparison of interest within each treatment axis—baseline versus week 2, baseline versus week 16, and, separately, lesional versus non-lesional skin at baseline—cell-type pseudobulk profiles were built with muscat^61^ (summed raw counts by cluster and sample) and tested with a patient-adjusted linear model using limma-voom via muscat’s pbDS, with a minimum threshold of 10 cells per pseudobulk sample. We considered complete paired data for this analysis, with one patient excluded from the IL-17 treatment group at week 2 and week 16.

Percent-of-cells-expressing frequencies were computed per gene and cluster and expressed relative to each gene’s mean expression frequency across all clusters, to flag cluster-enriched genes. For the IL-17-axis and IL-23-axis treatment comparisons, per-cluster effect sizes were jointly shrunk across both treatment axes using multivariate adaptive shrinkage (mashr)^62^, with standard errors derived from the limma-voom^63^ *t*-statistics. Canonical covariance matrices were used throughout, and data-driven covariances were added when at least 50 gene-cluster pairs reached nominal significance (mash_1by1, threshold 0.05). This produced posterior-mean shrunk log-fold changes and local false sign rates (lfsr) per gene, cluster, and treatment axis. Genes were classified per cluster/comparison as IL-17-specific, IL-23-specific, shared (“both”), or neither, using an lfsr threshold of 0.05 in each axis. The baseline lesional-versus-non-lesional comparison was shrunk analogously with mash using canonical covariances only. An lfsr < 0.05 was used to define differentially expressed genes for downstream reporting, with shrunk effect sizes driving gene classification or filtering in later analyses.

### Cell-type and tissue-compartment compositional analysis

Cell-type proportions per sample were estimated with the speckle R package^64^ (logit-transformed proportions), computed separately within an epidermal compartment (basal, cornified, granular, spinous, KRT16+, and SPRR2F+ KCs) and a dermal/immune compartment (FBs, endothelial cells, macrophages/monocytes, dendritic cells, T cells). Hair follicle, sebaceous gland, and sweat gland clusters were excluded as anatomically restricted to specific tissue regions. Within-patient longitudinal changes in lesional skin were tested per treatment axis and comparison (weeks 0–2, 0–16, and 2–16) with patient-adjusted linear models (empirical Bayes moderated *t*-statistics, robust variance shrinkage) on the logit-transformed proportions, testing the study-week coefficient. Baseline lesional-versus-non-lesional compositional differences were tested per compartment with an analogous patient-paired model, testing the site coefficient.

### Cell-cell communication analysis

Ligand-receptor communication was inferred with MultiNicheNet^26^ (human ligand-target and ligand-receptor prior models), run separately for the baseline lesional-versus-non-lesional comparison and for each treatment-axis/time point contrast, with patient modelled as a covariate (minimum 5 cells per group; log2 fold-change threshold 0.1; differential-expression *P*-value threshold 0.1, uncorrected; minimum expressed fraction 0.01; minimum sample proportion 0.1; differential expression via edgeR^65^). Genes were pre-filtered, per contrast, to those either differentially expressed (lfsr < 0.1 in the corresponding shrunk effect-size table) or present in the ligand-receptor network. Prioritized interactions were further restricted to those enriched in both the sending and receiving cell type relative to global expression frequency, then passed through a curated quality filter removing interactions flagged by cross-lineage contamination marker lists curated per cell type. Interactions between cell-type pairs considered spatially implausible under a curated tissue-adjacency model or any interaction involving cornified KCs, which are terminally differentiated and considered incapable of active signaling, and receptor calls considered database artefacts or cell-type-inappropriate. A curated panel of specific interactions of interest spanning alarmin (S100A8/A9), EGFR-family, IL-36-family, connexin, and additional chemokine/complement/adhesion signaling was assembled across all time points and treatment axes for combined presentation.

### Pathway, cytokine activity inference and GSEA

Transcription-factor-independent pathway activity was inferred with a multivariate linear model against the PROGENy^24^ human network (top 500 target genes per pathway), and cytokine-signaling activity was inferred with a univariate linear model against the CytoSig^25^ cytokine-response signature matrix (decoupleR^66^). Both were applied per cluster to the same contamination-filtered, shrunk log-fold-change vectors used for gene set enrichment (minimum 5 overlapping genes per source), for the baseline comparison and every treatment-axis/time point contrast. *P*-values were adjusted (Benjamini-Hochberg) within each analysis-cluster-network combination. Per-spot PROGENy pathway scores and CytoSig cytokine-activity scores were additionally computed directly, as a matrix product of normalized expression against the same PROGENy/CytoSig weight matrices used above, for export to an external spatial-data viewer alongside spot coordinates, histology images, and cluster color assignments.

Pre-ranked gene set enrichment analysis (fgsea^67^) was run per cell-type cluster and comparison, using shrunk log-fold changes as the ranking statistic (minimum gene-set size 15, maximum 500), against five MSigDB^68^ (Homo sapiens) collections: Hallmark, KEGG^69^, WikiPathways^70^, and Reactome^71^. Before ranking, genes matching a curated, per-cell-type cross-lineage contamination marker list were removed from each cluster’s ranked list. Clusters retaining fewer than 50 genes after this filter were excluded from that comparison. *P*-values were adjusted for multiple testing (Benjamini-Hochberg) within each MSigDB categories.

### S100A8/S100A9 quantification in skin rinsing samples

#### Procurement of skin rinsing samples

Skin rinsing samples from 72 patients with moderate-to-severe plaque psoriasis with a mean baseline PASI score of 14.3 ± 1.0 were procured. The patients were treated with brodalumab (*n* = 21), ixekizumab (*n* = 20), guselkumab (*n* = 11), and risankizumab (*n* = 20), and their demographics, as well as clinical scores over time, are summarized in Supplementary Table 7. The study cohort consisted of 49 males (68%) and 23 females (32%), with a mean age of 47.8 ± 1.6 years and a mean BMI of 31.1 ± 0.7. One patient (10151) was excluded from analysis, as the baseline (week 0) lesional skin-rinsing sample, used for correlation with PASI, was unavailable. Skin rinsing samples were obtained from matched lesional and non-lesional skin areas at baseline (lesional *n* = 72, non-lesional *n* = 72), at week 2 (lesional *n* = 67, non-lesional *n* = 65), and at week 16 (lesional *n* = 67, non-lesional *n* = 69) after initiation of treatment. Individual samples that were not obtained or did not yield a measurable value were treated as missing; the affected patients were retained and all available samples were included in the analysis. All values are expressed as mean ± SEM. The study was approved by the local ethics committee of the University of Luebeck under the protocol “Clinical Demonstrator 1” (ethics approval 20-170; ethicPool follow-on number 2023-800), and sample analysis was approved under the protocol “Identification of Biomarker to predict treatment outcome and disease prevention using multiomic analysis in patients with chronic inflammatory barrier diseases” (ethics approval 22-133; ethicPool follow-on number 2025-237).

To quantify the concentration of S100A8/S100A9 in skin rinsing samples obtained from lesional and non-lesional skin samples of patients with plaque psoriasis, a non-invasive collection method for skin rinsing samples was used. A cylindrical guide made from a polypropylene tube (Sarstedt, Nümbrecht) was placed on the targeted skin area, and 1 mL of rinsing solution containing PBS and 0.1% (v/v) Triton X-100 was introduced into the guide using a pipette. The solution was gently agitated in a circular motion to ensure thorough rinsing of the skin surface for 30 seconds, then transferred using a pipette to a clean 1.5 mL Eppendorf tube (Sarstedt). The skin rinsing sample was centrifuged at 10,000 × *g* at RT for 10 min, and the supernatant was recovered into a new clean collection tube. To this solution, 95 µL of storage buffer containing PBS supplemented with 0.1% (v/v) Triton X-100 and 10% (w/v) BSA was added. After thorough mixing and a brief centrifugation, the supernatant was aliquoted into five collection tubes with a total volume of 200 µL and stored at −80 °C until further analysis.

#### S100A8/S100A9 ELISA measurements

The concentration of S100A8/S100A9 in the skin rinsing sample was measured using a commercially available standard sandwich ELISA kit (R&D Systems) with a measurable concentration range of 93.75 to 6000 pg/mL.To ensure detection within this range, samples were diluted at a range of 1:10–1:2000 (v/v). The plates were read at 450 nm using the GloMax Discover System (Promega).

### Immunofluorescence of FFPE-skin sections

FFPE skin biopsies for IF analysis were obtained from plaque psoriasis patients enrolled in the “Clinical Demonstrator 1” study, under the ethics approval described above (see *Visium HD spatial transcriptomics*). For longitudinal analysis of SPRR2F expression (Fig. 5d), three patients with distinct clinical response trajectories were selected: a responder (≥75% reduction in PASI at week 16), a non-responder (<50% reduction in PASI at week 16), and a patient with relapse, defined as an initial improvement in PASI at week 2 followed by worsening beyond the baseline PASI at week 16. Clinical and demographic characteristics of the patients are provided in **Supplementary Table 1**.

FFPE tissue sections (5 µm) were deparaffinised and subjected to heat-induced antigen retrieval in a microwave-heated water bath for 20 minutes, using either Tris-EDTA buffer (pH 9) or citrate buffer (pH 6). After cooling, slides were washed three times with Dako wash buffer (Agilent Technologies, Glostrup, Denmark) (five minutes per wash) and blocked for 45 minutes with either 3% BSA or 5% host-specific serum diluted in Antibody Diluent (Agilent Technologies, Glostrup, Denmark). The sections were then incubated overnight at 4°C with primary antibodies listed in **Supplementary Table 39**, diluted in 5% (v/v) host serum or Antibody Diluent (Agilent Technologies, Glostrup, Denmark) at the concentration specified in **Supplementary Table 39**, or with corresponding isotype controls. After subsequent washing, the slides were incubated with the appropriate secondary antibodies (**Supplementary Table 39**) for 45 minutes at room temperature. Finally, the sections were washed three times and mounted using DAPI-containing Fluoromount (Invitrogen™, Massachusetts, USA).

For quantification of the SPRR2F-positive area, the total epidermal area was first manually delineated based on the DAPI signal identifying DAPI-positive nuclei. The area of red fluorescence corresponding to SPRR2F expression was then quantified within the delineated epidermal region. SPRR2F-positive area was expressed as a percentage of the total epidermal area.

### P-HEK cell culture

We obtained commercially available P-HEK-diseased human epidermal KCs from non-lesional skin of a Caucasian male donor in his 60s diagnosed with psoriasis (Lonza, Basel, Switzerland). The cells were maintained in KGM® Gold Keratinocyte Growth Medium BulletKit® (Lonza, Switzerland) and passaged using trypsin-EDTA 0.05%/0.02% in PBS (PANBiotech, Germany), followed by a trypsin-neutralizing solution (PromoCell, Heidelberg, Germany). For downstream experiments, we seeded the cells into 24-well plates.

### SPRR2F gene knockdown with siRNA and cytokine stimulation

P-HEK cultures at 60–70% confluence in 24-well plates were transfected with Silencer Select siRNA targeting *SPRR2F* (s13391, Cat# 4392420) or a Silencer Select negative control (Cat# 4390843; Thermo Fisher Scientific, MA, USA), using Lipofectamine RNAiMAX (Thermo Fisher Scientific) according to the manufacturer’s protocol, and incubated for 6 h (37°C, 5% CO₂). siRNA specificity was assessed by BLAST alignment of the guide strand against the human RefSeq RNA database (NCBI); the only full-length, 100%-identity hits were two annotated SPRR2F transcript isoforms (NM_001014450.3, NM_001382255.1), consistent with a design intended to target all known variants of the gene, whereas no full-length alignments of functional relevance were found for SPRR2A, SPRR2D, or SPRR2E. We therefore attribute the concurrent downregulation of other SPRR2 family members in knockdown cells to transcriptional co-dependency within the SPRR2 gene cluster (chromosome 1q21) rather than to direct off-target silencing. Cells were subsequently stimulated for 24 h with a psoriasis-relevant cytokine cocktail (50 ng/mL each of recombinant human IL-17A, IL-23, and TNF-α; R&D Systems and BioLegend, as specified).

### RNA isolation, cDNA conversion, and real-time RT-PCR

We extracted total RNA from cell lysates using QIAzol Lysis Reagent (Qiagen, Hilden, Germany). First-strand cDNA was synthesized with the RevertAid First Strand cDNA Synthesis Kit (Thermo Scientific™, Massachusetts, USA). Quantitative PCR (qPCR) was performed on the StepOnePlus Real-Time PCR System (Applied Biosystems™, Massachusetts, USA) using SYBR Green PCR Master Mix (Applied Biosystems™, Massachusetts, USA).

### Bulk RNA-seq and bioinformatic analysis

We performed four independent experiments, each comprising four experimental conditions, for a total of sixteen RNA samples. Total RNA from the SPRR2F knockdown experiments was submitted to NOVOGENE GmbH (Planegg, Munich, Germany) for bulk RNA sequencing. RNA concentration was quantified using a Qubit Fluorometer (Thermo Scientific™, Massachusetts, USA), and RNA integrity and purity were assessed using an Agilent 5400 system (Agilent Technologies, Glostrup, Denmark). RNA quality control, poly(A)-enriched mRNA library preparation, and paired-end sequencing (2 × 150 bp; ∼6 GB raw data per sample) were performed by NOVOGENE GmbH following standard protocols.

Raw sequencing reads were quality controlled using FASTQC (Andrews, S. (2010)) and aligned to the human reference genome (*Homo sapiens* GRCh38, Ensembl release 84) using HISAT2^72^ with a splice-aware SNP- and transcript-informed index. Resulting alignments were sorted and indexed with samtools^73^, and gene expression levels were quantified using featureCounts^74^ with paired-end counting and Homo_sapiens annotations (GRCh38.84).

For downstream analysis, we used the DESeq2^75^ package (v.1.51.7) in R (v4.5.0). After removing non-coding transcripts, the count matrix was restricted to protein-coding genes. Differential expression across the four experimental groups (negative control + vehicle, negative control + cytokines, SPRR2F siRNA + vehicle, SPRR2F siRNA + cytokines) was modelled with a metadata-driven design formula and tested using Likelihood Ratio Tests (LRT). To improve statistical power, we applied Independent Hypothesis Weighting (IHW) and considered genes significant at an adjusted *p*-value < 0.05. For downstream visualization and exploratory analysis, the raw counts were transformed with the variance stabilizing transformation (VST), which yields homoscedastic data suitable for clustering, principal-component analysis, and heat-map generation.

## Data Availability

All data produced in the present study are available upon reasonable request to the authors

## Acknowledgements

We thank Lou Plath for assistance with the Keyence microscope and image acquisition. We thank Silke Heymann for technical assistance with the preparation of histological samples. We are also grateful to Madeleine Jares, Amelie Kleefeldt, and Sandra Bruhn for processing the skin rinsing samples. We sincerely thank all patients and their families for generously contributing the human specimens used in this study.

## Contributions

T.S., Y.G. and D.T. conceptualized the study. T.S. and Y.G. developed the methodology of this study. Y.G. and N.P. carried out formal analysis and created the figures and tables. T.S., N.P., S.M., X.T. conducted the investigation. Y.G., N.P. and D.B. handled data visualization. H. Z., S. S., D.A.D.L., C.P., and L.P. collected patient samples and clinical information. T.S., N.P., and Y.G. wrote the original draft and did the writing, reviewing and editing of the manuscript. D.T. and T.S. secured funding. All authors read, reviewed and approved the final version of the manuscript. Generative artificial intelligence (AI) was used to assist in generating Figure 1A for this manuscript. The authors reviewed and approved the final figure and take full responsibility for its content and accuracy.

