## Supplementary Figure 1 for "High definition longitudinal spatial atlas of psoriasis reveals SPRR2F⁺ keratinocytes and distinct tissue-resolution trajectories under IL-17 and IL-23 inhibition"

**Supplementary Figures**

*Supplementary Figure 1*

Supplementary Fig. 1A	Single-cell UMAP of skin cell clusters.

Supplementary Fig. 1B	Marker gene expression defining keratinocyte subclusters.

Supplementary Fig. 1C	Marker gene expression defining immune cell subclusters.

Supplementary Fig. 1D	Marker gene expression defining adnexal cell subclusters.

Supplementary Fig. 1E	Marker gene expression defining stromal cell subclusters.

Supplementary Fig. 1F	Marker gene expression defining the melanocyte cluster.

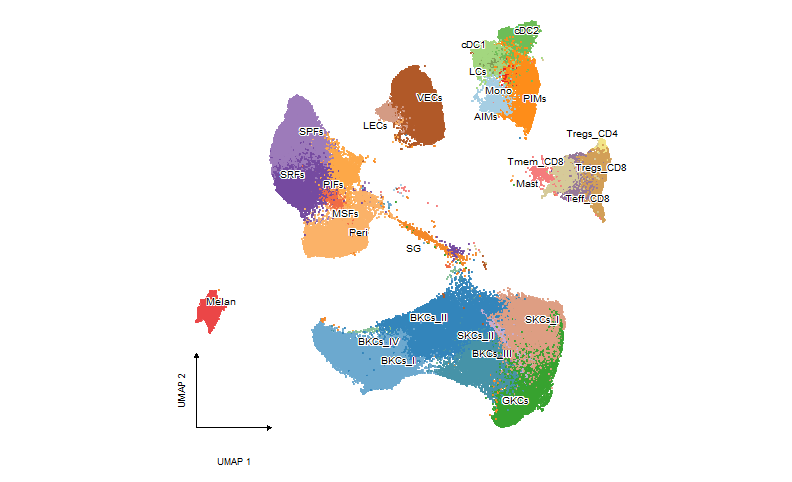

**Supplementary Fig. 1A | *scRNA-seq UMAP of skin cell clusters.***

Uniform manifold approximation and projection (UMAP) of all profiled cells, colored and labeled by cluster. Clusters span keratinocyte (basal KCs: BKCs_I, BKCs_II, BKCs_III, and BKCs_IV; spinous KCs: SKCs_I and SKCs_II; and granular KCs: GKCs), fibroblast and perivascular cells (secretory-papillary FBs: SPFs; secretory-reticular FBs: SRFs; mesenchymal FBs: MSFs; pro-inflammatory FBs: PIFs; and pericytes: Peri), endothelial cells (vascular ECs: VECs; and lymphatic ECs: LECs), sebaceous glands (SG), melanocytes (Melan), and immune cells (T cells: Tregs_CD4, Tregs_CD8, Tmem_CD8, Teff_CD8) dendritic cells (cDC1, cDC2, LCs), monocytes/macrophages (Mono), mast cells (Mast), AIMs and PIMs.

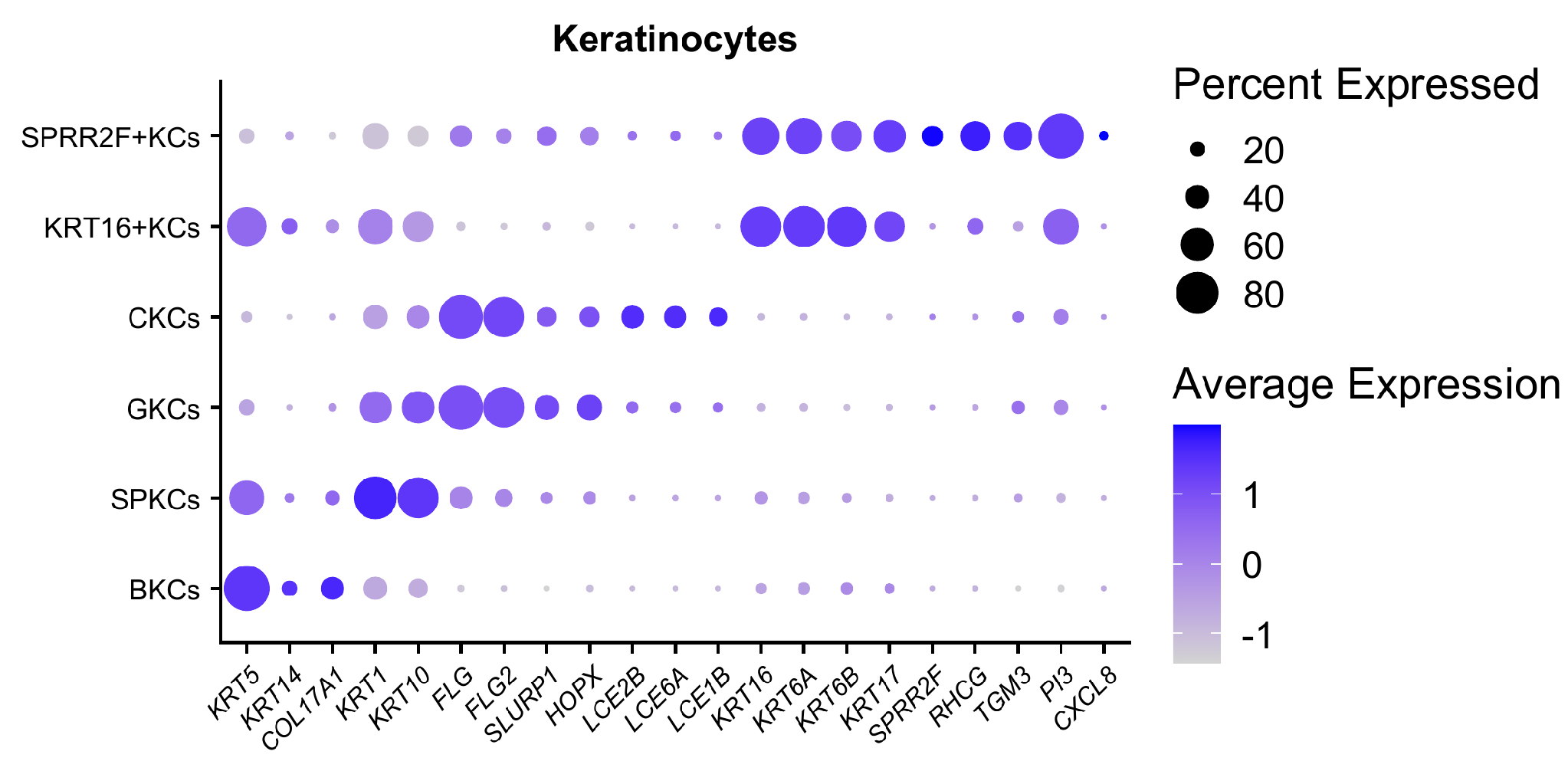

**Supplementary Fig. 1B | *Marker gene expression defining keratinocyte subclusters.***

Dot plot of canonical marker genes across keratinocyte subclusters (BKCs, SPKCs, GKCs, CKCs, KRT16+KCs, SPRR2F+KCs). Dot color represents scaled average expression; dot size represents the percentage of cells expressing each gene.

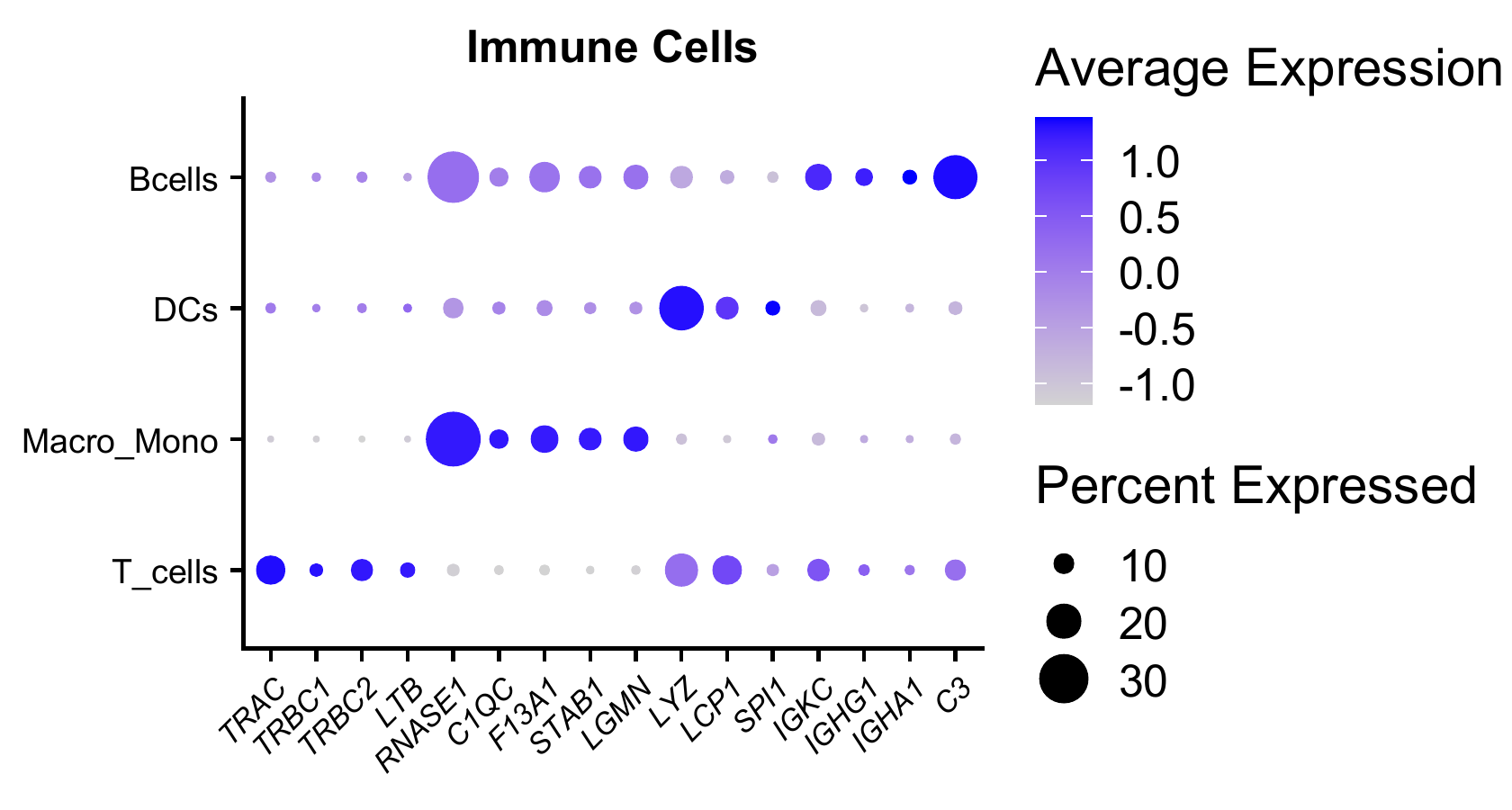

**Supplementary Fig. 1C | *Marker gene expression defining immune cell clusters.***

Dot plot of canonical marker genes across immune cell clusters (T_cells, DCs, Macro_Mono, Bcells). Dot color represents scaled average expression; dot size represents the percentage of cells expressing each gene.

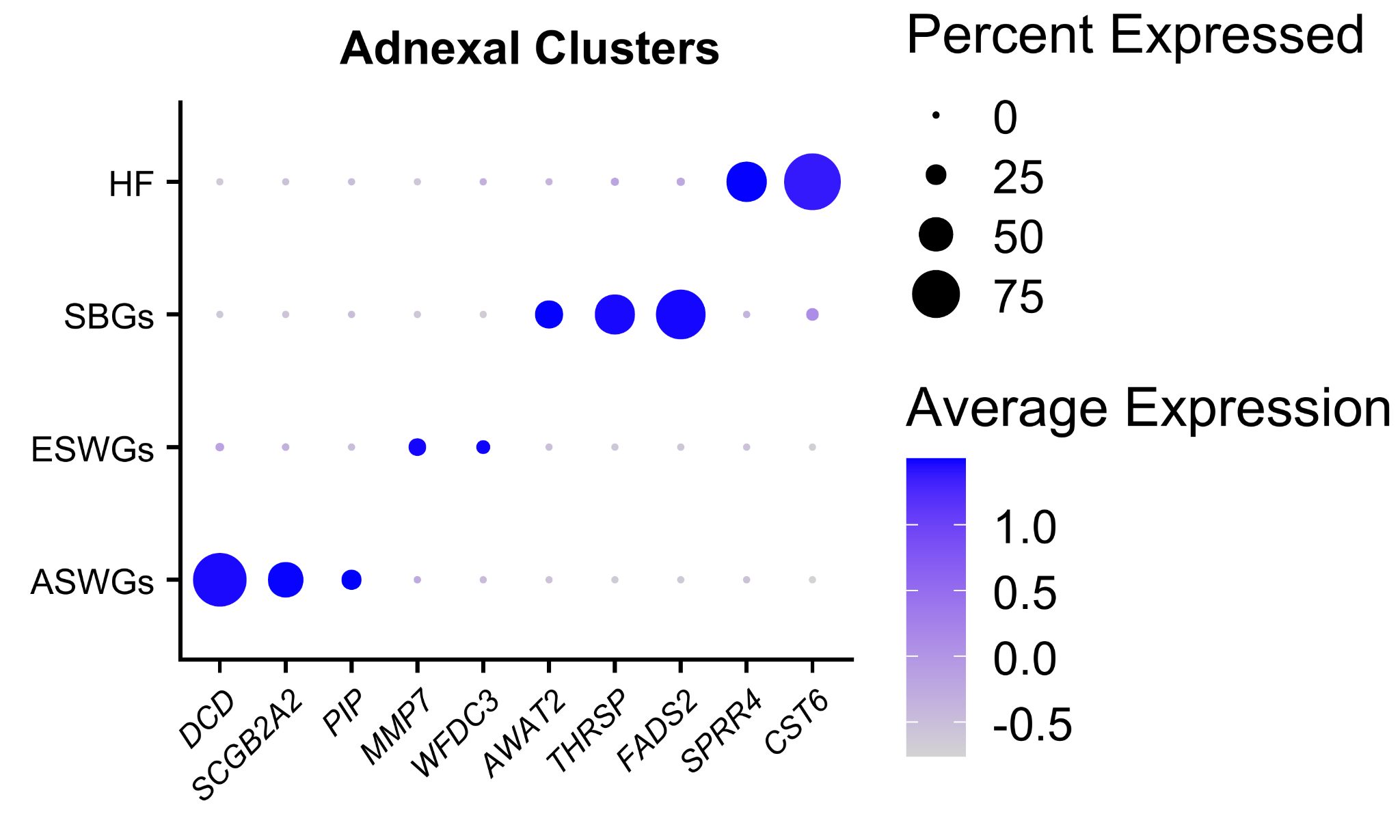

**Supplementary Fig. 1D | *Marker gene expression defining adnexal clusters.***

Dot plot of canonical marker genes across adnexal cell subclusters (ASWGs, ESWGs, SBGs, HF). Dot color represents scaled average expression; dot size represents the percentage of cells expressing each gene.

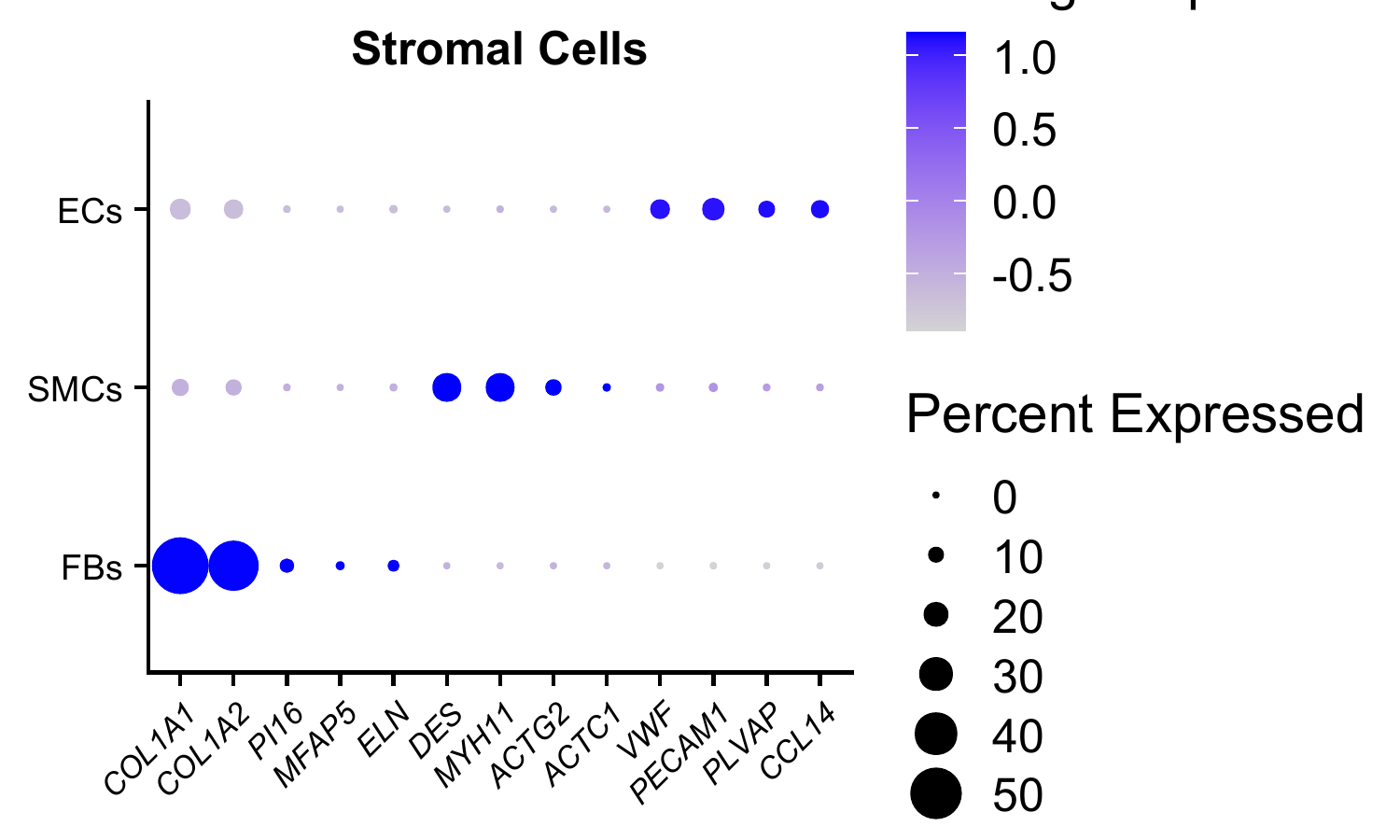

**Supplementary Fig. 1E | *Marker gene expression defining stromal cell subclusters.***

Dot plot of canonical marker genes across stromal cell subclusters (FBs, SMCs, ECs). Dot color represents scaled average expression; dot size represents the percentage of cells expressing each gene.

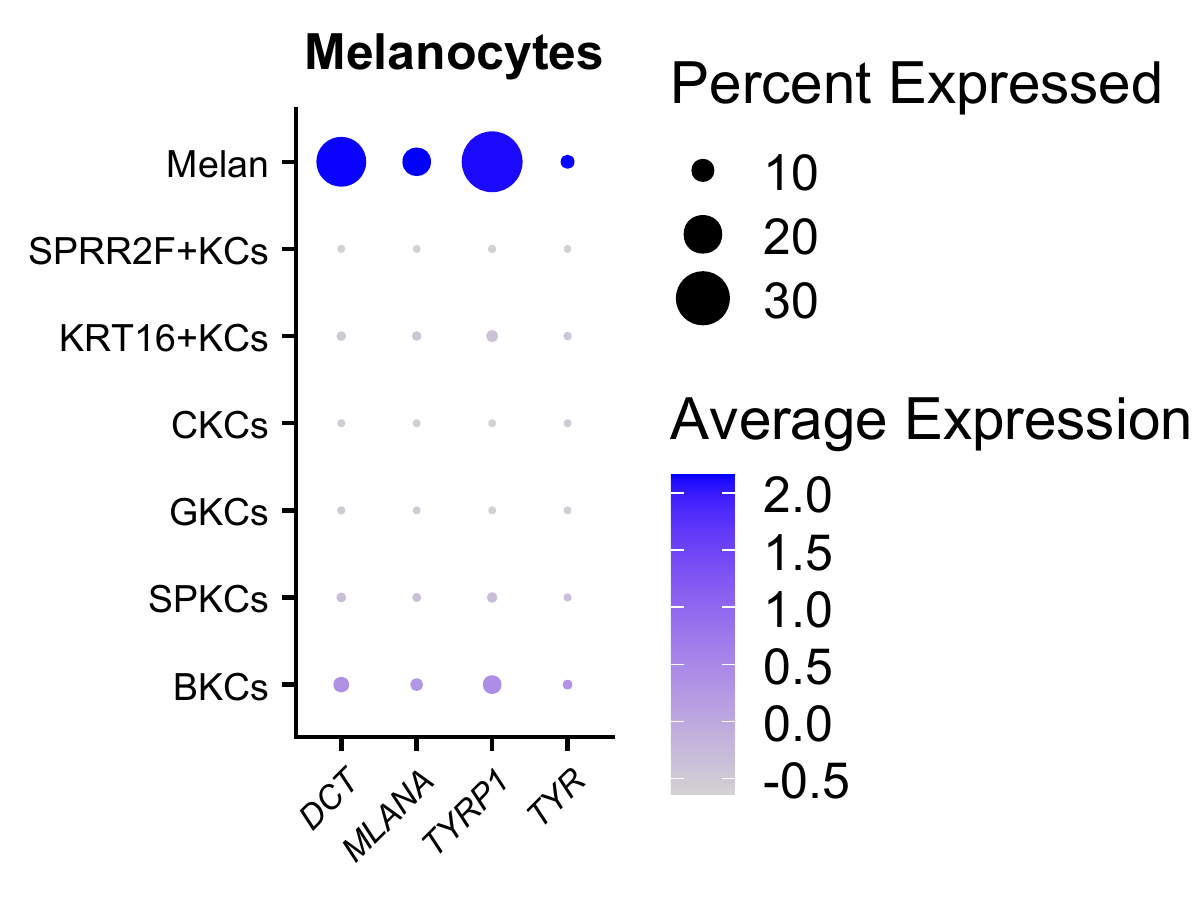

**Supplementary Fig. 1F | *Marker gene expression defining the melanocyte cluster.***

Dot plot of canonical melanocyte marker genes (*DCT, MLANA, TYRP1, TYR*) in the melanocyte cluster (Melan) relative to keratinocyte subclusters. Dot color represents scaled average expression; dot size represents the percentage of cells expressing each gene.
