## Supplementary Figure 2 for "High definition longitudinal spatial atlas of psoriasis reveals SPRR2F⁺ keratinocytes and distinct tissue-resolution trajectories under IL-17 and IL-23 inhibition"

**Supplementary Figures**

*Supplementary Figure 2*

Supplementary Fig. 2A	Cell-type proportions of endothelial cells and smooth muscle cells at baseline, lesional versus non-lesional.

Supplementary Fig. 2B	Differential gene expression in SPRR2F+ KCs at baseline, lesional versus non-lesional.

Supplementary Fig. 2C	Ligand-receptor communication network at baseline.

Supplementary Fig. 2D	Differential gene expression in T cells and DCs at baseline, lesional versus non-lesional.

Supplementary Fig. 2E	Pathway activity at baseline, keratinocyte clusters and FBs.

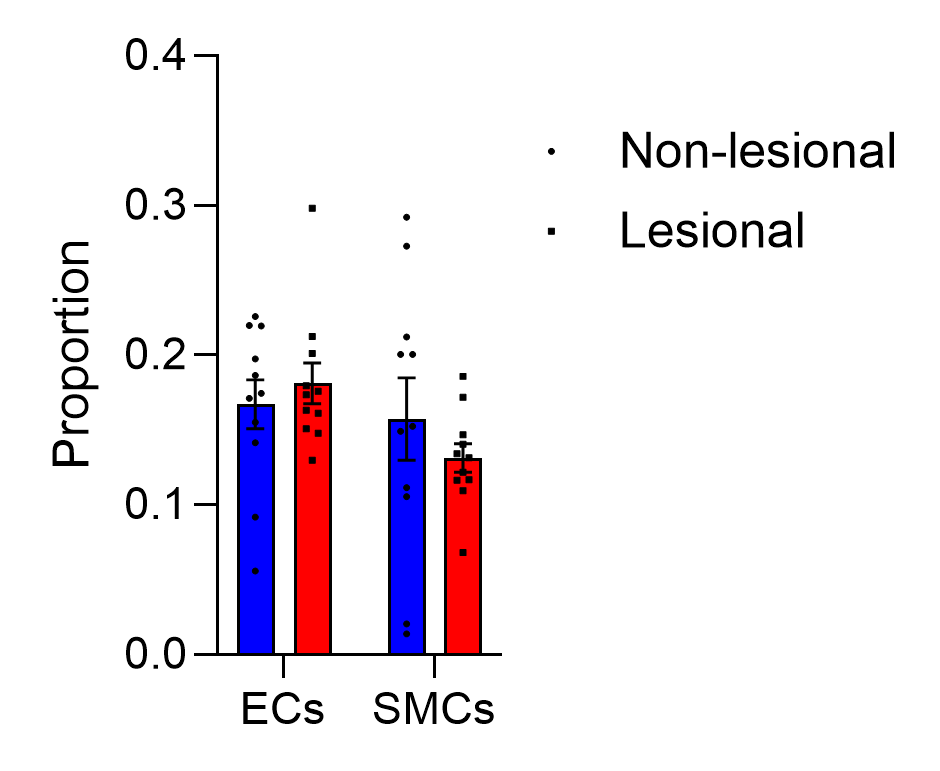

**Supplementary Fig. 2A | *Cell-type proportions of endothelial cells and smooth muscle cells at baseline, lesional versus non-lesional.***

Bar plots of the proportion of endothelial cells (ECs) and smooth muscle cells (SMCs) among non-keratinocyte clusters in non-lesional and lesional skin at baseline. Points represent individual samples. Bars represent mean proportion ± SEM. ns, not significant.

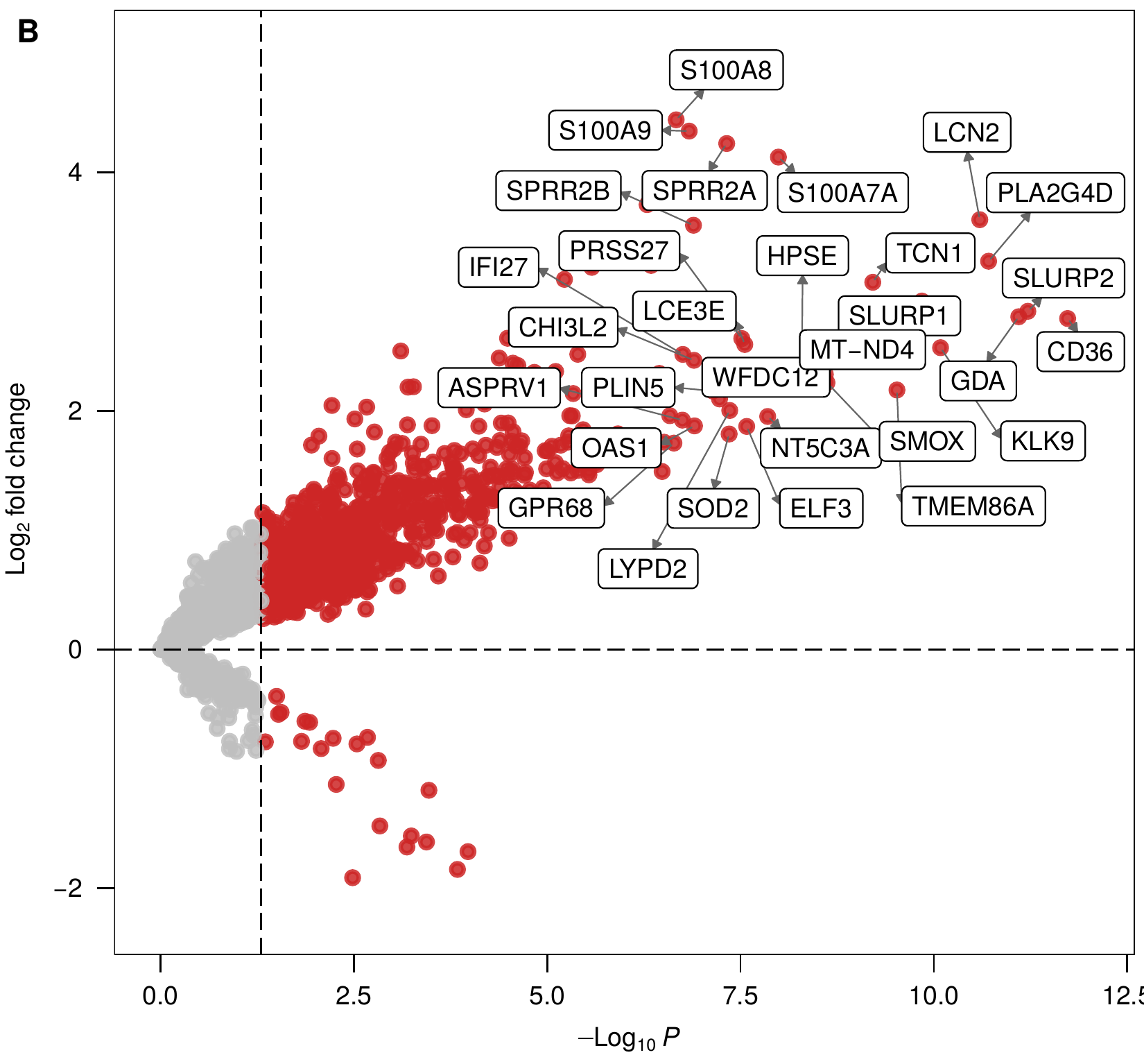

**Supplementary Fig. 2B | *Differential gene expression in SPRR2F+ KCs at baseline, lesional versus non-lesional.***

Volcano plot of log2 fold change (y-axis) versus -log10 P (x-axis) for lesional versus non-lesional SPRR2F+ KCs at baseline. Significant genes highlighted; the 30 most significant genes labeled.

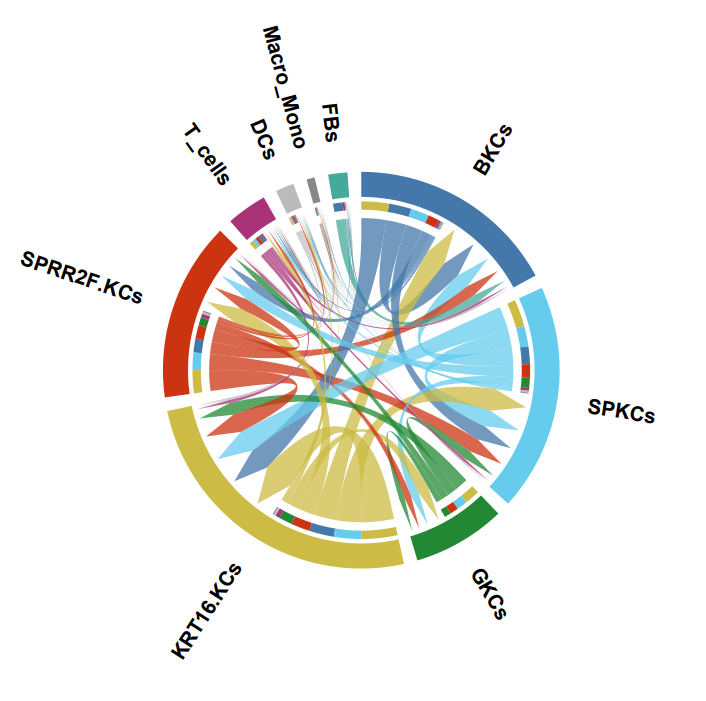

**Supplementary Fig. 2C | *Ligand-receptor communication network at baseline.***

Chord diagram of ligand-receptor interactions among keratinocyte, immune, and stromal clusters (BKCs, SPKCs, GKCs, KRT16+ KCs, SPRR2F+ KCs, T cells, DCs, Macro/Mono, FBs) at baseline. Chord width = number of ligand-receptor pairs between sender and receiver; arrows and differential height indicate directionality; color = sender cluster.

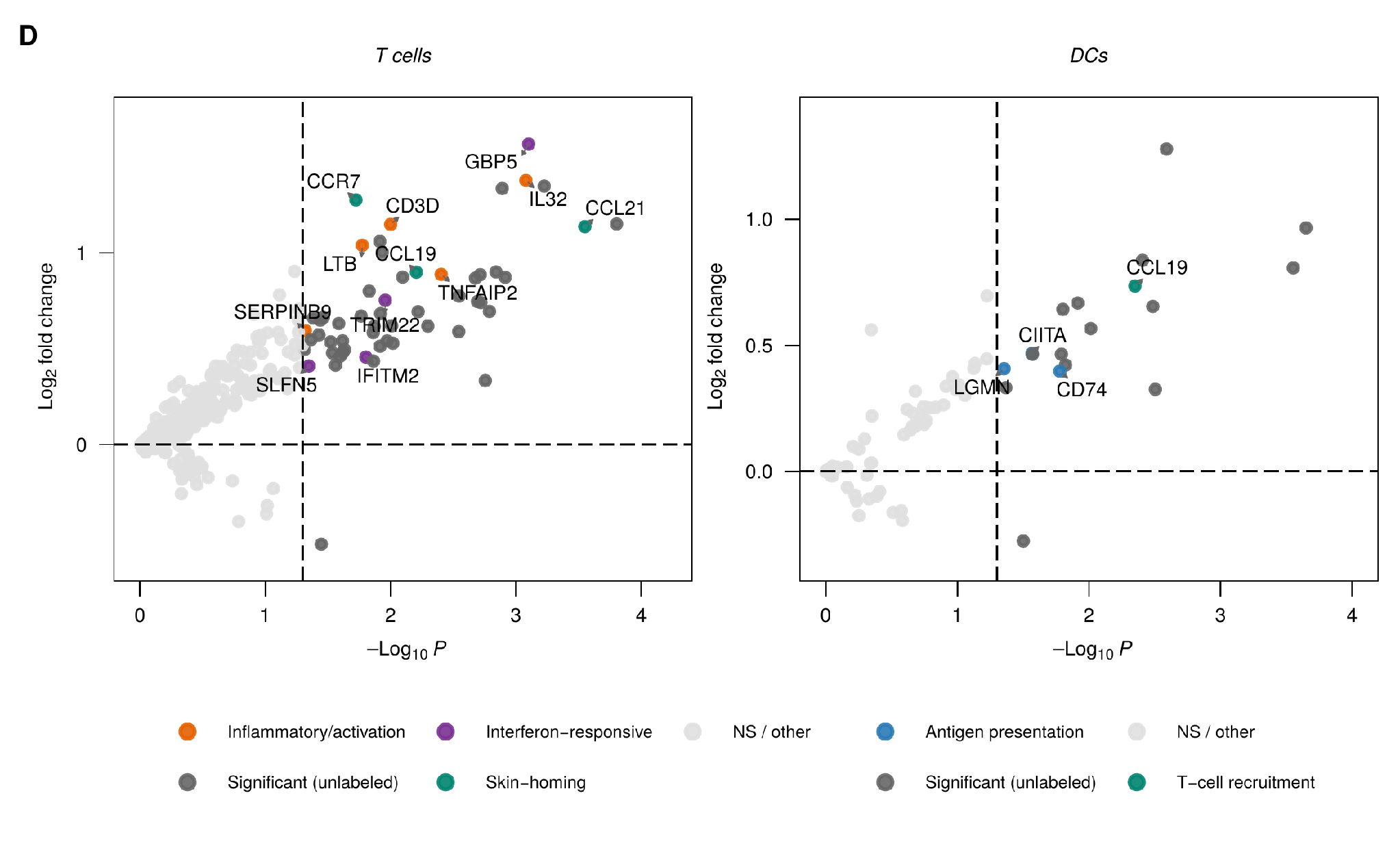

**Supplementary Fig. 2D | *Differential gene expression in T cells and DCs at baseline, lesional versus non-lesional.***

Volcano plots of log2 fold change (y-axis) versus -log10 P (x-axis) for T cells and DCs, lesional versus non-lesional skin at baseline. Points colored by functional category: interferon-responsive, inflammatory/activation, skin-homing (T cells); antigen presentation, T-cell recruitment (DCs); or not significant. Selected genes labeled.

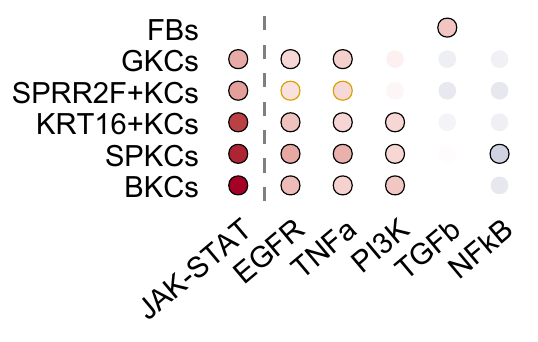

**Supplementary Fig. 2E | *Pathway activity at baseline, keratinocyte clusters and FBs.***

Dot matrix of PROGENy pathway activity scores (JAK-STAT, EGFR, TNFa, PI3K, TGFb, NFkB) across keratinocyte clusters and FBs at baseline. Dot color = activity score; dot size = magnitude of activity score; dot outline = statistical significance (padj < 0.05 versus not significant).
