## Supplementary Figure 3 for "High definition longitudinal spatial atlas of psoriasis reveals SPRR2F⁺ keratinocytes and distinct tissue-resolution trajectories under IL-17 and IL-23 inhibition"

**Supplementary Figures**

*Supplementary Figure 3*

**Contents**

Supplementary Fig. 3A	Immune and stromal cell-type proportions under IL-17 pathway inhibition.

Supplementary Fig. 3B	Differential gene expression under IL-17 pathway inhibition at week 2, all keratinocyte clusters.

Supplementary Fig. 3C	Reversal of the SPRR2F+ KC lesional gene program under IL-17 pathway inhibition at week 16.

Supplementary Fig. 3D	Effect size and overlap of differentially expressed genes across keratinocyte clusters under IL-17 pathway inhibition at week 16.

Supplementary Fig. 3E	Ligand-receptor prioritization changes involving keratinocyte clusters under IL-17 pathway inhibition.

Supplementary Fig. 3F	Immune and stromal transcriptional response to IL-17 pathway inhibition, baseline versus week 16.

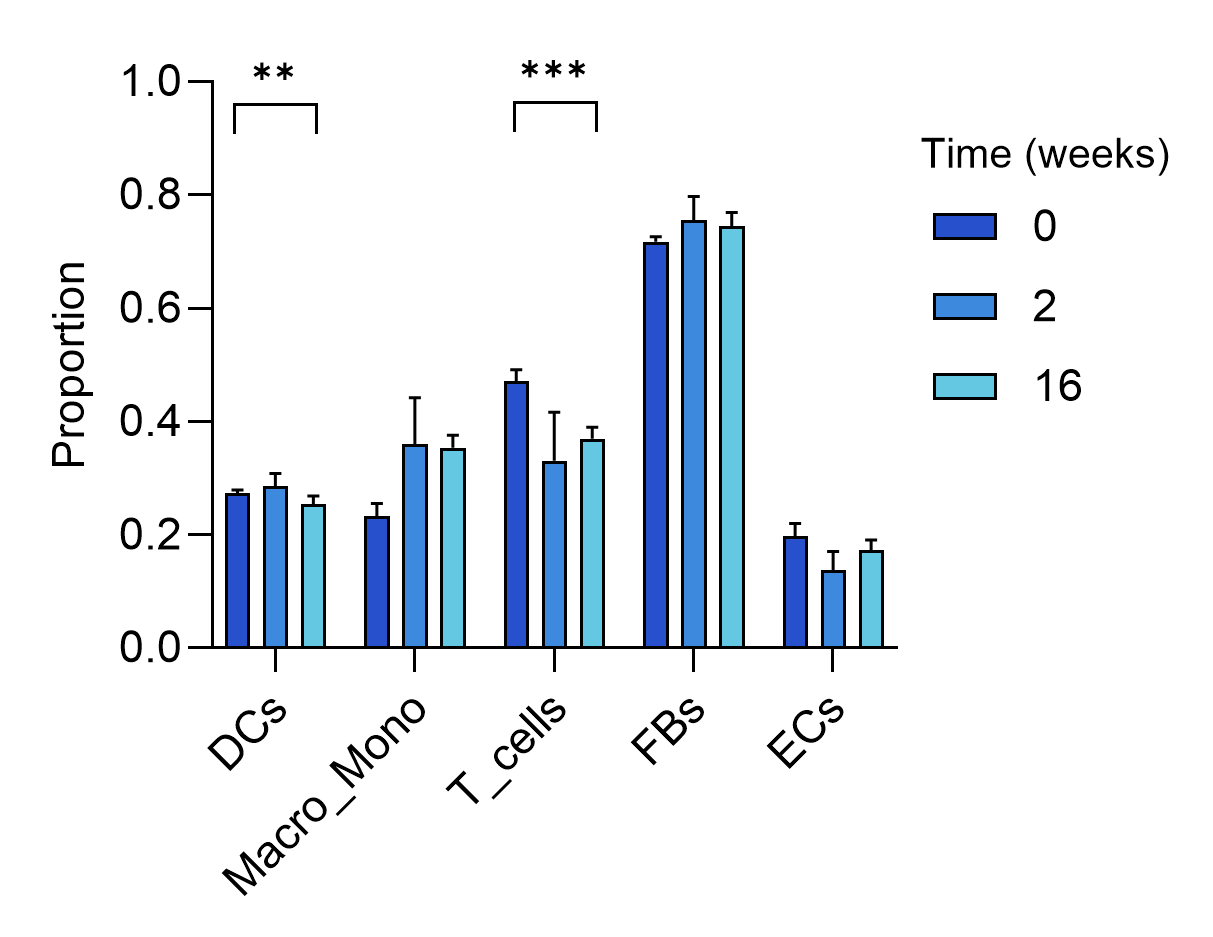

**Supplementary Fig. 3A | *Immune and stromal cell-type proportions under IL-17 pathway inhibition.***

Bar plots of the proportion of dendritic cells (DCs), macrophages/monocytes (Macro_Mono), T cells, fibroblasts (FBs), and endothelial cells (ECs) among non-keratinocyte clusters at baseline (week 0) and following IL-17 pathway inhibition (week 2, week 16), calculated using propeller. Bars represent mean proportion ± SEM. Brackets indicate significance of the change across timepoints (** P < 0.01; *** P < 0.001; ns, not significant).

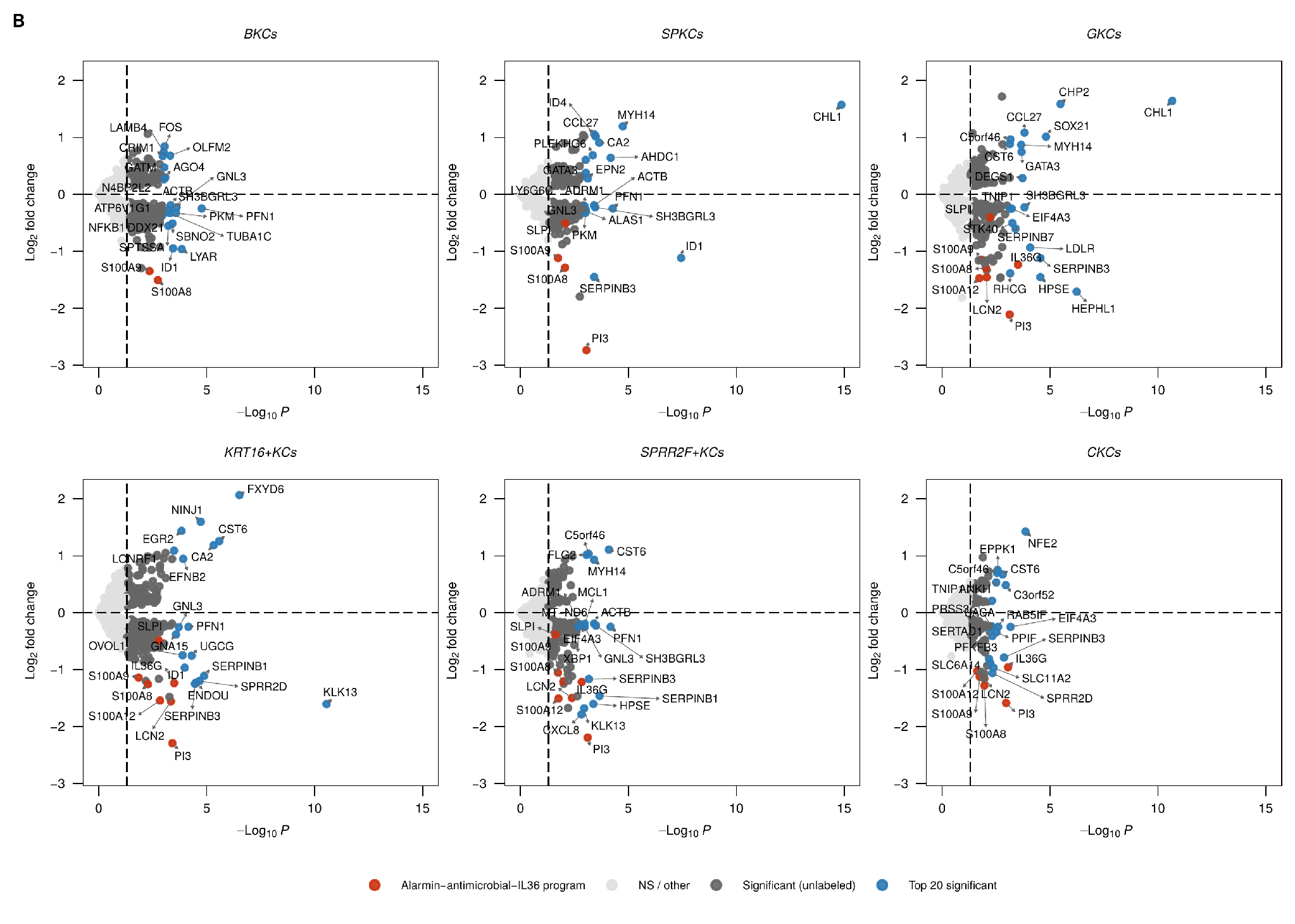

**Supplementary Fig. 3B | *Differential gene expression under IL-17 pathway inhibition at week 2, all keratinocyte clusters.***

Volcano plots of log2 fold change (y-axis) versus -log10 P (x-axis) for six keratinocyte clusters (BKCs, SPKCs, GKCs, KRT16+ KCs, SPRR2F+ KCs, CKCs) under IL-17 pathway inhibition at week 2. Points colored by category: alarmin-antimicrobial-IL36 program genes, top 20 most significant genes, other significant genes, and not significant.

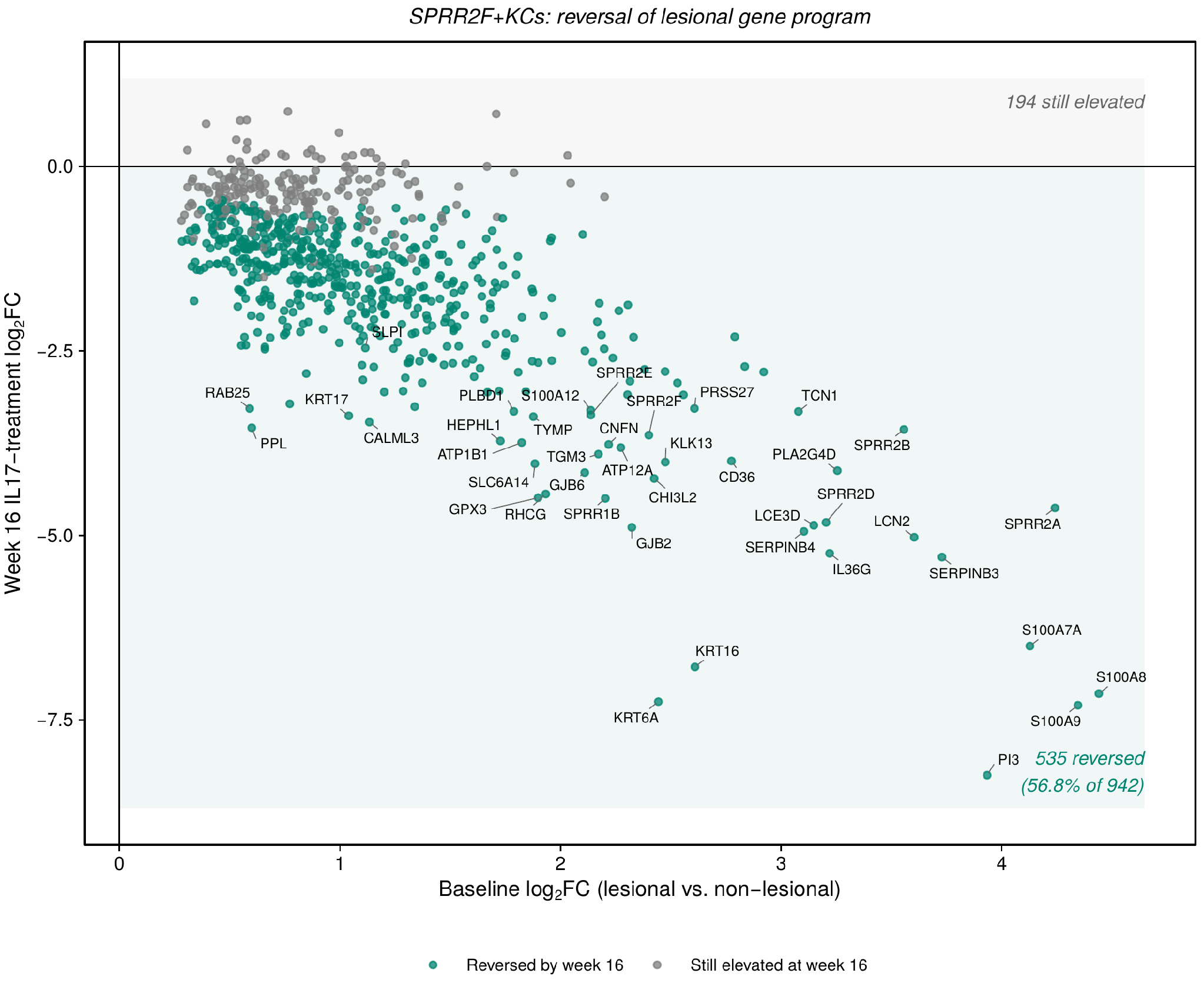

**Supplementary Fig. 3C | *Reversal of the SPRR2F+ KC lesional gene program under IL-17 pathway inhibition at week 16.***

Scatter plot of baseline log2FC (lesional versus non-lesional, x-axis) versus week-16 IL-17-axis-treatment log2FC (y-axis) for genes significantly upregulated in lesional SPRR2F+ KCs at baseline. Points colored by reversal status: reversed by week 16 or still elevated at week 16. Selected genes labeled.

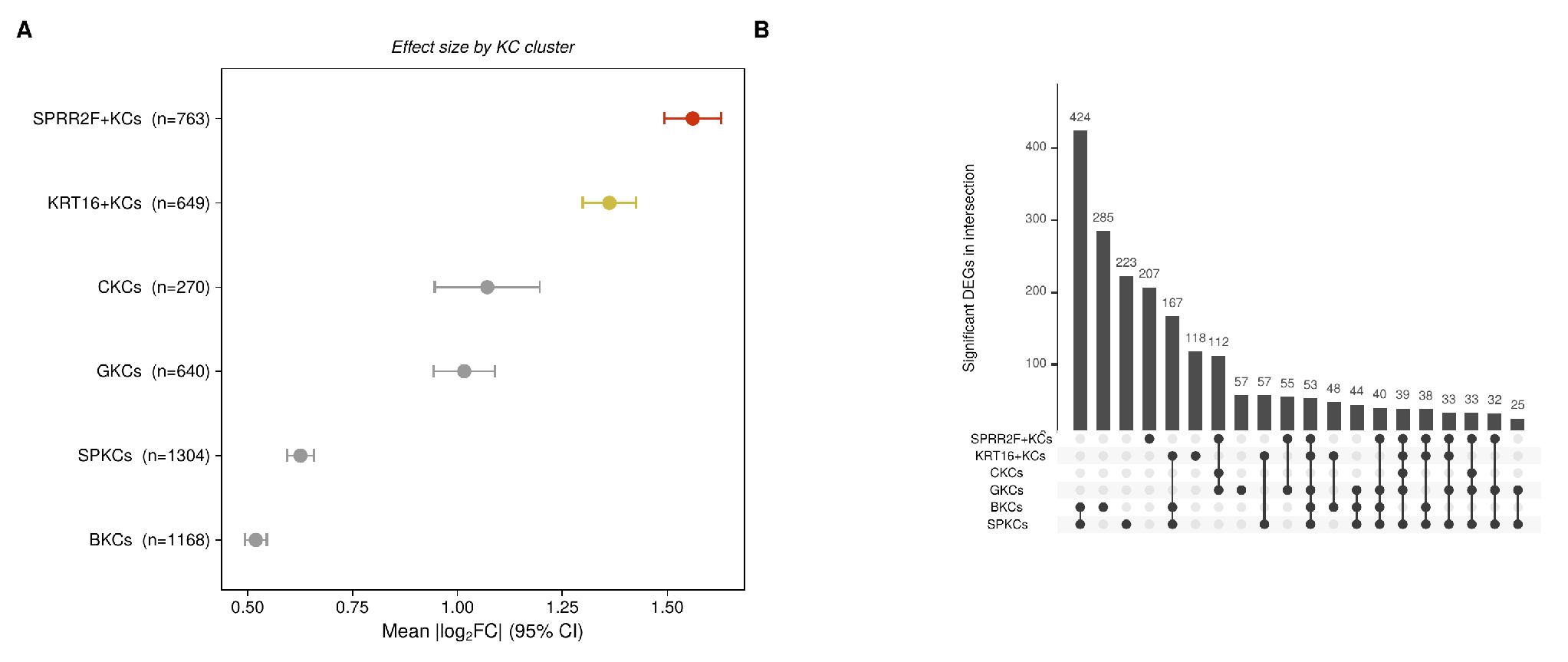

**Supplementary Fig. 3D | *Effect size and overlap of differentially expressed genes across keratinocyte clusters under IL-17 pathway inhibition at week 16.***

(A) Forest plot of mean absolute log2FC (95% CI) among significant genes (lfsr < 0.05) per keratinocyte cluster. (B) UpSet plot of the intersections among each cluster's significant gene set, ordered by intersection size.

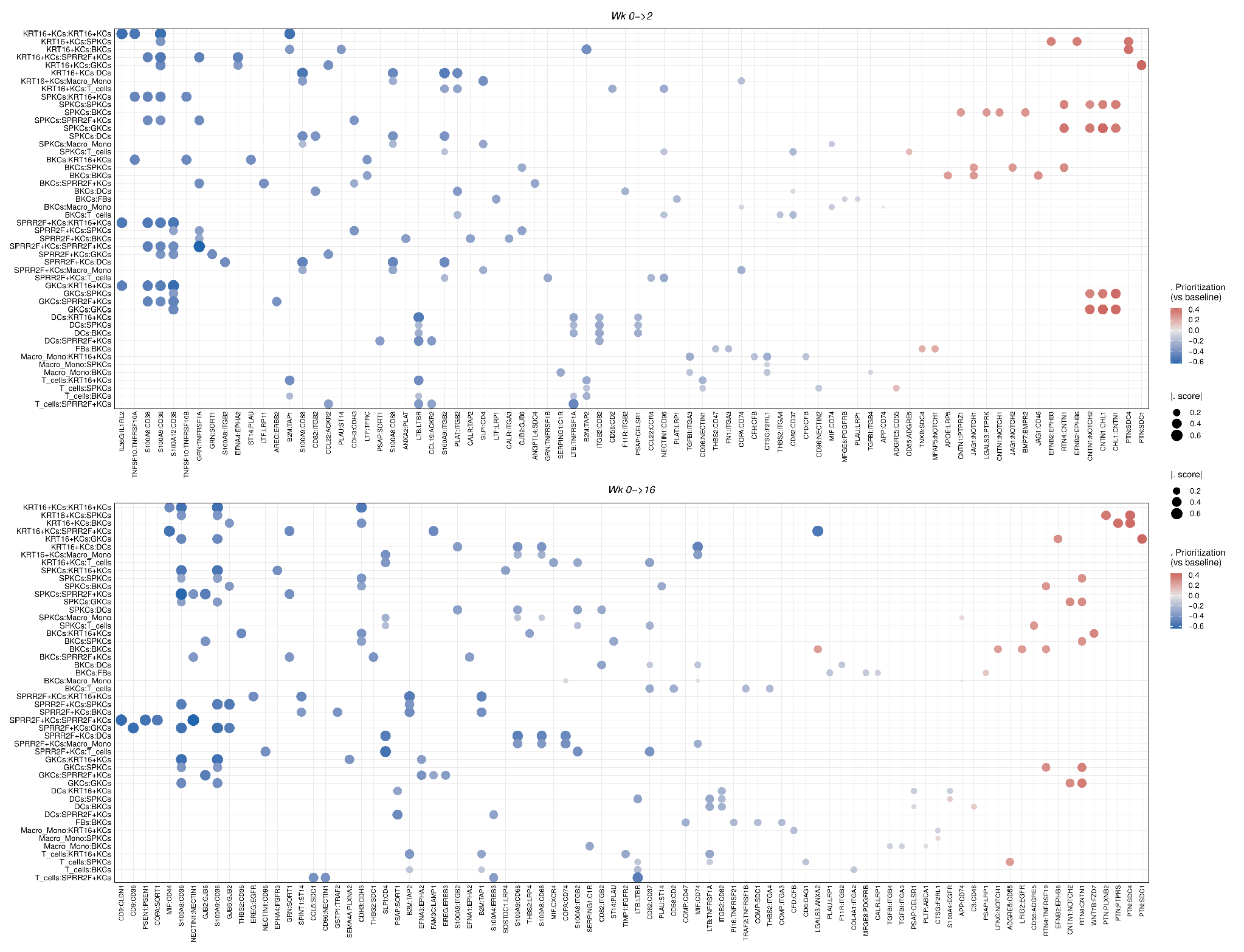

**Supplementary Fig. 3E | *Ligand-receptor prioritization changes involving keratinocyte clusters under IL-17 pathway inhibition.***

Dot-plot matrix of the top changing ligand-receptor pairs (by prioritization-score delta versus baseline) for each sender:receiver combination involving a keratinocyte cluster, at week 2 and week 16 under IL-17 pathway inhibition. Dot color = change in prioritization score versus baseline; dot size = magnitude of that change.

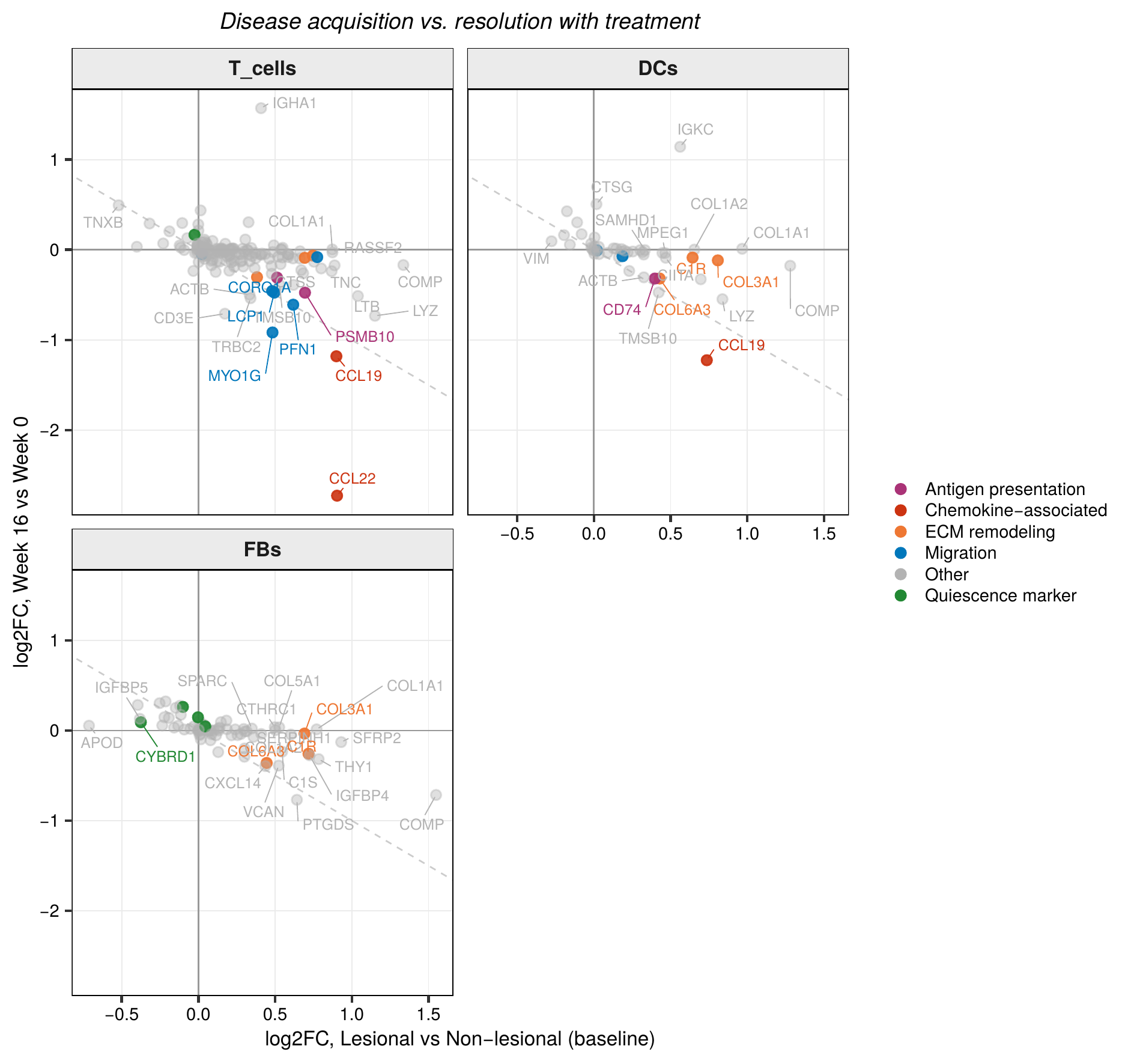

**Supplementary Fig. 3F | *Immune and stromal transcriptional response to IL-17 pathway inhibition, baseline versus week 16.***

Scatter plots of baseline log2FC (lesional versus non-lesional, x-axis) versus week-16 log2FC (y-axis) under IL-17 pathway inhibition, shown separately for T cells, DCs, and FBs. Points colored by functional category: antigen presentation, chemokine-associated, ECM remodeling, migration, quiescence marker, or other. Selected genes labeled.
