## Supplementary Figure 4 for "High definition longitudinal spatial atlas of psoriasis reveals SPRR2F⁺ keratinocytes and distinct tissue-resolution trajectories under IL-17 and IL-23 inhibition"

**Supplementary Figures**

*Supplementary Figure 4*

**Contents**

Supplementary Fig. 4A	Keratinocyte cluster proportions under IL-23 inhibition, baseline through week 16.

Supplementary Fig. 4B	Reversal of baseline SPRR2F+ KC genes at Week 2, IL-17 pathway versus IL-23 inhibition.

Supplementary Fig. 4C	Cytokine activity at Week 2, all cytokines.

Supplementary Fig. 4D	Cytokine and pathway activity at Week 2 in SPRR2F+ KCs.

Supplementary Fig. 4E	KRT16+ KC gene reversal and hallmark pathway activity at Week 2.

Supplementary Fig. 4F	GKC gene reversal at Week 2.

Supplementary Fig. 4G	Baseline lesional expression versus Week-2 IL-23 inhibition response in BKCs, SPKCs, and CKCs.

Supplementary Fig. 4H	SPRR2F+ KC gene reversal and keratinization gene families at Week 16.

Supplementary Fig. 4I	SPRR2F+ KC baseline-versus-week-16 correlation and PROGENy pathway activity.

Supplementary Fig. 4J	Cytokine and pathway activity in BKCs, SPKCs, GKCs, and KRT16+ KCs at Week 16.

Supplementary Fig. 4K	Baseline lesional expression versus Week-16 response across KC clusters (RINT-normalized).

Supplementary Fig. 4L	KC-KC ligand-receptor communication network, baseline and treatment.

Supplementary Fig. 4M	FB transcriptional response, baseline versus treatment log2FC.

Supplementary Fig. 4N	T cell transcriptional response, baseline versus treatment log2FC.

Supplementary Fig. 4O	DC transcriptional response, baseline versus treatment log2FC.

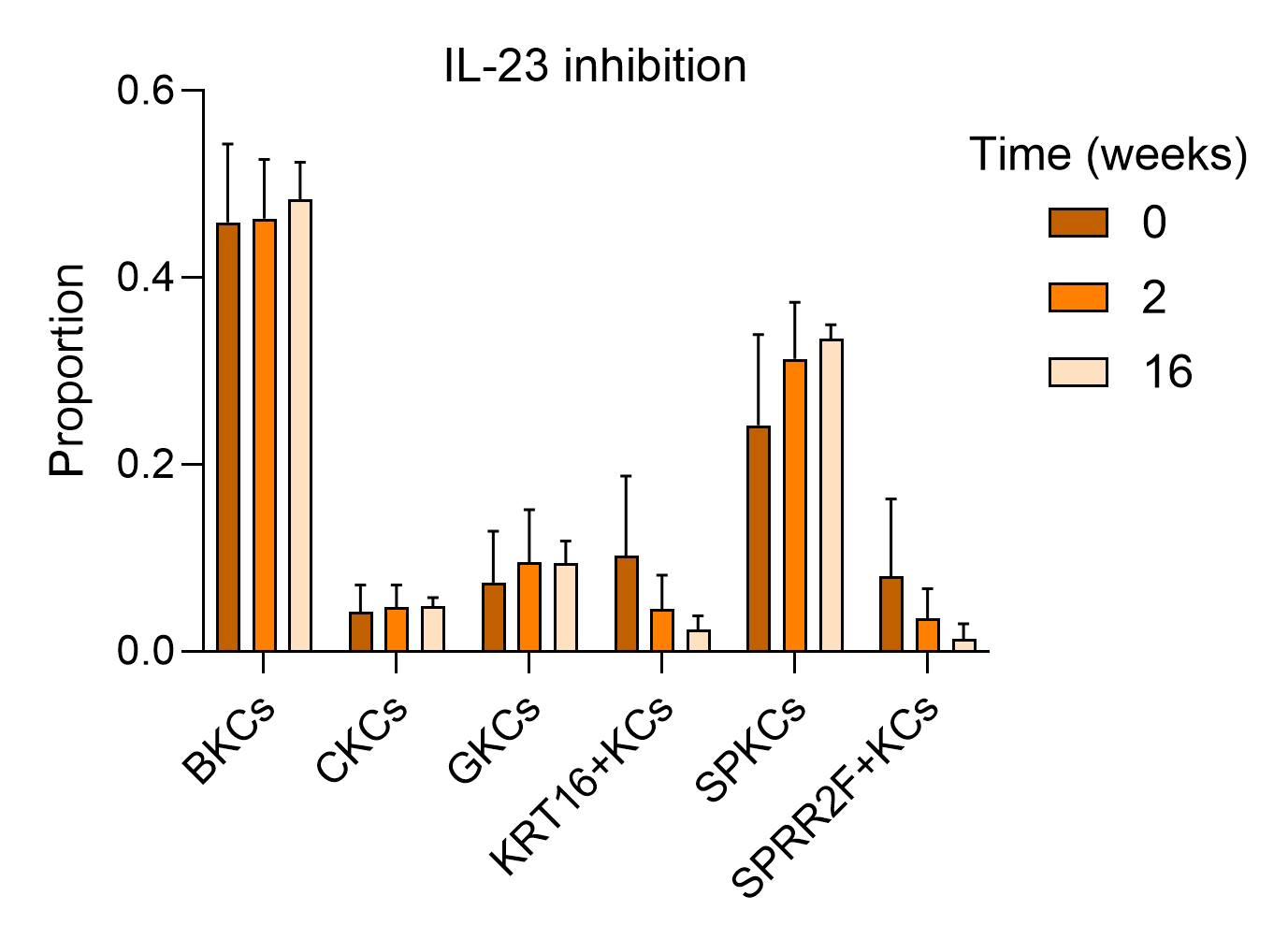

**Supplementary Fig. 4A | *Keratinocyte cluster proportions under IL-23 inhibition, baseline through week 16.***

Bar plots of the proportion of six keratinocyte clusters (BKCs, CKCs, GKCs, KRT16+ KCs, SPKCs, SPRR2F+ KCs) at baseline (week 0) and following IL-23 inhibition (week 2, week 16). Bars represent mean proportion ± SEM.

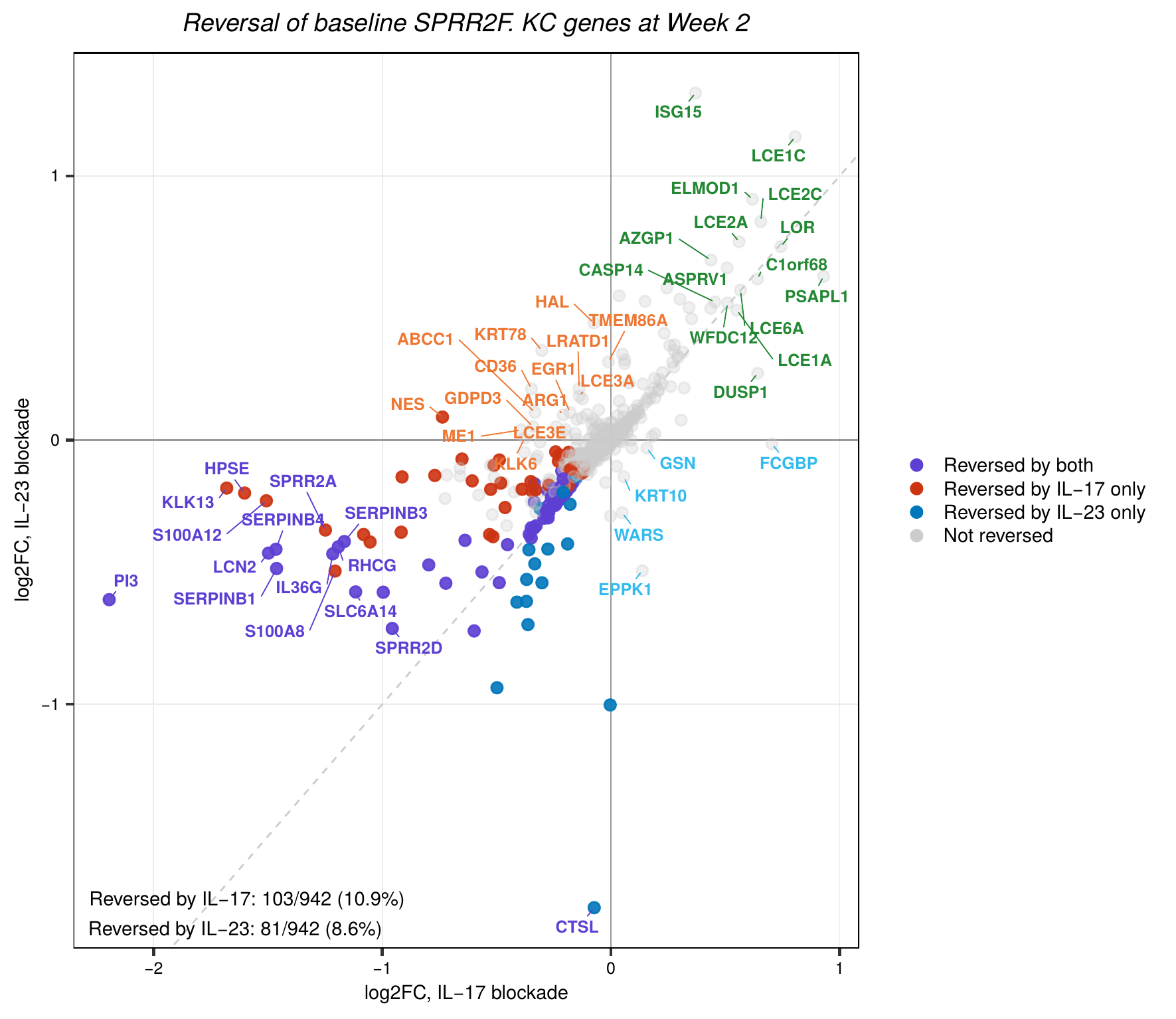

**Supplementary Fig. 4B | *Reversal of baseline SPRR2F+ KC genes at Week 2, IL-17 pathway versus IL-23 inhibition.***

Scatter plot of log2FC under IL-17 pathway blockade (x-axis) versus IL-23 inhibition (y-axis) for genes significantly dysregulated in lesional SPRR2F+ KCs at baseline. Points colored by reversal status: reversed by both treatments, IL-17 pathway only, IL-23 only, or not reversed. Selected genes labeled.

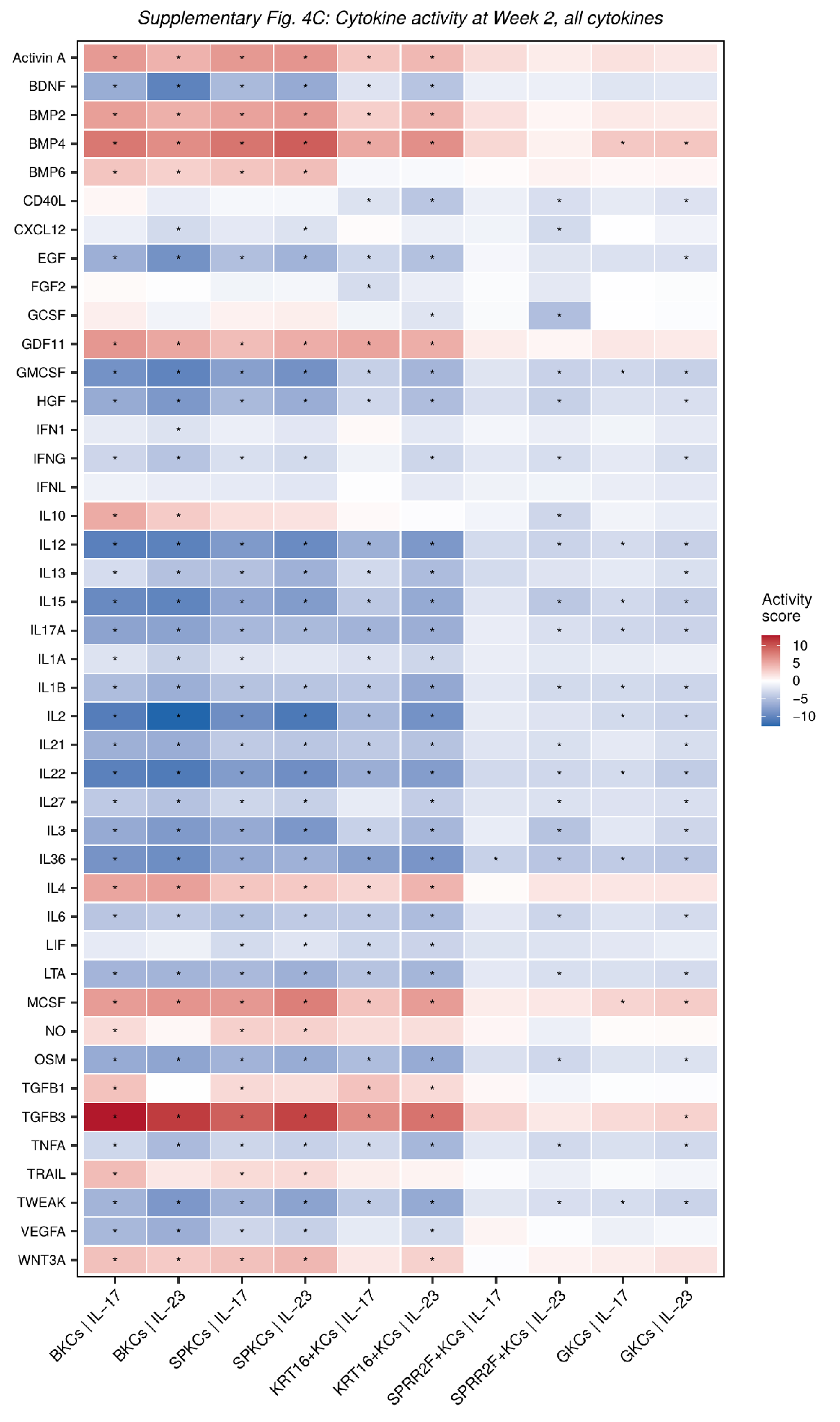

**Supplementary Fig. 4C | *Cytokine activity at Week 2, all cytokines.***

Heatmap of predicted cytokine activity (CytoSig) for 41 cytokines across five KC clusters (BKCs, SPKCs, KRT16+ KCs, SPRR2F+ KCs, GKCs), IL-17 pathway versus IL-23 inhibition. Tile color = activity score; asterisks denote adjusted P < 0.05.

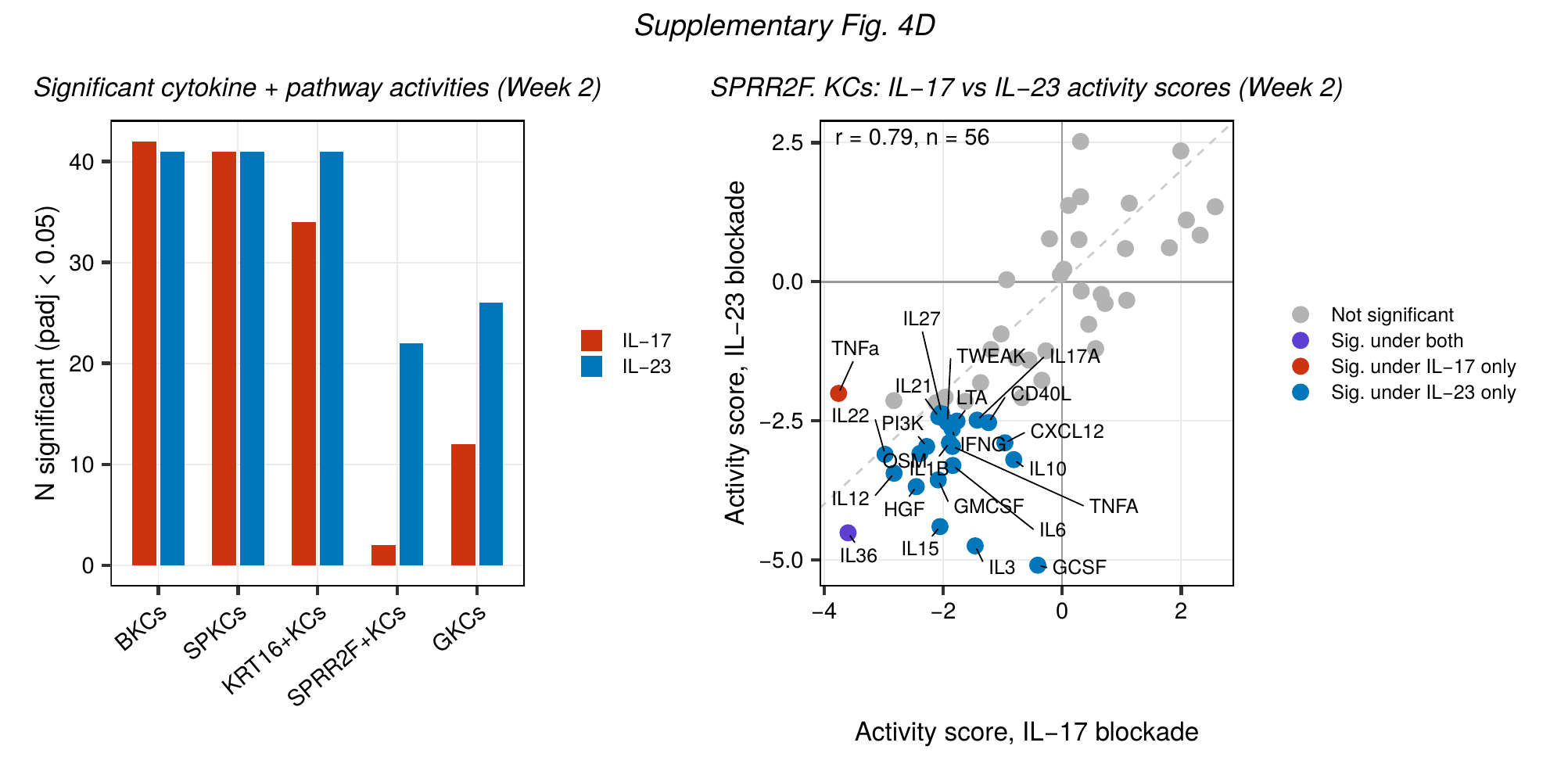

**Supplementary Fig. 4D | *Cytokine and pathway activity at Week 2 in SPRR2F+ KCs.***

(Left) Number of cytokine/pathway activities reaching adjusted P < 0.05 per KC cluster, IL-17 pathway versus IL-23 inhibition. (Right) Scatter plot of activity scores in SPRR2F+ KCs, IL-17 pathway (x-axis) versus IL-23 (y-axis) inhibition, colored by significance category. Pearson r and n as indicated.

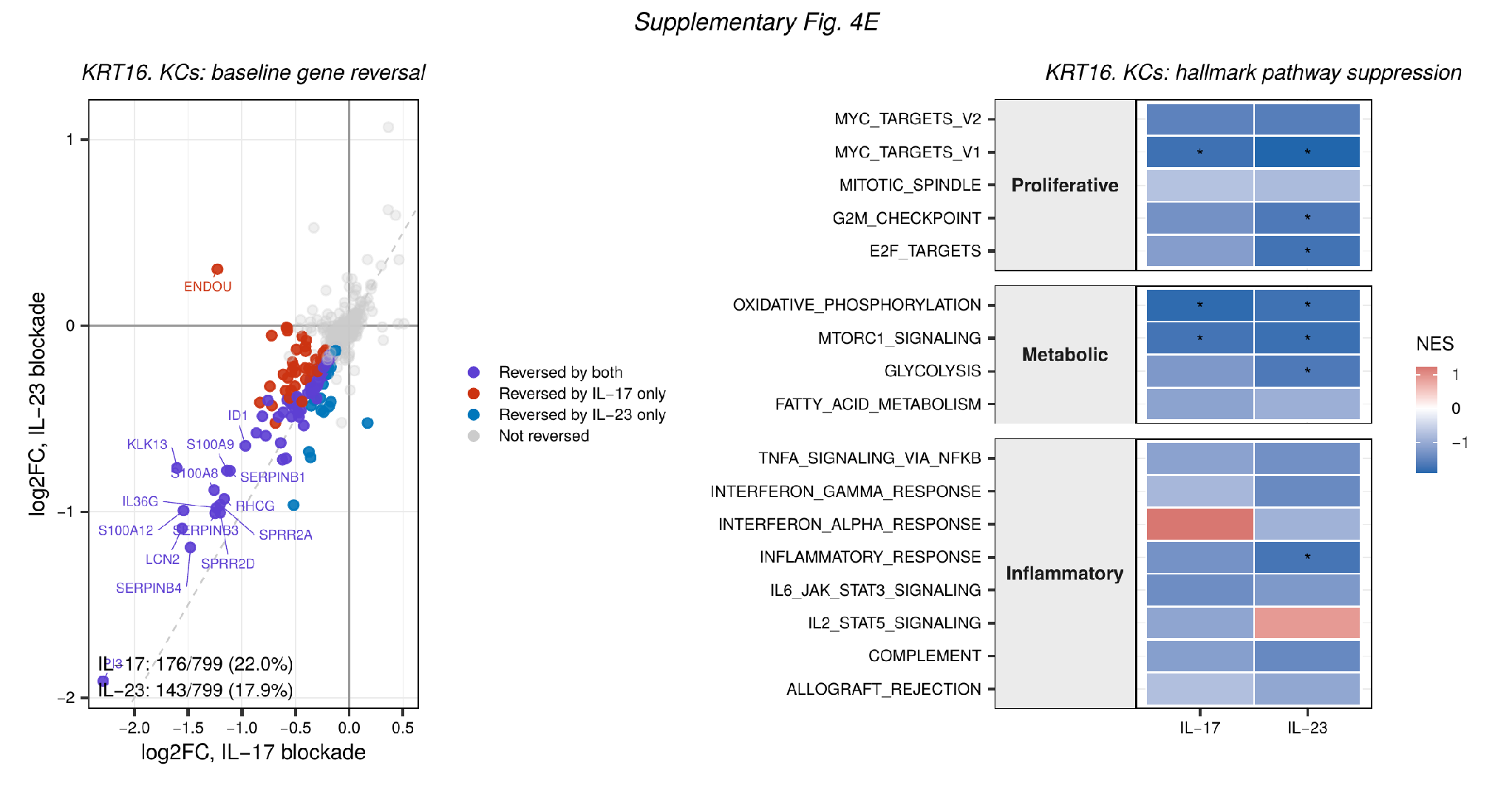

**Supplementary Fig. 4E | *KRT16+ KC gene reversal and hallmark pathway activity at Week 2.***

(Left) Scatter plot of log2FC under IL-17 pathway versus IL-23 inhibition for baseline-upregulated KRT16+ KC genes, colored by reversal status. (Right) Heatmap of hallmark gene-set normalized enrichment scores (NES) by pathway category and treatment; asterisks denote adjusted P < 0.05.

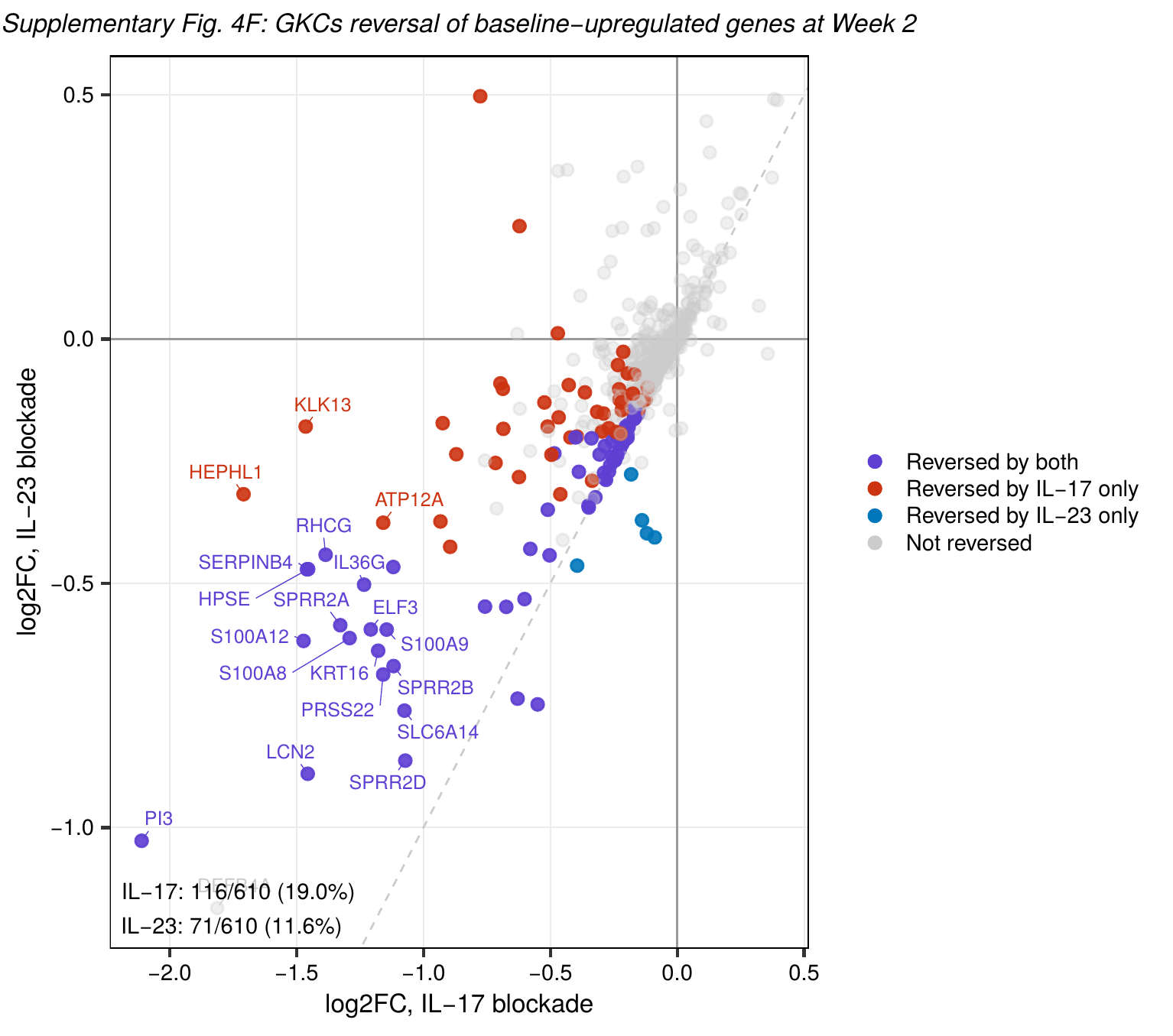

**Supplementary Fig. 4F | *GKC gene reversal at Week 2.***

Scatter plot of log2FC under IL-17 pathway versus IL-23 inhibition for baseline-upregulated GKC genes, colored by reversal status.

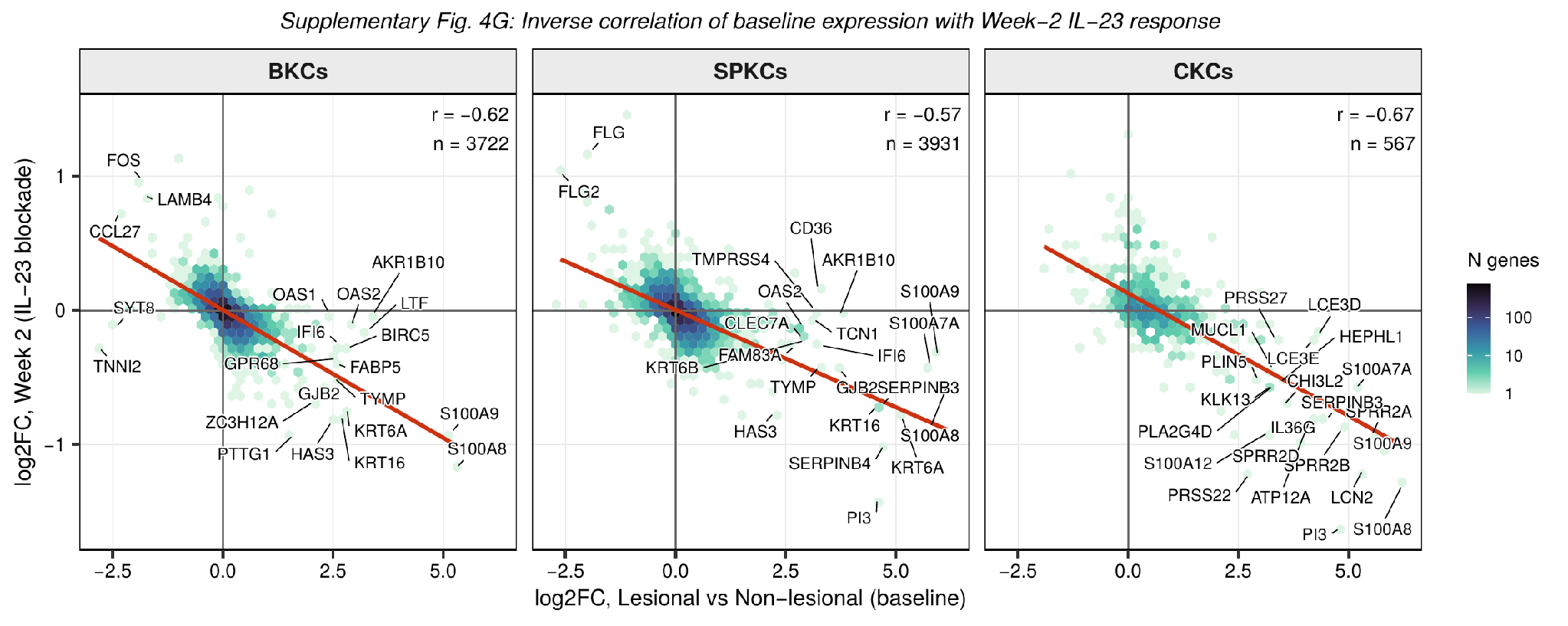

**Supplementary Fig. 4G | *Baseline lesional expression versus Week-2 IL-23 inhibition response in BKCs, SPKCs, and CKCs.***

Hexbin density scatter plots of baseline lesional log2FC (x-axis) versus week-2 log2FC under IL-23 inhibition (y-axis). Red line = linear fit; Pearson r and n given per cluster. Selected genes labeled.

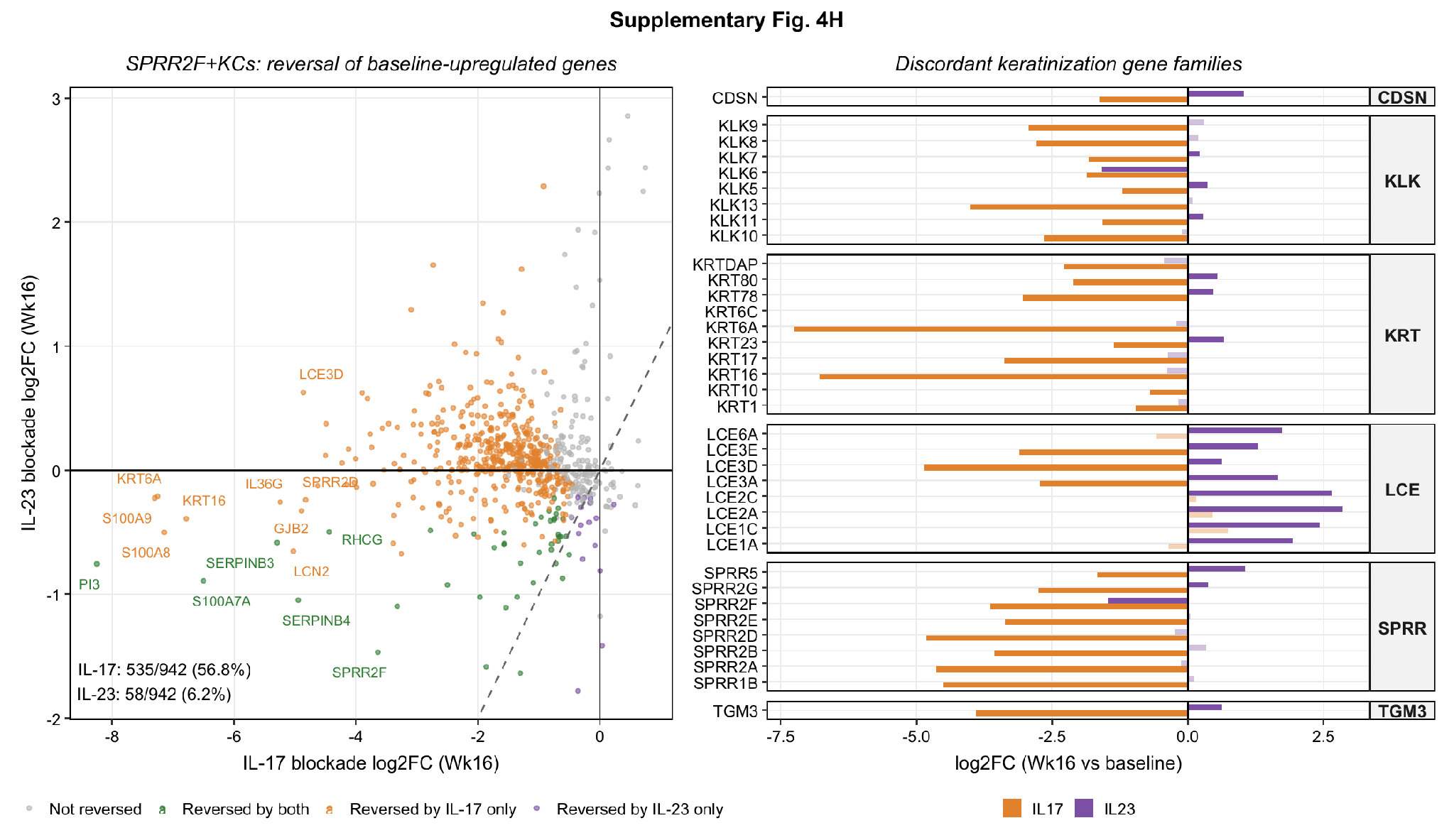

**Supplementary Fig. 4H | *SPRR2F+ KC gene reversal and keratinization gene families at Week 16.***

(Left) Scatter plot of log2FC under IL-17 pathway versus IL-23 inhibition at week 16 for baseline-upregulated SPRR2F+ KC genes, colored by reversal status. (Right) Paired bar chart of log2FC (week 16 vs baseline) under each treatment for genes within six keratinization-associated gene families (CDSN, KLK, KRT, LCE, SPRR, TGM3).

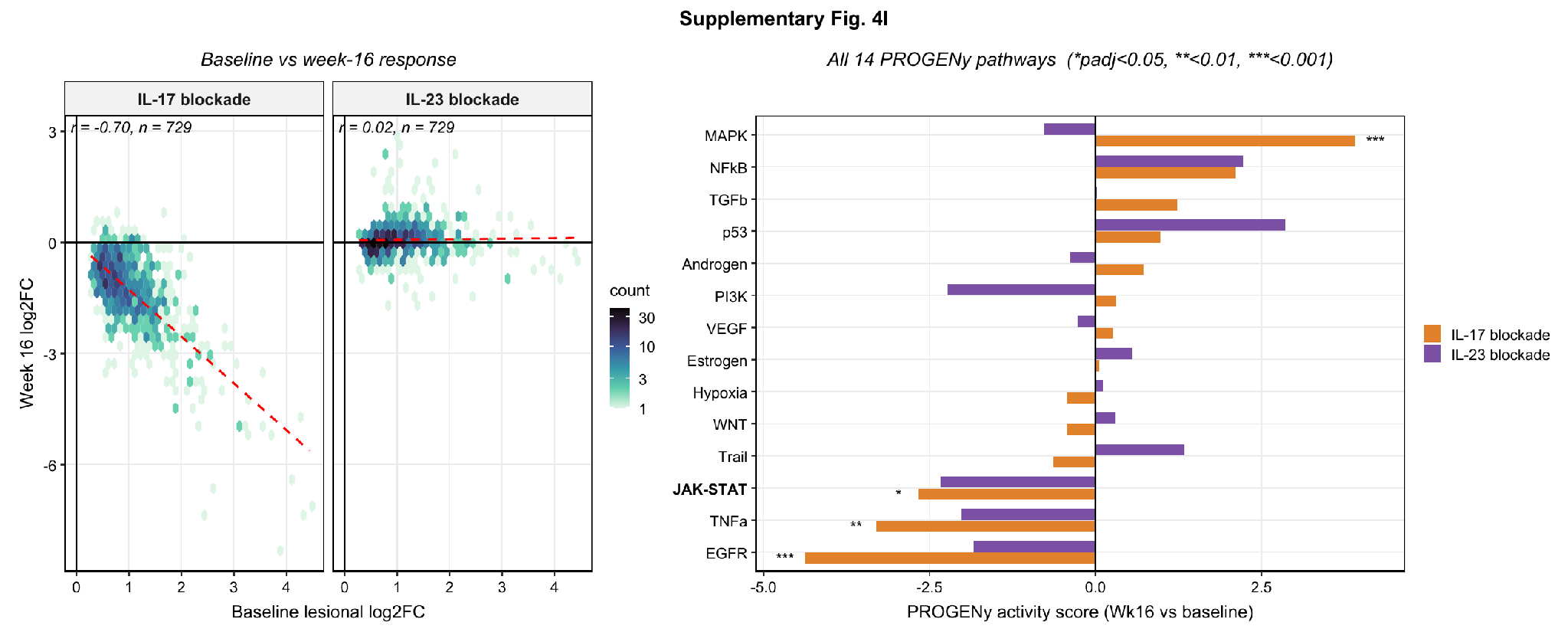

**Supplementary Fig. 4I | *SPRR2F+ KC baseline-versus-week-16 correlation and PROGENy pathway activity.***

(Left) Hexbin density scatter plots of baseline lesional log2FC versus week-16 log2FC in SPRR2F+ KCs, IL-17 pathway and IL-23 inhibition shown separately; dashed line = linear fit. (Right) Bar chart of PROGENy pathway activity scores (week 16 vs baseline) for all 14 pathways under each treatment; asterisks denote adjusted significance (*P < 0.05, **P < 0.01, ***P < 0.001).

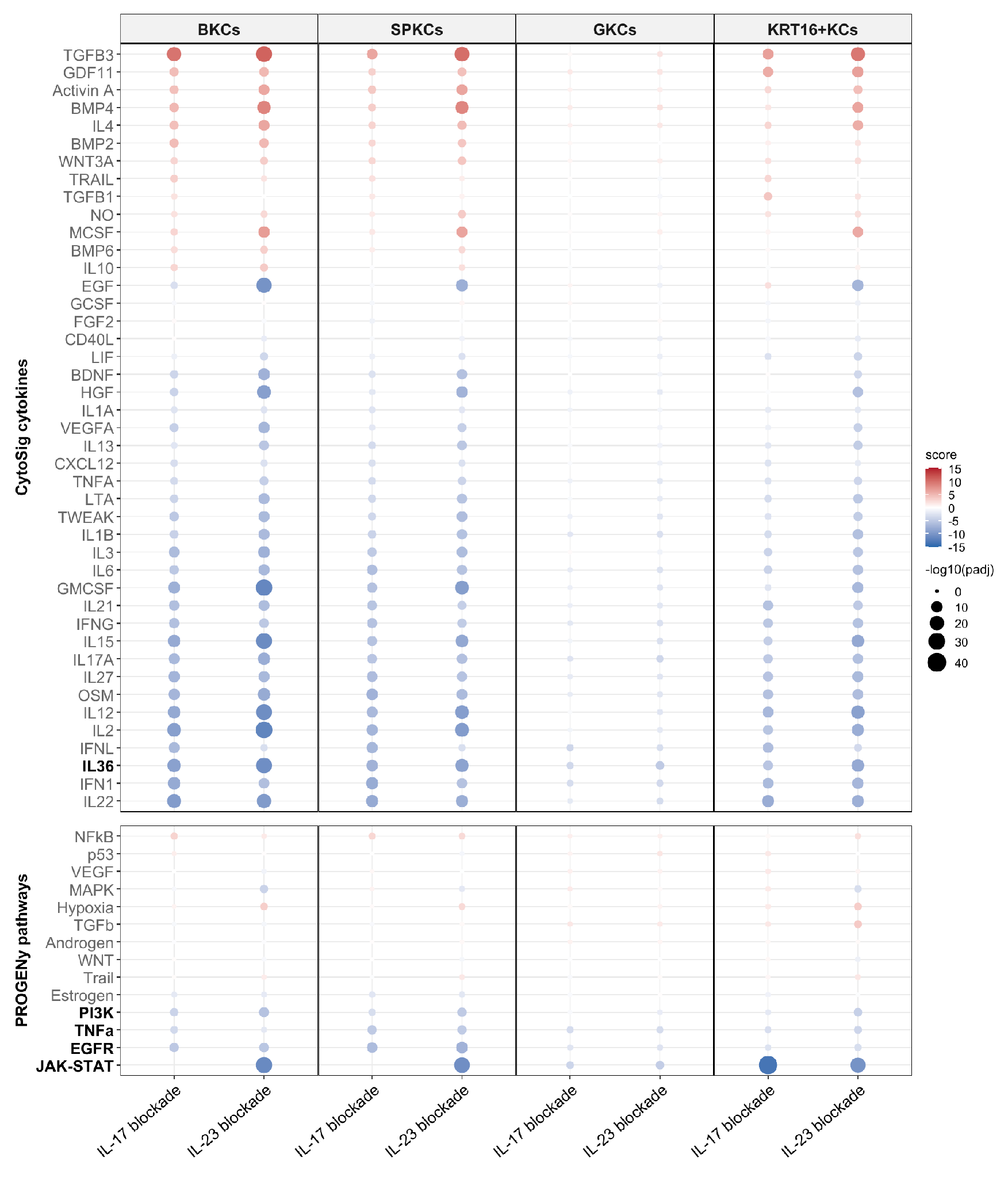

**Supplementary Fig. 4J | *Cytokine and pathway activity in BKCs, SPKCs, GKCs, and KRT16+ KCs at Week 16.***

Dot-plot matrix of CytoSig cytokine activity (top) and PROGENy pathway activity (bottom), by treatment. Dot color = activity score; dot size = -log10(adjusted P).

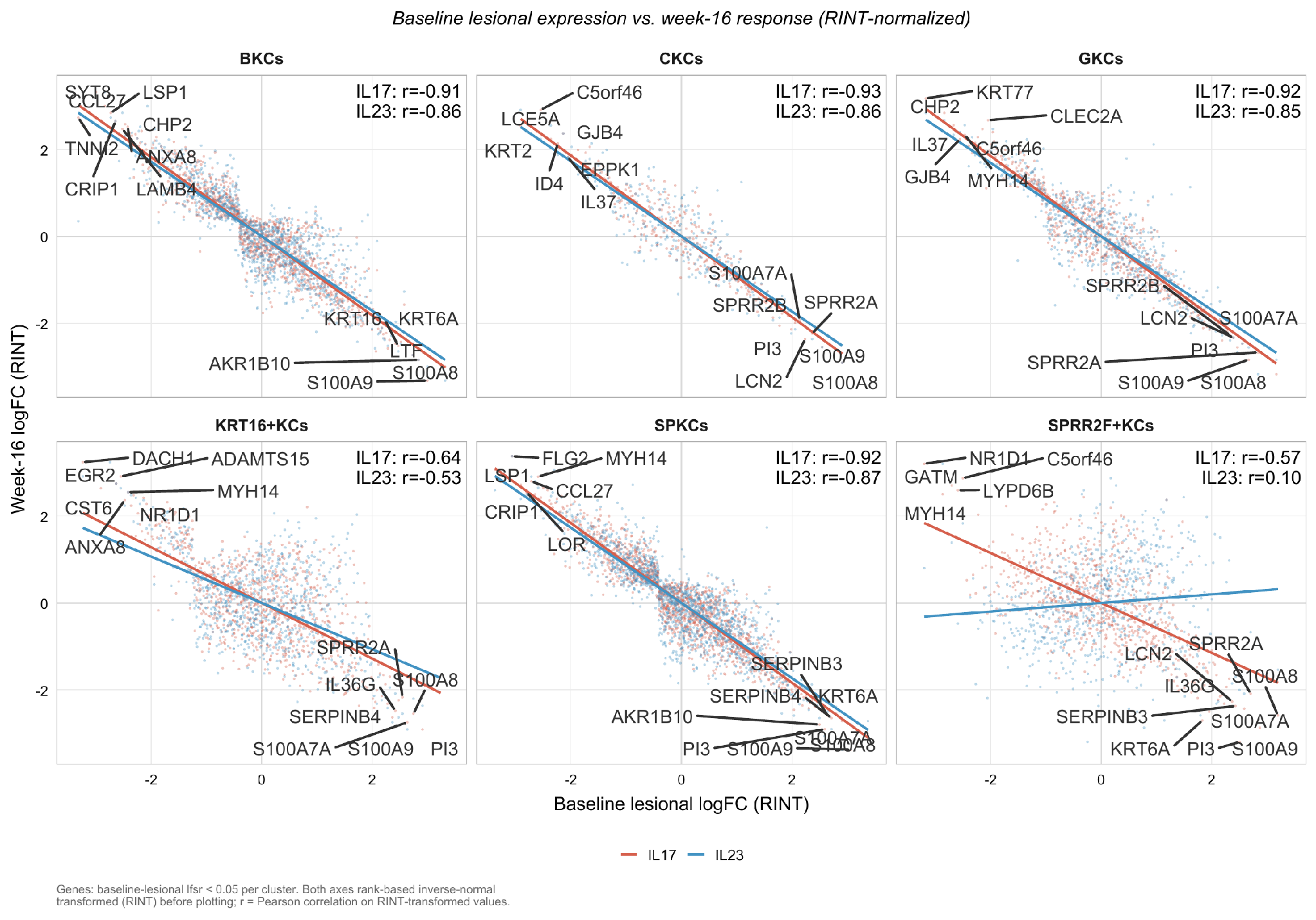

**Supplementary Fig. 4K | *Baseline lesional expression versus Week-16 response across KC clusters (RINT-normalized).***

Scatter plots of baseline lesional log2FC versus week-16 log2FC per KC cluster, IL-17 pathway (red) and IL-23 (blue) inhibition overlaid with linear fits. Genes: baseline-lesional lfsr < 0.05. Both axes rank-based inverse-normal transformed (RINT); r = Pearson correlation on RINT-transformed values.

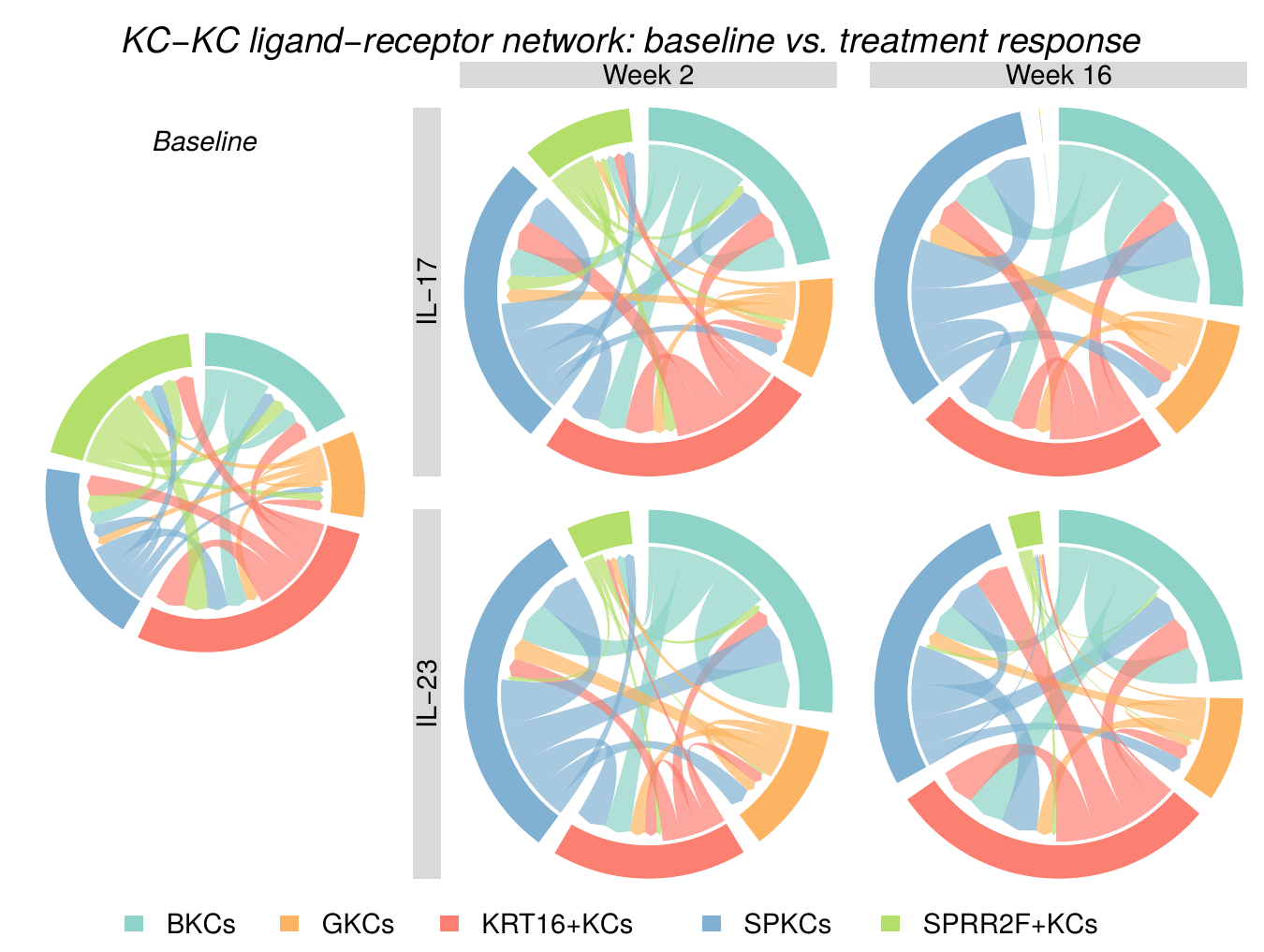

**Supplementary Fig. 4L | *KC-KC ligand-receptor communication network, baseline and treatment.***

Chord diagrams of the aggregate KC-KC ligand-receptor network among five KC subtypes at baseline and under IL-17 pathway or IL-23 inhibition at week 2 and week 16. Chord width = number of ligand-receptor pairs (MultiNicheNet prioritization score >= 0.6) between sender and receiver; arrows and differential height indicate directionality; color = KC subtype.

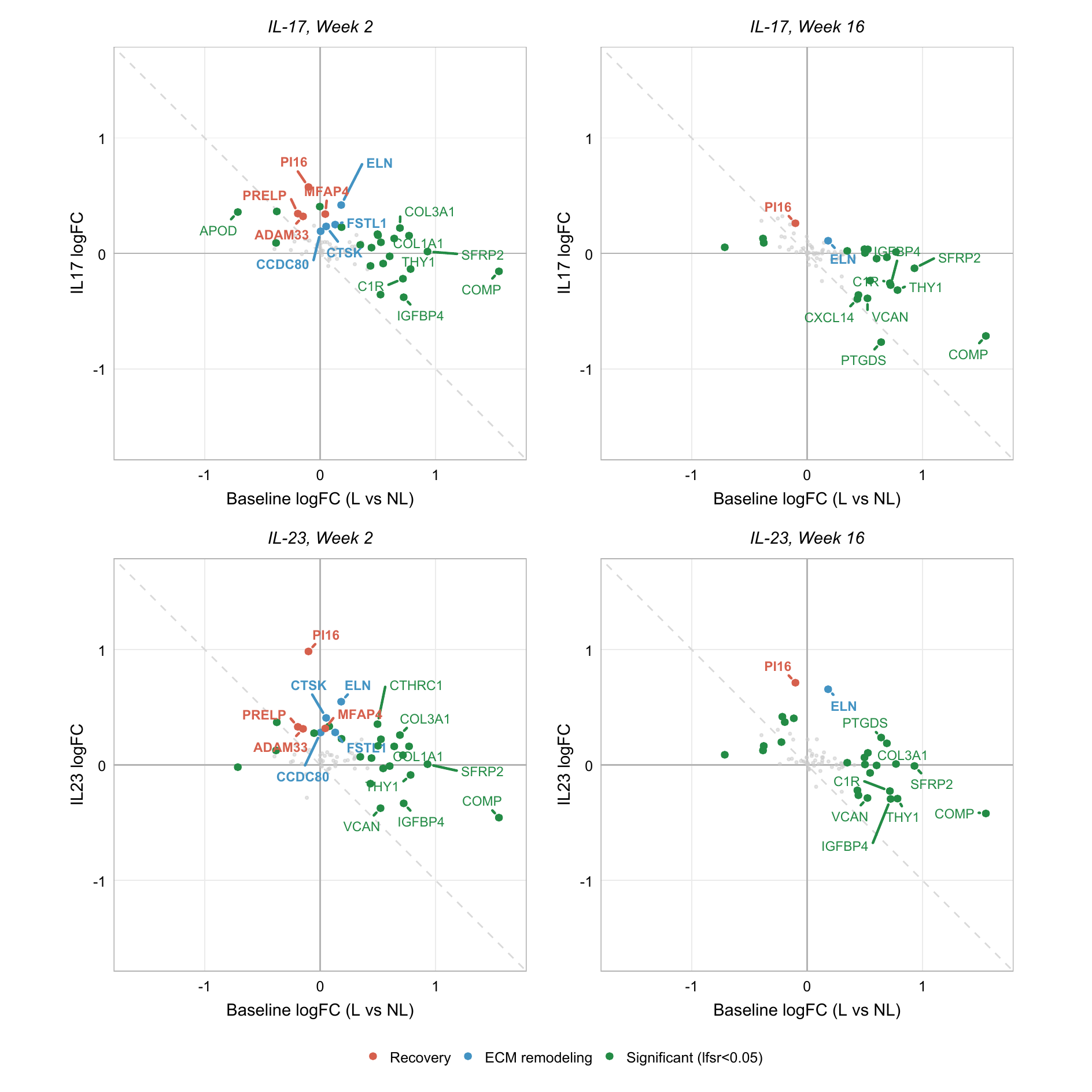

**Supplementary Fig. 4M | *FB transcriptional response, baseline versus treatment log2FC.***

Quadrant plots of baseline lesional log2FC (x-axis) versus treatment log2FC (y-axis) in FBs, IL-17 pathway (top) and IL-23 (bottom) inhibition, week 2 and week 16. Recovery genes (PI16, MFAP4, PRELP, ADAM33) and ECM-remodeling genes (CTSK, FSTL1, CCDC80, ELN) highlighted; other significant genes (lfsr < 0.05) shown in green.

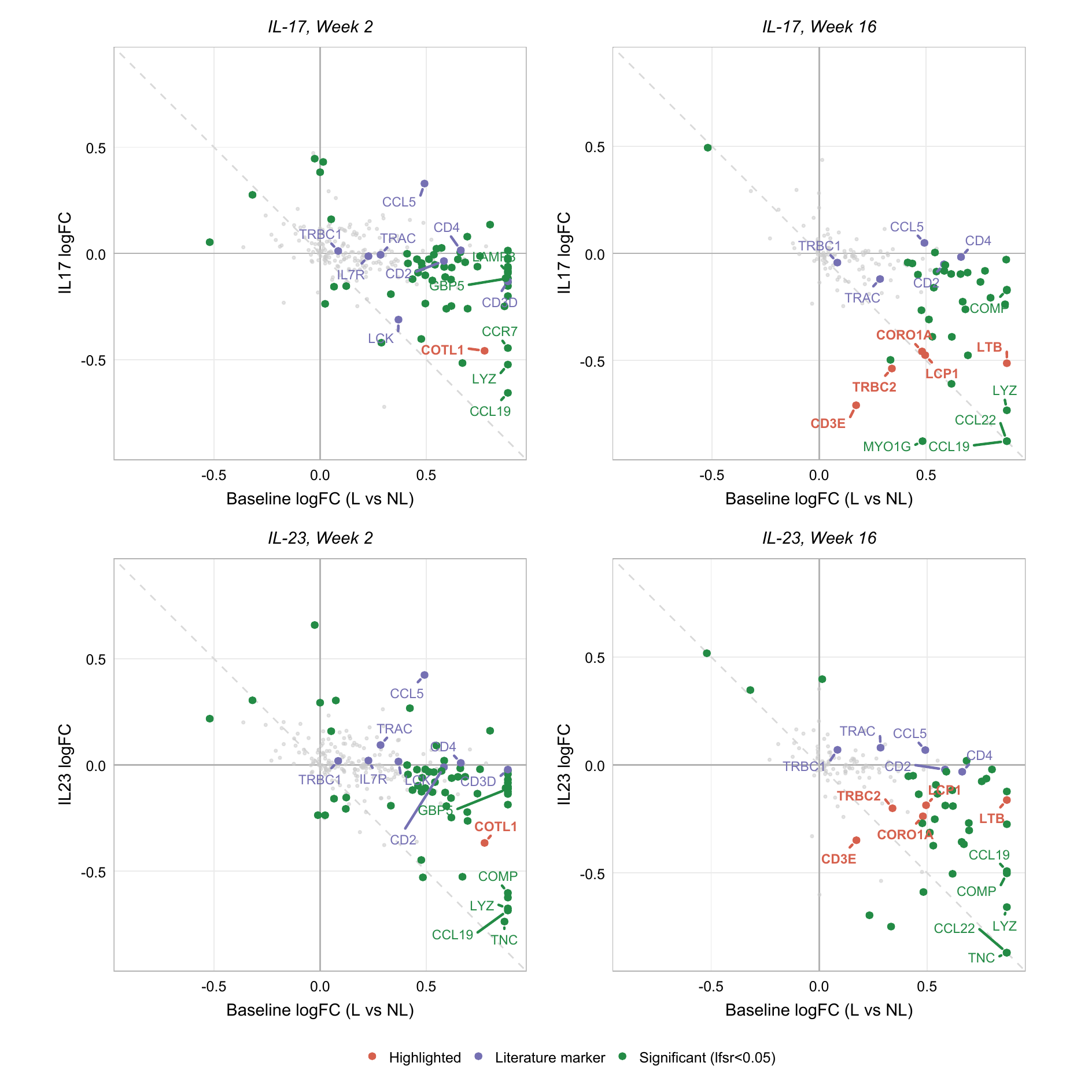

**Supplementary Fig. 4N | *T cell transcriptional response, baseline versus treatment log2FC.***

Quadrant plots as in Supplementary Fig. 4M, for T cells. Genes cited in the main text (COTL1, CD3E, LTB, TRBC2, CORO1A, LCP1) highlighted; canonical T cell markers (CD3D, CD2, TRAC, TRBC1, IL7R, CCL5, CD4, LCK, CD28) shown for reference; other significant genes in green. Immunoglobulin and collagen genes excluded.

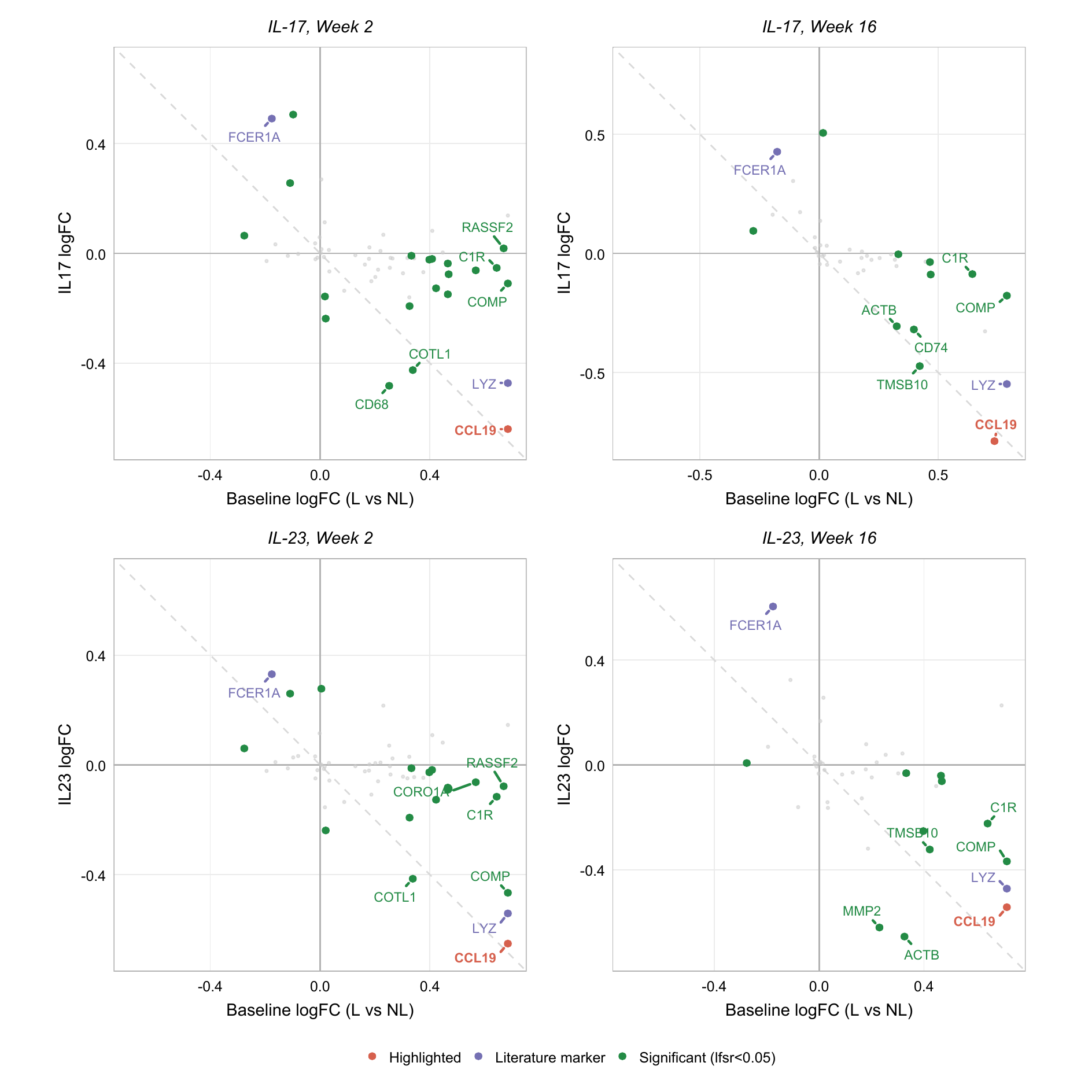

**Supplementary Fig. 4O | *DC transcriptional response, baseline versus treatment log2FC.***

Quadrant plots as in Supplementary Fig. 4M-N, for DCs. CCL19 highlighted; canonical DC markers (CD1C, FCER1A, LYZ) shown for reference; other significant genes in green. Immunoglobulin and collagen genes excluded.
