## Supplementary Figure 5 for "High definition longitudinal spatial atlas of psoriasis reveals SPRR2F⁺ keratinocytes and distinct tissue-resolution trajectories under IL-17 and IL-23 inhibition"

**Supplementary Figures**

*Supplementary Figure 5*

Supplementary Fig. 5A	Mouse and rabbit isotype control for S100A7/SPRR2F immunofluorescence.

Supplementary Fig. 5B	Mouse and rabbit isotype control for LL-37/SPRR2F immunofluorescence.

Supplementary Fig. 5C	Goat and rabbit isotype control for HBD-2/SPRR2F immunofluorescence.

Supplementary Fig. 5D	Single-antibody specificity controls for PI3/SPRR2F immunofluorescence.

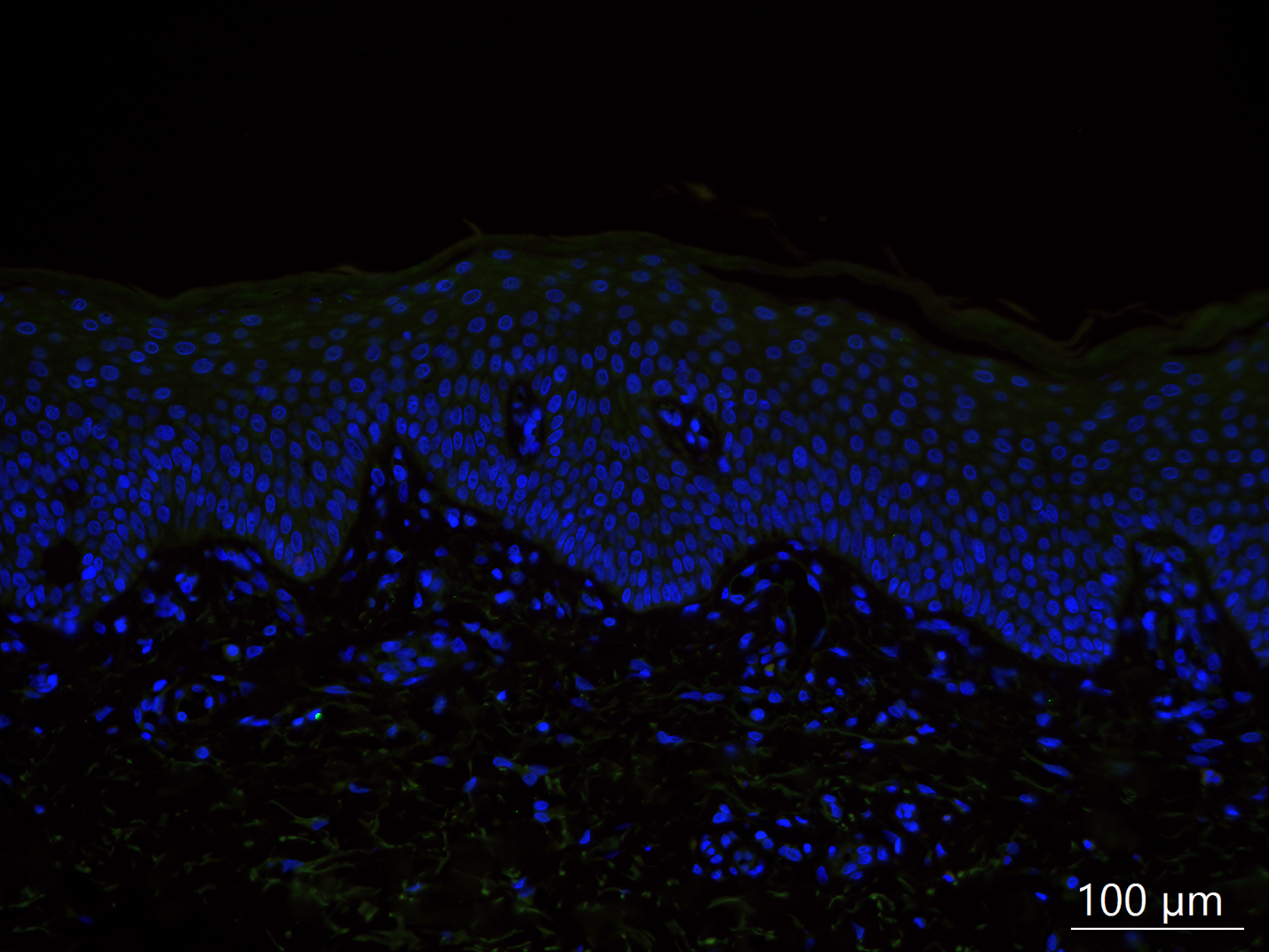

**Supplementary Fig. 5A | *Mouse and rabbit isotype control for S100A7/SPRR2F immunofluorescence.***

Combined mouse IgG (matched to the S100A7 primary antibody) and rabbit IgG (matched to the SPRR2F primary antibody) isotype control. White dashed line marks the dermal-epidermal junction. Scale bar, 100 µm.

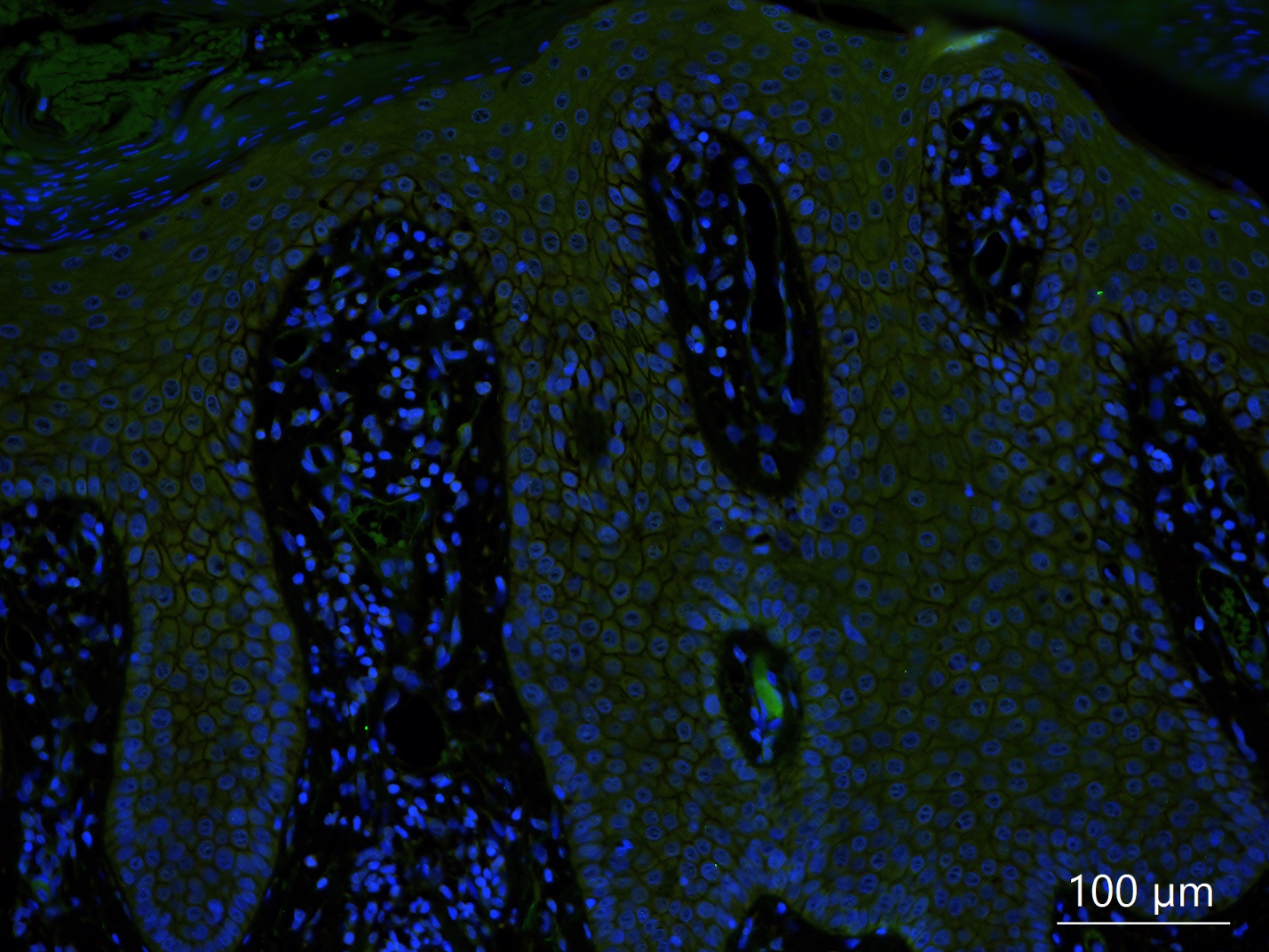

**Supplementary Fig. 5B | *Mouse and rabbit isotype control for LL-37/SPRR2F immunofluorescence.***

Combined mouse IgG (matched to the LL-37 primary antibody) and rabbit IgG (matched to the SPRR2F primary antibody) isotype control. White dashed line marks the dermal-epidermal junction. Scale bar, 100 µm.

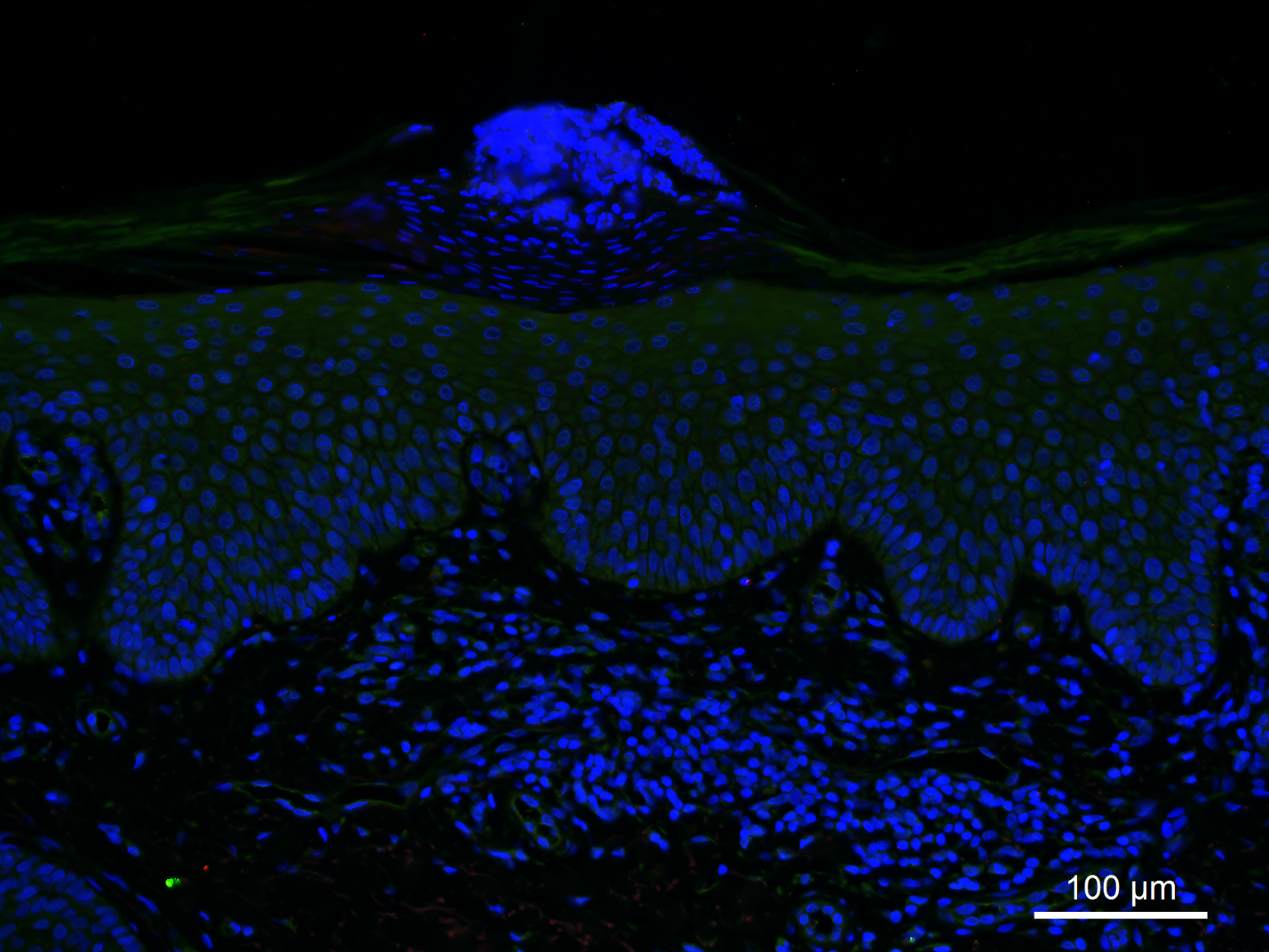

**Supplementary Fig. 5C | *Goat and rabbit isotype control for HBD-2/SPRR2F immunofluorescence.***

Combined goat IgG (matched to the HBD-2 primary antibody) and rabbit IgG (matched to the SPRR2F primary antibody) isotype control. White dashed line marks the dermal-epidermal junction. Scale bar, 100 µm.

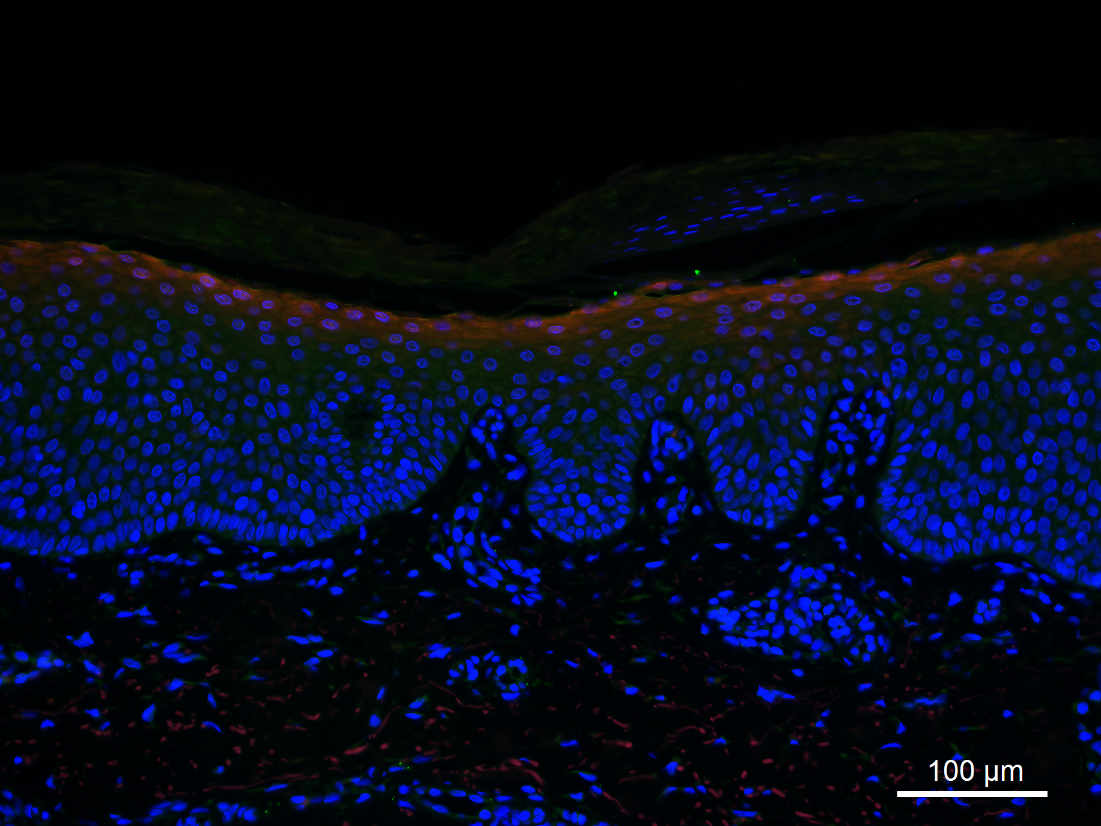

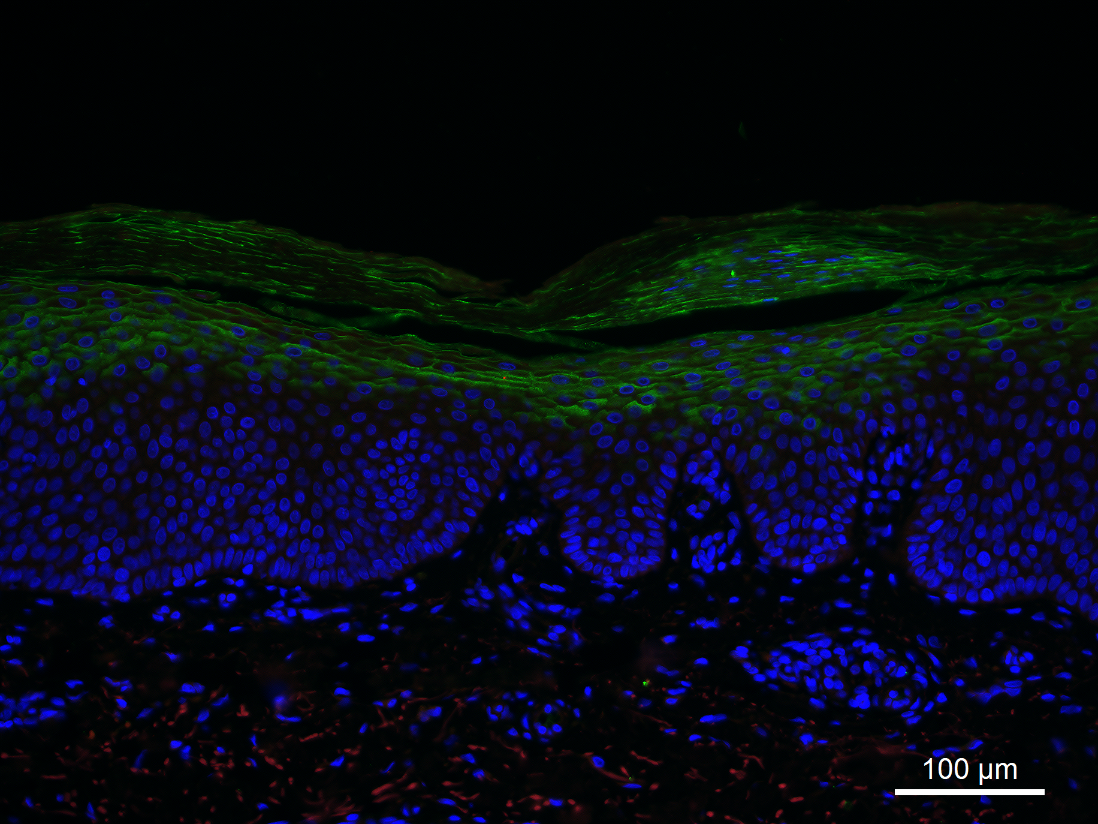

**Supplementary Fig. 5D | *Single-antibody specificity controls for PI3/SPRR2F immunofluorescence.***

Representative single-antibody isotype-swap controls for the PI3/SPRR2F co-stain. Upper: SPRR2F (red) with a rabbit IgG isotype control substituted for the PI3 primary antibody (green channel). Bottom: PI3 (green) with a rabbit IgG isotype control substituted for the SPRR2F primary antibody (red channel). White dashed line marks the dermal-epidermal junction. Scale bars, 100 µm.
