## Supplementary Figure 6 for "High definition longitudinal spatial atlas of psoriasis reveals SPRR2F⁺ keratinocytes and distinct tissue-resolution trajectories under IL-17 and IL-23 inhibition"

**Supplementary Figures**

*Supplementary Figure 6*

Supplementary Fig. 6A	SPRR2F immunofluorescence staining in plaque psoriasis patients treated with IL-17A and IL-17RA inhibitors.

Supplementary Fig. 6B	Rabbit isotype control for SPRR2F immunofluorescence.

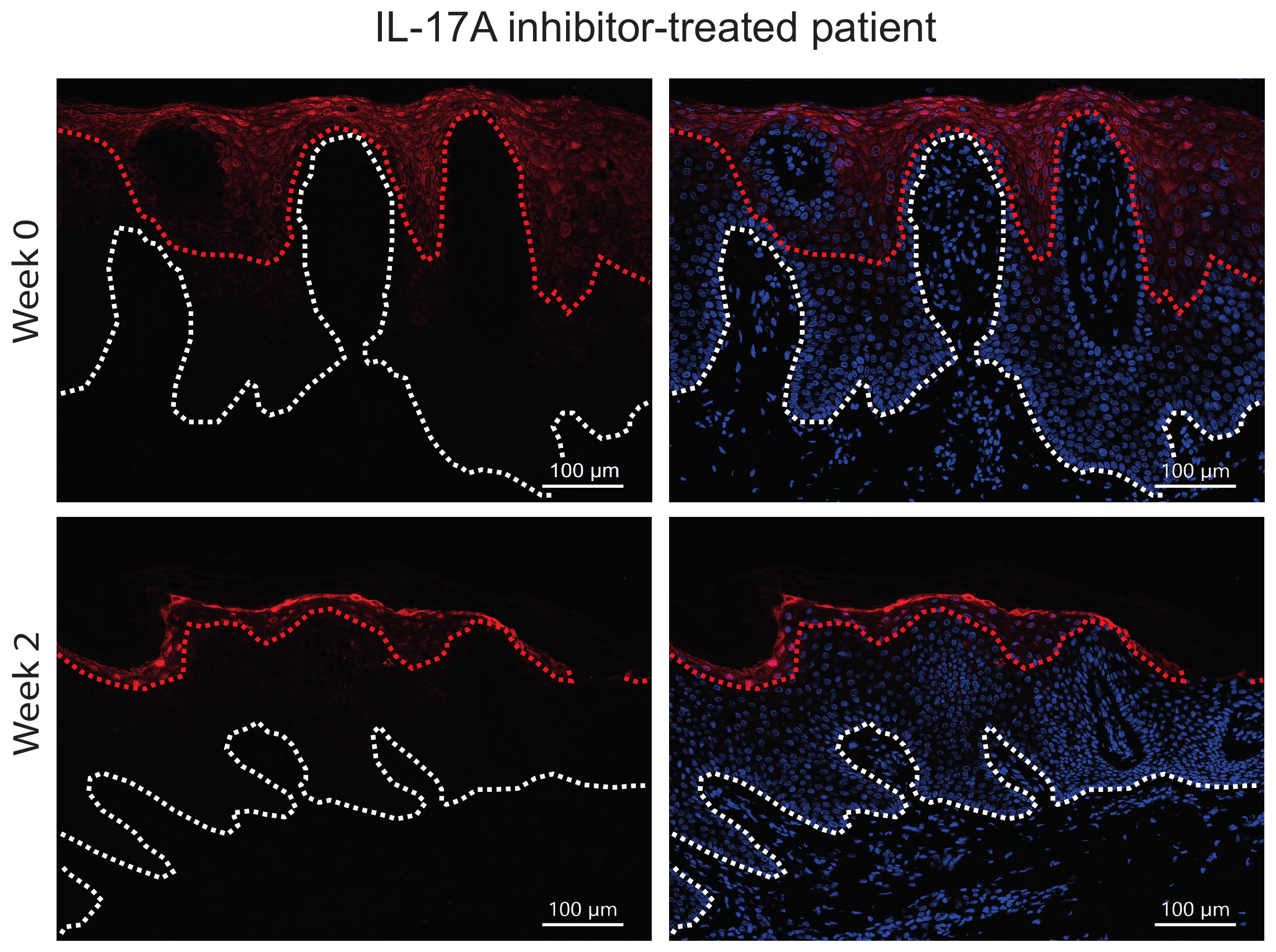

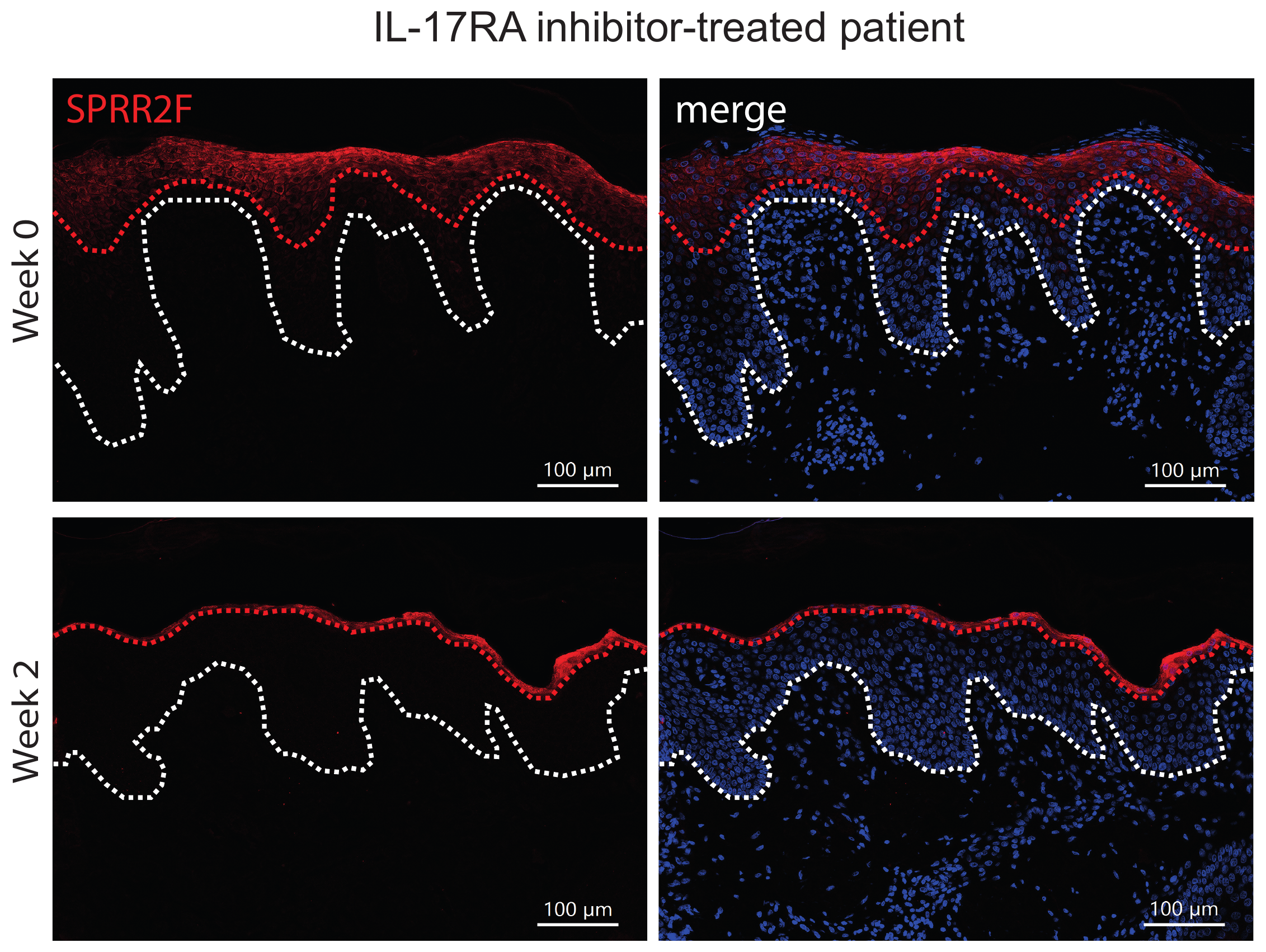

**Supplementary Fig. 6A | *SPRR2F immunofluorescence staining in plaque psoriasis patients treated with IL-17A and IL-17RA inhibitors.***

Immunofluorescence staining of SPRR2F (red) in lesional skin of one patient treated with an IL-17A inhibitor (ixekizumab) and one patient treated with an IL-17RA inhibitor (brodalumab) at baseline (week 0) and week 2. Left, SPRR2F; right, merge with DAPI (blue). Red dotted line marks the lower boundary of the SPRR2F-positive epidermal layer; white dotted line marks the dermal–epidermal junction. Scale bars, 100 µm.

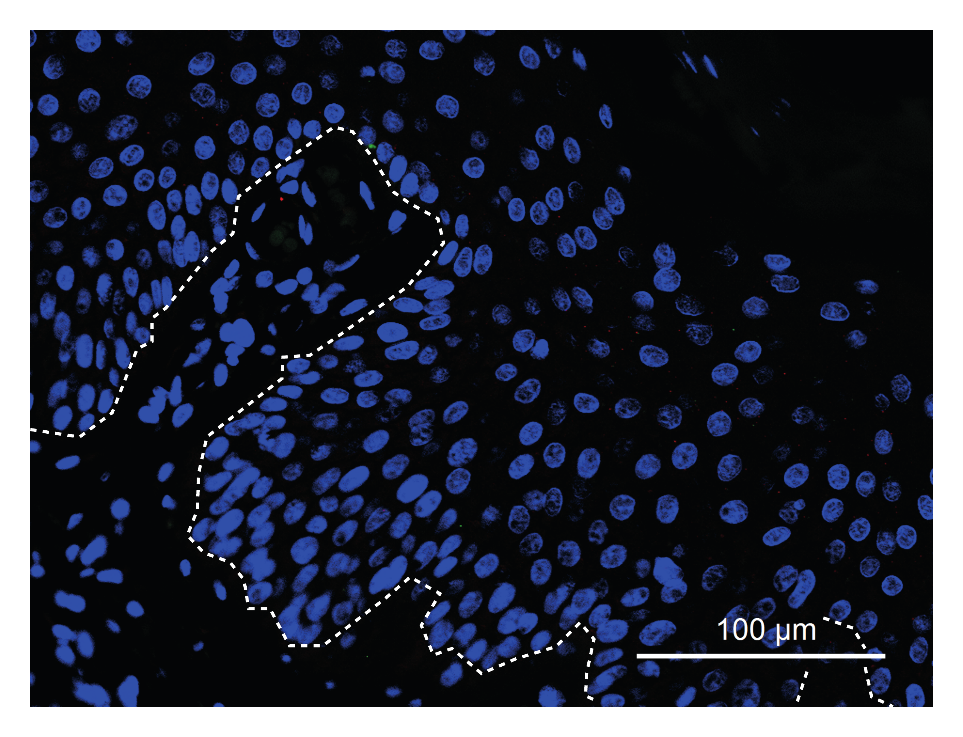

**Supplementary Fig. 6B | *Rabbit isotype control for SPRR2F immunofluorescence.***

Rabbit IgG isotype control matched to the SPRR2F primary antibody. Nuclei were counterstained with DAPI (blue). White dashed line marks the dermal–epidermal junction. Scale bar, 100 µm.
